# Single-cell dissection of immune-state architecture across diverse human population cohorts

**DOI:** 10.64898/2026.11.17.26366584

**Authors:** Edoardo Giacopuzzi, Olli Dufva, Alessandro Raveane, Jan P. Engelmann, Manuel Tardáguila Sancho, Daniela Zanotti, Giuditta Clerici, Stephen Watt, Matiss Ozols, Arianna Landini, Paola Benaglio, Lucas Arnoldt, Beatrice Costa, Bruno Ariano, Sodbo Sharapov, Danai Vagiaki, Davide Bolognini, Eva Holtkamp, Shuang Li, Ruth Eberhardt, Andrea Senacheribbe, Andrei Cristea, Emanuele Saitto, Eljas Roellin, Dominique Von Schiller, Eve Coomber, May Hu, Gabriella Ivanova, Fereshteh Torabi, Nima Subasinghe, Laura Durrant, Vahid Shariati, Chiara Chiereghin, Michela Colombo, Ilaria Gori, Federica Santonastaso, Matilde I. Conte, Cecilia Dominguez Conde, Alice Giustacchini, Genes & Health Research Team, Nicole Staudt, John Wright, Richard Trembath, Carl A. Anderson, Blagoje Soskic, Craig A. Glastonbury, Julien Gagneur, Sarah Finer, Brian Clarke, Francesco Paolo Casale, Marc Jan Bonder, Fabian J. Theis, Fabiola Curion, Vivek Iyer, Oliver Stegle, David A. van Heel, Sarah A. Teichmann, Nicola Pirastu, Nicole Soranzo

**Author notes:** These authors contributed equally to the manuscript (Edoardo Giacopuzzi, Olli Dufva, Alessandro Raveane, Jan P. Engelmann, Manuel Tardáguila Sancho, Daniela Zanotti, Giuditta Clerici, Stephen Watt, Matiss Ozols). These authors contributed equally to the manuscript (Fabian J. Theis, Fabiola Curion, Vivek Iyer, Oliver Stegle, David A. van Heel, Sarah A. Teichmann, Nicola Pirastu, Nicole Soranzo). Correspondence to be addressed to NS, Fondazione Human Technopole, Via Levi Montalcini 1, 20157 Milano, Italy. D.A.v.H., Blizard Institute, Queen Mary University of London, London, United Kingdom. SAT., Department of Medicine, Cambridge Stem Cell Institute, University of Cambridge, Cambridge, UK,. OS., Division of Computational Genomics and Systems Genetics German Cancer Research Center (DKFZ) Heidelberg, Germany.

## Abstract

The extension of human lifespan is accompanied by a growing burden of chronic disease and multimorbidity. The immune system, whose functional state is continuously remodelled by ageing, environmental exposures, infection history and disease, is pivotal to respond to these challenges. However, how these factors jointly influence immune variation, regulatory mechanisms and disease susceptibility across populations remains poorly understood. Here we present an outline and first results of the CARDINAL (Cellular Architecture of immune-state Regulation, Disease, and Immune Networks across the Adult Lifecourse) project. We generated a new population-scale single-cell atlas of peripheral blood comprising almost 10 million immune cells from 6,003 individuals across two UK cohorts with integrated genomic and longitudinal electronic health record data: the UK Biobank (predominantly European ancestry British) and Genes & Health (British Bangladeshi and British Pakistani). We identify immune-state architectures, shared across cohorts, associated with ageing, chronic viral exposure, inflammatory activation and premalignant conditions. We show that demographic and lifestyle factors induce largely conserved immune programmes, while chronic cardiometabolic diseases are associated with cohort-dominant cellular programmes and transcriptional states. Disentangling of genetic and clinical influences reveals coordinated regulatory programmes associated with metabolic, inflammatory and infectious remodelling. These programmes are observed in specific disease contexts and immune states, such as those induced by chronic viral exposure, often independently of genetic differences. Together, our findings support a model of immune-state architecture shaped by ageing, environment, disease history and genetic variation, which can be read from population-specific regulatory landscapes.

## Introduction

The increasing burden of chronic disease and multimorbidity in ageing populations presents a major challenge for global health^1,2^. Changes in immune function are central to this burden, linking declining protection against infection with chronic inflammation, cardiovascular disease, neurodegeneration, cancer and impaired tissue repair^3,4^. Immune composition and function vary substantially between individuals, reflecting the combined influence of inherited genetic variation and lifelong environmental, metabolic, infectious and physiological exposures^5–9^. Understanding how these influences shape immunity is essential to defining its contribution to human health and disease.

Single-cell genomics now enables immune cell composition, transcriptional states and genetic regulation to be studied at population scale^10–15^. These studies have revealed age-associated remodelling of immune compartments, genetically regulated cellular states and context-dependent regulatory effects^10–14,16–21^. However, most have focused on ageing^17,19–22^ or genetic regulation within relatively homogeneous populations^10,14,16^. Human populations differ in genetic background and in their histories of infection, lifestyle, metabolic stress and social conditions, with consequences for immune phenotypes and the cellular contexts in which genetic variants act^23–25^. Studying population cohorts with distinct genetic and exposure histories therefore offers an opportunity to distinguish shared features of immune organisation from those associated with population context. How these influences jointly relate to immune composition, regulatory variation and chronic disease remains poorly understood.

Here we present CARDINAL (Cellular Architecture of immune-state Regulation, Disease, and Immune Networks across the Adult Lifecourse), a single-cell transcriptomic atlas of peripheral blood mononuclear cells from 6,003 individuals across two UK cohorts representing three distinct ethnic groups, integrated with genetic and longitudinal health data. We demonstrate the value of this integrated resource for investigating immune architecture across scales, from blood cell composition to finely resolved cellular states and gene regulatory programmes. We systematically map shared and cohort-specific immune signatures associated with demographic factors, environmental exposures, infection history and chronic disease, and examine how disease and infection history shape the cellular contexts in which genetic regulatory effects are detected. Companion papers extend these analyses through detailed characterisation of immune cell composition^26^, genetic regulation of gene expression and pleiotropy^27^, blood archetypes and extreme blood states^28^ and predictive cellular programme modelling^29^. Together, these findings establish a framework for understanding how genetic variation and life-course exposures jointly shape immune architecture and disease across populations, revealing that population differences in disease biology arise not only from differences in allele frequencies but also from environmentally conditioned immune states that modify the effects of genetic variation.

## Results

### A population-scale single-cell atlas of peripheral blood across multiethnic UK cohorts

We generated a dataset of ∼10 million peripheral blood mononuclear cells from 6,003 recalled participants across two large-scale UK cohorts: the UK Biobank (UKB) ^30^ and Genes & Health (G&H) ^31^ (**Fig. 1a**). The cohorts differ in ancestry composition, as assessed using genetic and self-reported data (**Methods**). UKB participants (*N*= 4,754) are predominantly of European ancestry (N= 4,594; 96% White British), with 160 donors of diverse non-European ancestries. The G&H recalled participants included 1,249 individuals of South Asian ancestry, of which 741 are of British Bangladeshi origin, 504 are of British Pakistani origin, and four are from other population groups (**Fig. 1b,c** and **Supplementary Table 1**). These volunteers from the two cohorts exhibit distinct demographic profiles (median UKB age at recall = 67 years, range 53–84 and median G&H age at recall = 42, range 16–89), risk factors and prevalence of major diseases (**Supplementary Table 1** and **Supplementary Table 2**), with greater prevalence of neoplasms and cardiovascular diseases in UKB, and greater prevalence of infectious, metabolic and dermatological disorders in G&H, reflective of cohort-specific age distribution and risk factors. Blood samples were processed through a dedicated automated workflow **(Supplementary Fig. 1a, Methods**), with PBMCs isolated from fresh blood for ∼95% of samples to minimize cryopreservation artefacts ^32^ (**Supplementary Table 3**). Additionally, parallel complete blood counts (CBCs) were obtained for 92% of donors using a Sysmex XN-1000 analyser and integrated within the single-cell data pipeline. Across the full dataset, we applied a scalable quality control (QC) framework combining ambient RNA removal^33^, multi-method doublet detection^34–36^, and stringent donor deconvolution across 634 sequencing pools, followed by donor-level and cell-level adaptive QC (**Supplementary Fig. 2**; **Methods**). Assignment of cells to donors was validated through complementary genotype-unaware deconvolution^37^, jackknife subsampling, and read-level concordance analyses, ensuring high-confidence cell–donor pairings (**Methods**).

**Figure 1:**
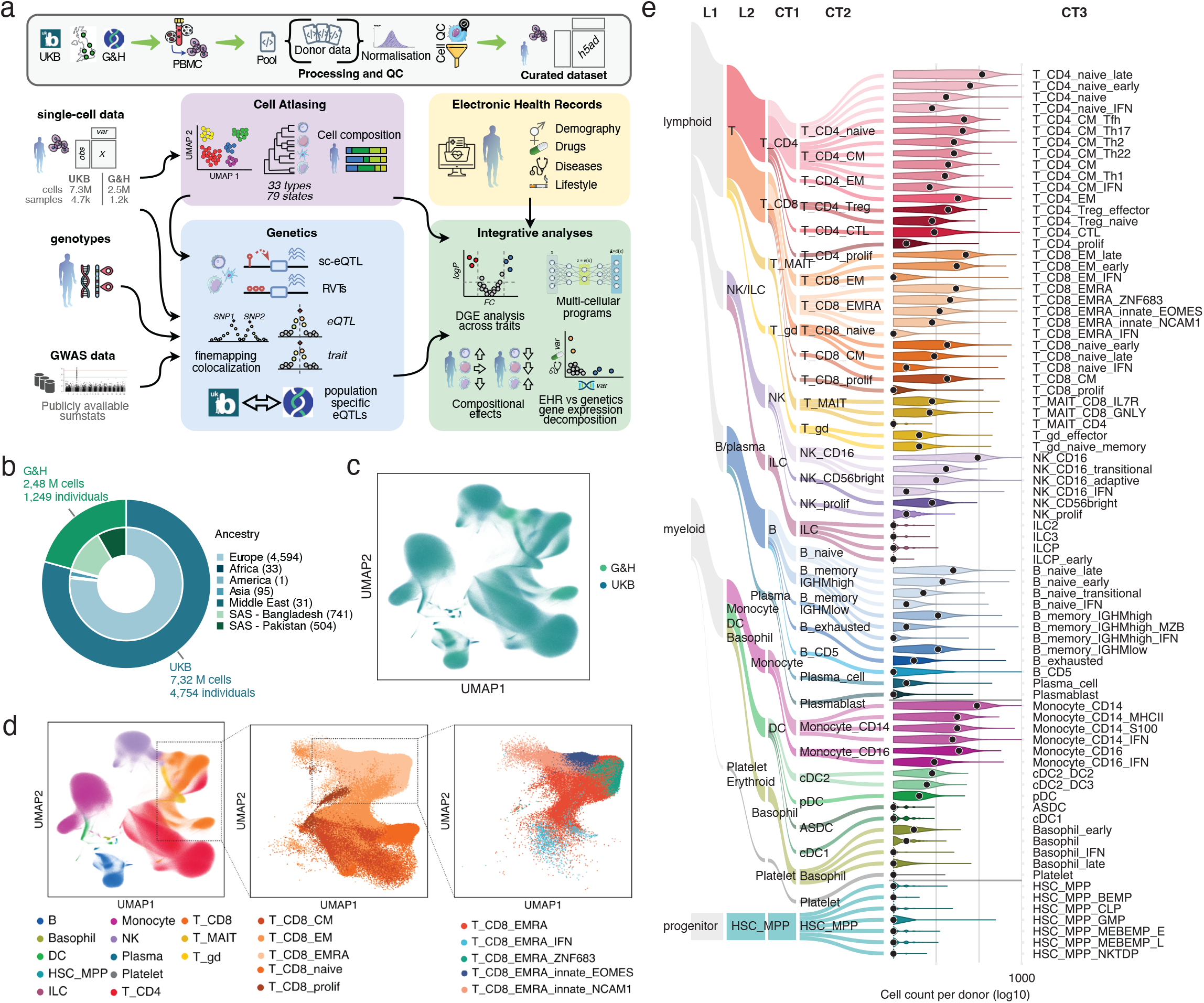
Integrated single-cell atlas and genetic analysis of human immune cell populations. **a**, Overview of the study design and analytical workflow. Single-cell RNA-seq data from multiple cohorts were processed through quality control, normalization and integration to generate a unified cell atlas. Downstream analyses included cell-type annotation and composition, genetic association analyses (including eQTL mapping and colocalization), and integration with electronic health record (EHR) data to link cellular phenotypes with disease traits and drug exposures. **b**, Cohort composition and integration. Donut chart shows the distribution of individuals and cells across cohorts and ancestry groups. **c**, UMAP projection of all cells coloured by cohort demonstrates effective integration across datasets. **d**, UMAP embeddings of immune cell populations. Left, global clustering of major cell types. Middle and right, higher-resolution embeddings of CD8+ T cell subsets, highlighting distinct functional states including naive, central memory, effector memory, proliferating, and interferon-responsive populations. **e**, Hierarchical organization and abundance of immune cell subtypes. Lineage relationships across lymphoid and myeloid compartments are shown alongside violin plots of cell counts per donor (log10 scale). Points indicate individual donors. Diverse immune subsets, including T, NK, B, monocyte, and dendritic cell populations, exhibit substantial inter-individual variability in abundance.

High-quality UKB cells were then used to construct a reference atlas based on scANVI embeddings^38^, selected after systematic benchmarking of integration approaches, to remove major batch effects related to sex and experimental factors, ensuring a uniform representation across the two cohorts (**Supplementary Fig. 3**). Initial cell type labels were obtained by mapping cells to reference cell atlases ^39–41^, and then manually curated from scANVI embeddings, using Leiden clustering and marker gene expression (**Supplementary Fig. 4**, **Supplementary Fig. 5**). Cell annotations were structured hierarchically across five levels, ranging from lineage to cell state, to reflect biological lineage and cell-type granularity: three major lineages (L1; e.g., lymphoid, myeloid, progenitor), six broad lineage subtypes (L2; e.g., B/plasma, T cells), 13 major cell types (CT1; e.g., B cells, CD4^+^ T cells, CD8^+^ T cells), 32 classical cell subtypes (CT2; e.g., effector memory (EM) CD8^+^ T cells, central memory (CM) CD8^+^ T cells, effector memory re-expressing CD45RA (EMRA) CD8^+^ T cells), and 79 distinct cell states (CT3; e.g., ZNF683^+^ EMRA CD8^+^ T cells, EOMES^+^ innate-like EMRA CD8^+^ T cells, NCAM1^+^ innate-like EMRA CD8^+^ T cells) (**Fig. 1d,e**). Leveraging the UKB as reference atlas, labels from L1, L2, CT1 and CT2 were transferred from UKB to G&H using scArches^42^, after which a high-quality full dataset was manually annotated for CT3 cell states. A complete summary of the hierarchical cell types is reported in **Supplementary Table 4** and summary of cell and sample counts for each cell type in **Supplementary Table 5**.

Overall, we obtained high-quality data for 94% of donors, resulting in a final atlas of 9.8 million high-quality cells from 6,003 individuals (7.32M cells from 4,754 donors in UKB; 2.48M cells from 1,249 donors in G&H; **Supplementary Fig. 1b**). Additional filters based on inferred ancestry and sample preparation (**Methods**) were applied for analysis where appropriate (**Supplementary Table 6**).

### High-resolution blood profiling identifies distinct immune composition signatures across populations

We first used the atlas to illustrate variation in immune-state architecture across the two cohorts, from individual cell states to coordinated patterns of blood cell composition (**Fig. 2**). These patterns, their determinants and associated clonal processes are characterised in detail in the companion study by Dufva et al.^28^. At the finest annotation level (CT3), cell states ranged from common (found in >90% donors at minimum frequency of 0.1% out of total cells), restricted (found in 10-90% donors) and rare (found in <10% donors) (**Extended Data Fig. 1**). Several differed markedly in abundance between cohorts: UKB showed greater representation of CD16+ and MHCIIhigh monocytes, central memory T cells, conventional CD16+ NK cells and CD5+ B cells, whereas G&H showed enrichment of naïve T cells, several B naive and B memory cell types, innate lymphoid cell (ILC) populations, innate-like EMRA CD8+ T cells and adaptive NK cells (**Fig. 2b**). These differences illustrate the variation captured by the atlas across cohorts with distinct demographic and exposure histories.

**Figure 2:**
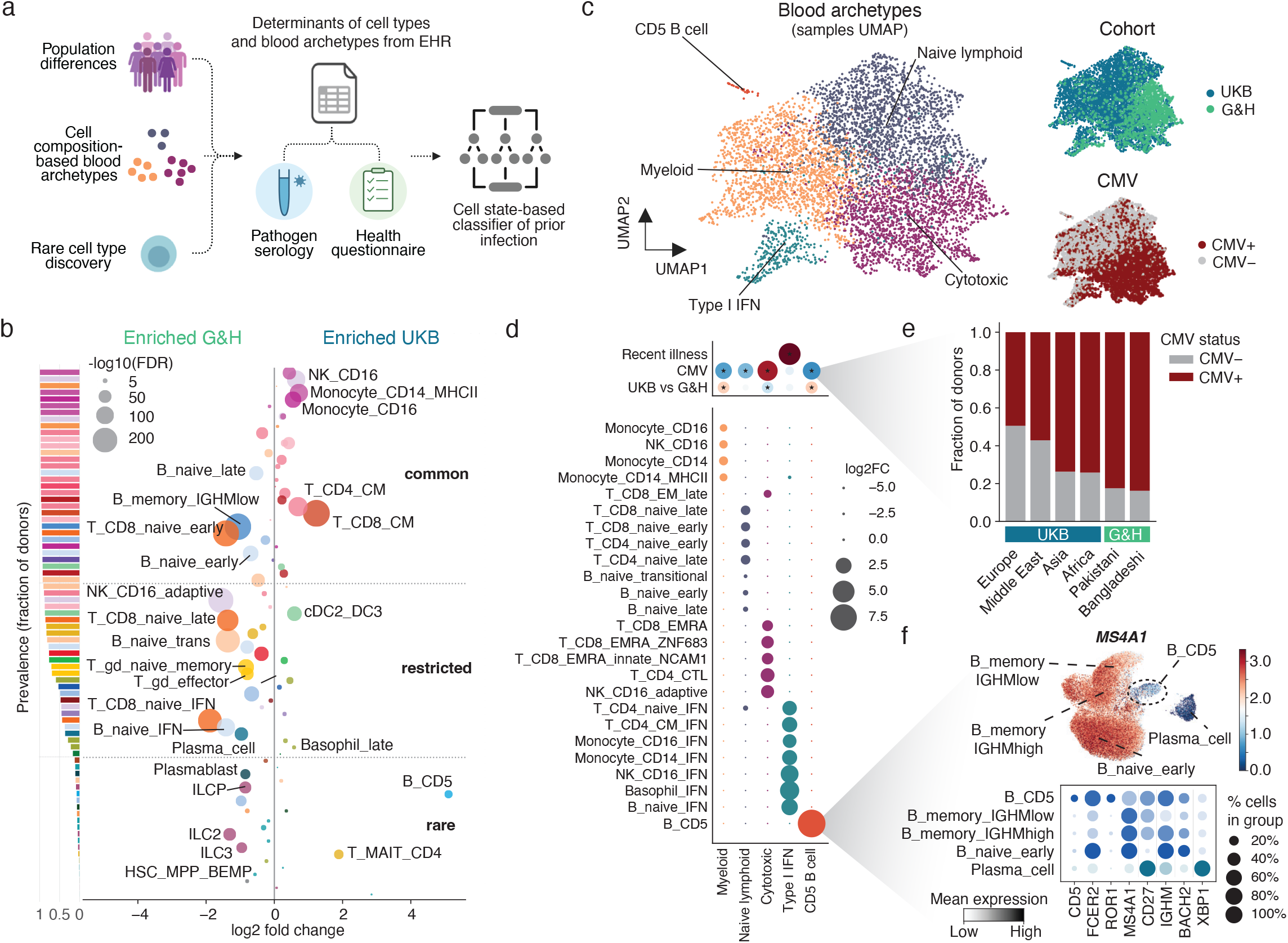
Population structure, infection-associated cell states and predictive single-cell signatures. **a**, Study design linking population-level variation to immune cell composition and infection status. Differences in blood cell composition and rare cell populations were integrated with epidemiological data, including pathogen serology and health questionnaires, to define determinants of cell-type abundance and enable cell state–based classification of prior infection. **b**, Differential abundance of immune cell subsets between cohorts. Scatter plot shows log2 fold change in cell type frequency (CT3 level) comparing Genes & Health and UK Biobank, with point size proportional to significance and labels indicating selected populations. Cell types are ordered based on prevalence across donors, depicted as a bar plot on the left showing fraction of samples with at least 0.1% of a given cell type. Dashed lines denote thresholds for common, restricted and rare populations. **c**, UMAP embedding based on cell type frequencies (CT3 level) of samples of the integrated dataset coloured by blood archetypes (myeloid, naive lymphoid, cytotoxic and type I IFN–responsive, based on the dominant cell types). Insets show samples coloured by cohort (UKB and G&H) and by predicted cytomegalovirus (CMV) serostatus, highlighting infection-associated shifts in cellular distribution. **d**, Association of immune cell states with recent illness, CMV serostatus and cohort. Upper dot plot shows effect sizes (log2 fold change) and significance across blood subtypes with dot size representing significance and colour indicating direction of effect. Lower dot plot shows enrichment of cell types (CT3 level) in the blood archetypes, with dot size representing log2 fold change. **e**, Prediction of CMV serostatus from single-cell data across cohorts. Bar plots show frequencies of predicted CMV serostatus across populations. **f**, Marker gene expression in CD5+ B cells. UMAP shows *MS4A1* (CD20) expression in selected B and plasma cell subsets. Dot plot shows the fraction of cells expressing each gene (dot size) and mean expression level (colour intensity) across selected B and plasma cell subsets, highlighting transcriptional features of CD5+ B cells consistent with monoclonal B cell lymphocytosis.

At the donor level, the companion analysis describes three major groups of donors based on cell state composition termed *blood archetypes*: myeloid, naïve lymphoid and cytotoxic, together with less frequent profiles characterised by coordinated type I interferon (IFN) responses or expanded CD5+ B cell populations^28^ (**Fig. 2c–d**). Their distribution differed between cohorts, with greater representation of the cytotoxic archetype in G&H and of the myeloid and CD5+ B cell archetypes in UKB.

Two examples illustrate the biological information captured by these profiles. CD5+ B cell expansions were more frequent in UKB than in G&H donors (9.5% versus 1.8%; **Fig. 2f)** and remained more frequent in an age-matched comparison (2.3% vs 9.0% of donors of age 55 or older with at least 0.1% CD5+ B cells out of total cells; P = 2.3 × 10^-5^, Fisher’s exact test). Their expression profiles^43^ and skewed immunoglobulin light-chain usage were consistent with monoclonal B cell lymphocytosis, a condition affecting ∼5-12% healthy adults where clonal B cells accumulate in peripheral blood and which, in some cases, progresses to chronic lymphocytic leukaemia (CLL)^44,45^, as examined further in the companion study^28^. The relative paucity of these premalignant states in G&H compared to UKB could be associated with the reported lower CLL incidence in South Asian populations^46–48^.

By contrast, the cytotoxic archetype was associated with cytomegalovirus (CMV) infection, including EMRA CD8+ T cells, cytotoxic CD4+ T cells and adaptive NK cells^17,49^. A cell state-based classifier trained on UKB donors with available serology predicted higher CMV seropositivity in G&H than in UKB (83% versus 50%; **Fig. 2c,e; Extended Data Fig. 2**), aligned with reported seroprevalence^50^ and consistent with a contribution of infection history to the contrasting archetype distributions. The type I IFN archetype was enriched among G&H donors reporting recent infection symptoms, suggesting a coordinated type I IFN signature on circulating immune cells due to an ongoing infection (**Fig. 2d**).

Together, these examples illustrate how the atlas captures immune variation across scales, linking specific cellular states to broader compositional patterns associated with demographic and infection histories. The companion study provides a comprehensive analysis of these organisational patterns, their associated exposures and the clonal processes underlying outlier blood states^28^.

### Demographic, exposure and disease associations across immune composition and cellular programmes

We next illustrate how the atlas, integrated with longitudinal health records and biomedical measurements, captures associations of immune architecture with demographic factors, modifiable exposures and cardiometabolic disease (**Fig. 3a-c**; **Supplementary Table 2; Methods)**. We consider two complementary levels of variation -immune cell abundance and disease-associated cellular programmes-which are explored in depth in companion studies by Zanotti et al.^26^ and Clerici et al.^29^.

**Figure 3:**
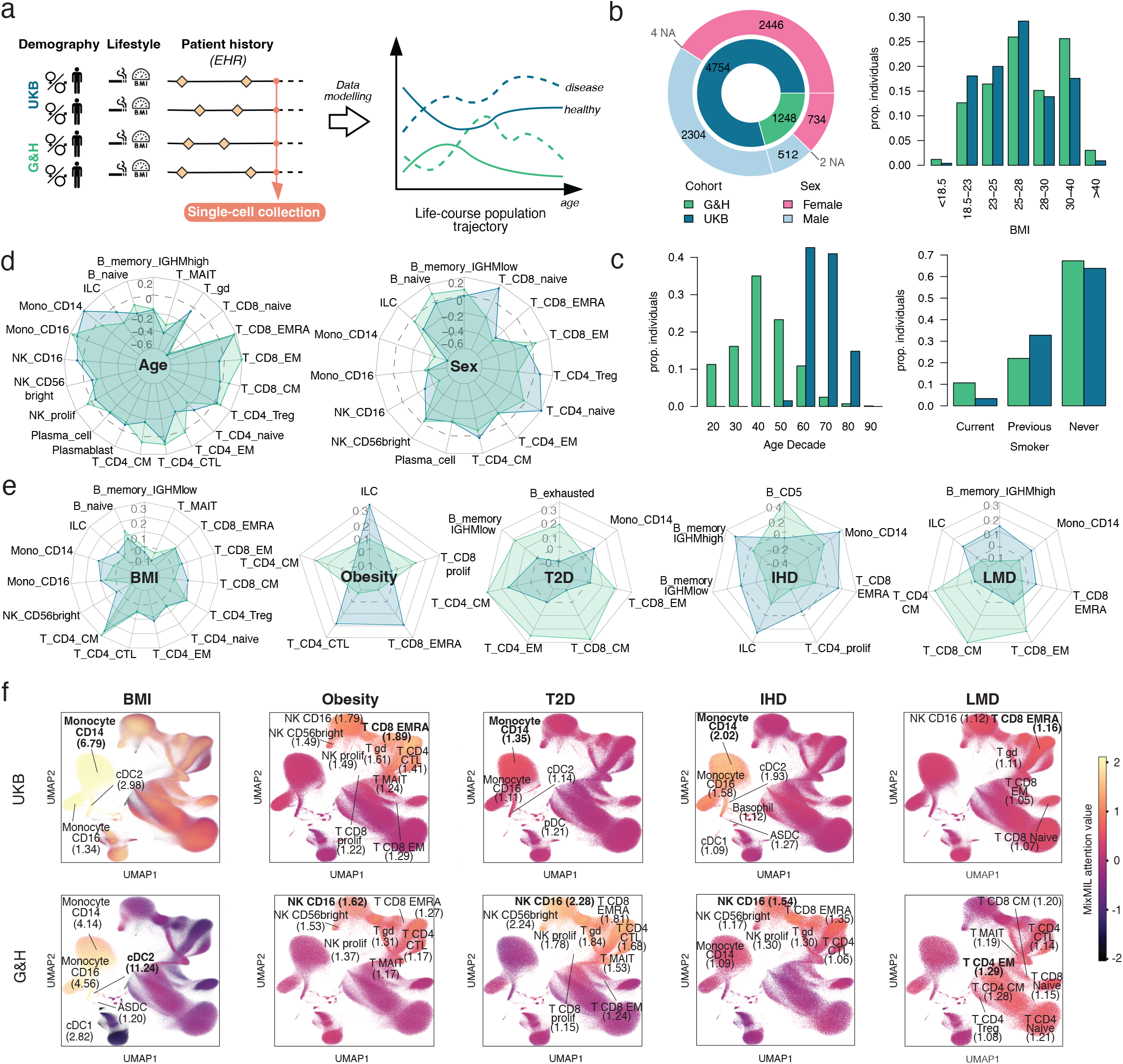
Compositional and gene regulatory programmes from eHR integration with single-cell data. **a**, Conceptual framework for integrating single-cell data with life-course population trajectories inferred from demographic, lifestyle and eHR variables. **b, c** Distribution of demographic (b) and risk factor (c) variables in UKB and G&H cohorts study participants. **d**, Radar plots showing beta coefficients from generalised additive models of associations between scCBC abundance and sex or age in UKB (blue) and G&H (green). Models were adjusted for sex (for age), age (for sex), ancestry, BMI, and smoking status. Only cell types nominally significant in at least one cohort are shown. **e**, Radar plots showing beta coefficients from generalised additive models of associations between scCBC abundance and BMI or disease status in UKB (blue) and G&H (green). Models were adjusted for sex, age, ancestry, BMI (for disease) and smoking status. For each disease, only cell types nominally significant in at least one cohort are shown. The dotted circle indicates a beta coefficient of zero. **f**, UMAP projections of single cells from UKB (n = 4,415) and G&H (n = 844) cohorts. Points are coloured by MixMIL attention weights (pre-softmax, Z-score normalised) for BMI, obesity (n cases: 183 in UKB, 165 in G&H), type 2 diabetes (T2D; n cases: 151 in UKB, 148 in G&H), ischaemic heart disease (IHD; n cases: 237 in UKB, 65 in G&H), and lipoprotein metabolism disorder (LMD; n cases: in UKB, in G&H). Top panels display UKB data; bottom panels display G&H data. Abbreviations: BMI = body mass index; T2D = type 2 diabetes; IHD = ischeamic heart disease; LMD = lipoprotein metabolism disorder.

To examine cell abundance, relative single-cell proportions were combined with matched haematology measurements to estimate absolute counts for 26 cell types and 62 cell states (single-cell complete blood counts, scCBCs; **Supplementary Fig. 6**; **Methods**)^26^. Associations with age, sex, BMI and smoking were broadly concordant between UKB and G&H, with some differences consistent with cohort age distributions and non-linear associations across adulthood (**Fig. 3d, e; Supplementary Table 7**). Analyses of four common cardiometabolic conditions—obesity, type 2 diabetes (T2D), ischaemic heart disease (IHD) and lipid metabolism disorders (LMD)—illustrated greater variation between cohorts in disease-associated compositional signatures than in those associated with demographic factors and modifiable exposures (**Fig. 3e, Supplementary Table 7**), revealing conserved and cohort-specific patterns of immune remodelling characteristic of distinct stages of cardiometabolic disease progression. For example, type 2 diabetes (T2D) was associated with increased central memory CD4+ T cell abundance in both cohorts, whereas ischaemic heart disease (IHD) and lipid metabolism disorders showed predominantly myeloid enrichment in UKB and adaptive lymphoid enrichment in G&H. These examples illustrate shared and cohort-associated features of immune composition explored in detail elsewhere^26^.

Complementing these abundance-based analyses, multiple instance learning (MixMIL)^51,52^ was used to resolve phenotype-associated cellular programmes without relying on predefined cell annotations (**Methods**). The resulting patterns similarly distinguished broadly shared demographic and exposure associations from cohort specific disease-associated signatures (**Fig. 3f; Supplementary Table 8**). For example, BMI was predominantly associated with myeloid programmes, whereas clinical obesity was associated with terminally differentiated cytotoxic programmes across both cohorts^53–56^. The conservation of these patterns suggests that BMI and clinical obesity engage distinct immune pathways, with obesity associated with a broader systemic inflammation response involving a different cellular repertoire. In contrast, we identified divergence in immune programmes associated with disease between UKB and G&H (**Fig. 3f, Supplementary Fig. 7**). T2D and IHD showed greater cohort divergence, with stronger myeloid associations in UKB and cytotoxic lymphoid associations in G&H (**Supplementary Table 8**). Given the increased genetic risk and earlier onset of T2D among individuals of South Asian ancestry ^57^, the adaptive lymphoid remodelling observed in G&H may reflect a distinct immunometabolic trajectory associated with earlier and more prolonged metabolic stress, differences in the timing, duration, or severity of metabolic dysfunction across populations. The companion study examines these disease-associated patterns in greater depth, including age-stratified analyses showing that age accounts only partially for the divergence for selected cardiometabolic phenotypes ^29^.

Overall, these analyses illustrate broadly shared demographic and exposure associations alongside greater cohort variation in disease-associated immune signatures, evident at the levels of both cell abundance and cellular programmes.

### Clinical and genetic influences shape shared and population-specific disease gene regulatory architecture

Having characterised differences in disease-associated immune state architecture between cohorts through compositional and multiple instance learning, we next sought to investigate the genes and transcriptional programmes underlying these patterns.

We first used canonical differential gene expression analyses to identify sets of genes differentially expressed across age, sex, smoking, adiposity and cardiometabolic disease (**Extended Data Fig. 3**; **Supplementary Fig. 8**; **Methods**). Across UKB and G&H, population factors were associated with distinct cell-type-specific transcriptional differences, combining shared genes with cohort-specific patterns. Sex-associated genes included *FAM13A* and *MYEF2*, while age-associated genes included *TSHZ2*, *EDAR, HPGD* and *AUTS2*. BMI associations involved *NFIA, ABCG1 and HPGD* in UKB, and *SOCS3, DUSP2 and JAK3* in G&H. Current smoking showed concordant upregulation of *GPR15* and *LRRN3* across multiple T-cell subsets. T2D-associated genes included *ABCA1, ABCG1* and *KLF10* in both cohorts. IHD signatures differed, highlighting lipid-associated genes in UKB and interferon-response genes, including *ISG15, IFI6 and MX1*, in G&H (**Extended Data Fig. 3**).

Differences in gene expression between cohorts may reflect inherited genetic variation as well as differences in environmental exposures, clinical history and broader cohort context. To distinguish between these factors, we therefore used a more robust statistical framework based on variance decomposition^58,59^ to estimate the relative contributions to gene expression variation of genetic factors versus clinical history and other measured non-genetic factors (**Methods; Supplementary Fig. 9**). To increase power, we analysed pseudobulk gene expression in eight major cell populations (CT1). EHR-derived traits explained significant expression variation beyond that attributable to cis-genetic effects for 1,165 gene-cell-type pairs in UKB (1,034 genes) and 60 pairs (49 genes) in G&H (**Fig. 4a**; **Extended Data Fig. 4a-c; Extended Data Fig. 5; Supplementary Table 9**; **Supplementary Table 10**; **Supplementary Table 11**; **Supplementary Table 12**). Among these gene–cell-type pairs, BMI, smoking and EHR-encoded patient history jointly explained a median of 1.5% (range 0.3-16.1%) in UKB and 4.9% (range 2.6-12.9%) in G&H, compared with 2.8% (0.0-72.7%) and 2.3% (0.0-57.2%) respectively, attributable to cis-genetic variation (**Extended Data Fig. 4d-f**). Although discovery power was substantially greater in UKB, both cohorts confirmed the robust signatures associated with major environmental and cardiometabolic exposures (**Fig. 4a**), including smoking (e.g. *GPR15*^60^*, LRRN3*^60^*, AHRR*^61^) and adiposity-related metabolic dysregulation (e.g. *ABCG1*^62^*, CLU*^63^) (**Fig. 4b-e**, **Extended Data Fig. 6a-c**). Cohort-specific signatures associated with smoking and BMI were also evident, involving genes such as *LINC000599*, *LGALS1*, *FAM241B* and *PDE3B* (**Extended Data Fig. 6d-g**). To assess whether the observed associations with EHR variables were consistent with a causal effect on gene expression, we performed Mendelian randomisation (MR) for gene–cell-type pairs significantly associated with BMI in UKB (**Methods**). We found strong directional concordance between variance decomposition and MR analyses, with 90% of nominally significant gene–cell type pairs (p < 0.05) showing concordant directions of effect and 19 genes reaching experiment-wide significance in the MR analysis (FDR < 0.05 and Q-stat > 0.05; 89% with directionally concordant effects; **Extended Data Fig. 7**; **Supplementary Table 13**). This supports a causal contribution of adiposity to the perturbation of immune-cell transcriptional states as the main driver in this relationship. More broadly, this experiment demonstrates the value of single-cell data integrated with EHRs to explore how exposures and disease diagnoses influence gene expression in specific cell types.

**Figure 4:**
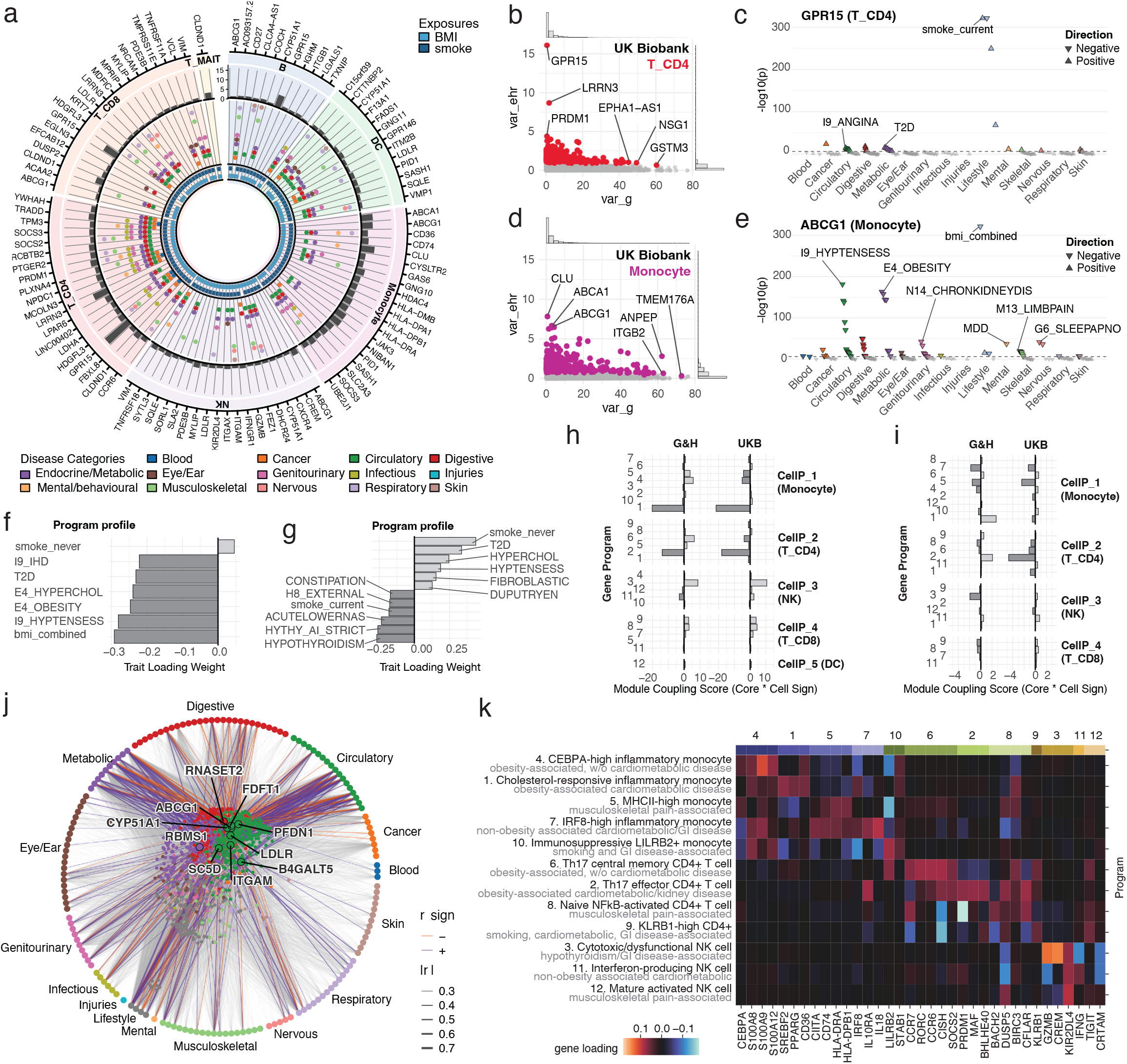
Exposure-driven gene expression and latent programs architecture. **a**, Top associations identified by variance decomposition between single-cell gene expression and eHR-derived traits in UKB. Shown are the 20 genes with the largest eHR-explained variance (Pearson’s r *≥* 0.2). The outer ring indicates the fraction of gene expression variance explained by eHR variables, and inner rings denote significant disease and lifestyle-trait associations. **b**, **c**, Smoking-associated signatures. **b**, Variance explained by eHR (var_ehr) versus cis-genetic effects (var_g) for genes tested in CD4+ T cells in UKB. Highlighted genes show the strongest environmental or genetic contributions. **c**, Phenome-wide association profile of *GPR15* in T_CD4 cells. **d**, **e**, Adiposity-associated signatures. **d**, Variance explained by eHR versus cis-genetic effects in monocytes. **e**, Phenome-wide association profile of *ABCG1*. **f**, **g**, Trait loadings from Tucker decomposition for Trait Program 1 (cardiometabolic traits) (f) and Trait Program 4 (smoking-related traits) (g). **h**, **i**, Coupling scores between trait and gene programmes across cell types for Trait Program 1 (h) and Trait Program 4 (i). **j**, Associations between the top 10 genes in Gene Program 1 and all tested traits. Line colour denotes the direction of association and line width is proportional to Pearson’s r. Gene colours indicate the predominant associated trait category. **k**, Gene loadings across gene programmes. Representative marker genes were selected from the top positive loadings of each programme based on biological relevance. Gene programmes are annotated by dominant functional categories and ordered according to the cell types showing highest programme activity. Grey annotations indicate major trait-program associations.

We next asked whether patterns of EHR-associated gene expression across genes, cell types, traits and cohorts could be used to define coordinated transcriptional responses to environmental and clinical exposures. First, using variance decomposition, we computed the expression component explained by medical history across celltype-gene-traits combinations and we performed hierarchical clustering on gene by trait correlation matrices (Pearson’s r) across cell types, focusing on genes with the highest variance explained (**Supplementary Fig. 10**; **Supplementary Fig. 11**). Then, using Tucker decomposition^64^, we modelled the single-cell transcriptomic correlations between gene expression and EHR traits into a multidimensional tensor to capture complex cohort-specific effects across all the dimensions. Based on ranks optimisation, we identified twelve gene programs, six cell-type programs, and six trait programs, a final configuration that collectively describes the organisation of immune responses to systemic exposures across both UKB and G&H (**Methods**; **Supplementary Fig. 12**; **Supplementary Table 14**; **Supplementary Table 15**; **Supplementary Table 16**). Trait programmes capture distinct phenotypic clusters with lineage-specific activation, while gene programmes reflect discrete biological functions (**Supplementary Fig. 12**).

Metabolic trait programmes (Trait Programme 1 and 3) were strongly coupled to monocyte-centred gene modules, especially to gene program 1 enriched for cholesterol homeostasis (normalised enrichment score [NES]=1.88, adjusted p=1.75×10^−3^), steroid metabolism (NES=2.01, adjusted p=3.91×10^-4^), and *MTORC1* signalling (NES=1.99, adjusted p=2.31×10^-5^), whereas inflammatory and infectious trait programmes (Trait Programme 6) were associated with broader immune-cell repertoires, enrichment in anti-viral response (gene program 2, viral process: NES=1.83, adjusted p=1.12×10^-2^; significant enrichments in multiple viral response activation signatures from ImmuneSigDB), and suggestive enrichment of *IL2-STAT5* (gene program 2, NES=1.51, p=5.53×10^-2^), *IL6-JAK-STAT3* (gene program 3, NES=1.56, p=2.84×10^-2^) pathways (**Supplementary Fig. 13**; **Supplementary Table 17**; **Supplementary Table 18**; **Supplementary Table 19**; **Supplementary Table 20**). Monocytes consistently dominated metabolic and inflammatory trait programs, whereas infectious/allergic programs involved broader immune participation, including dendritic and NK cells (**Extended Data Fig. 8**).

Many programme interactions were conserved across cohorts, indicating shared biological responses to common exposures. For instance, trait programme 1, including obesity, hypertension, and T2D (**Fig. 4f**) showed highly concordant coupling across cohorts with gene programme 1 and 2 (**Fig. 4h**) in monocytes and CD4^+^ T cells (*PDE3B*, *LDLR, ABCG1;* **Fig. 4j**). Further, hypercholesterolemia and cardiometabolic disease coupled with cholesterol-responsive inflammatory monocyte gene programs (*SREBF2, PPARG, CD36*) and Th17 effector T-cell states across both cohorts. Other couplings exhibited marked activation differences in the two cohorts (**Extended Data Fig. 8**), particularly for smoking-related immune responses (Gene Programme 2 and 3; **Fig. 4g, i**; **Extended Data Fig. 9**), and cardiometabolic traits (Gene Programme 4 and 5; **Fig. 4h**). Consistent with these programme-level results, differential expression analyses highlighted predominantly monocyte-driven dysregulation in UKB (*ABCA1* and *ABCG1*) and stronger T-cell-associated signatures in G&H involving interferon-responsive genes such as *ISG15*, *IFI16*, and *IFI44* (**Extended Data Fig. 3**). These findings suggest that single cell genomics is able to capture signatures of the different stages of cardiometabolic immune remodelling, potentially representative of obesity-associated inflammation, activation of cholesterol-responsive innate programmes and adaptive effector-cell activation during cardiometabolic disease progression. The cellular deployment of these programmes in the two cohorts may be shaped by demographic structure (e.g. age) as well as environmental context (e.g. CMV infection or obesity rates; **Fig. 4k**).

Together, these analyses demonstrate that integration of single-cell transcriptomics with longitudinal health records enables high-resolution mapping of gene regulatory programmes associated with environmental exposures and disease. While cardiometabolic conditions engage conserved transcriptional programmes across populations, the cellular contexts in which these programmes are deployed differ substantially, revealing cohort-specific immune architectures underlying common disease processes.

### Population-specific regulatory genetic variation reflects both genetic architecture and cohort context

Genetic variation influences immune cell identity and function, whereas lifetime exposures can reshape immune composition and transcriptional states, thereby altering the contexts in which regulatory variants act. We therefore tested whether genetic regulatory variation exhibits cohort-specific effects associated with clinical and exposure-related differences captured by EHR, beyond ancestry-related differences in allele frequency. We systematically mapped cis-regulatory expression quantitative trait loci (cis-eQTLs) associated with pseudobulk gene expression across 23 main immune cell types in CT2^27^. Using the FLANDERS framework ^27^ we determined 41,691 independent causal genetic variants (eSNPs) in 4,452 European-ancestry UKB donors (starting from 114,063 fine-mapped eQTL signals) and 17,274 eSNPs in 729 British Pakistani- and 499 British Bangladeshi-ancestry G&H donors (starting from 45,819 fine-mapped eQTL signals) **(Extended Data Fig. 10a-c; Methods)**. Overall, we identified 15,419 significant eGenes in the UKB and 10,339 in the G&H cohort (**Extended Data Fig. 10e**, **Supplementary Table 21**). Discovery power increased with donor number, cell abundance and the proportion of donors expressing each gene, yielding more associations in myeloid than lymphoid populations (**Supplementary Fig. 14**; see the data availability section to access complete eQTL and fine-mapping results). As expected, most signals mapped to non-coding regions near the transcription start site (TSS) of the associated gene, without directly altering its protein-coding sequence (**Supplementary Fig. 15).** FLANDERS classified eSNPs into four categories according to their effects across genes and cell types, revealing a highly structured pattern of regulatory pleiotropy: approximately 60% affected a single gene in a single cell type (specific), and 25% affected a single gene across multiple cell types (multi-context), whereas effects on multiple genes within a single cell type (multi-gene) or across multiple cell types (highly pleiotropic) were less frequent (**Extended Data Fig. 10f-g**, **Supplementary Table 22**). The genetic architecture of gene expression control across cell types and its relation to disease is explored in detail in a companion paper by Pirastu et al ^27^.

To quantify shared versus potential population-specific regulation, we further performed systematic cross-cohort fine-mapping and colocalisation, restricting comparisons to matched cell type–gene pairs (**Methods**). This analysis identified 28,314 fine-mapped credible sets shared between the two cohorts, spanning 26,653 gene–cell type pairs and 7,193 unique eGenes across 19 immune cell types (**Fig. 5a–c**). Shared genetic signals exhibited highly correlated effect sizes between cohorts (r = 0.97; **Supplementary Fig. 16**), indicating largely conserved genetic regulation of immune gene expression. After excluding 2,670 and 38,553 loci (G&H and UKB, respectively) insufficiently resolved because of differences in sample size, allele frequency, and statistical power, we identified 5,582 UKB-specific credible sets (3,727 unique eGenes, 4.89% of UKB eQTLs) and 6,878 G&H-specific credible sets (3,888 unique eGenes, 15.01% of G&H eQTLs) (**Fig. 5a, Supplementary Table 23; Supplementary Table 24; Methods**). Together, these findings reveal extensive sharing of regulatory effects across populations, alongside a subset of population-specific regulatory variants (**Fig. 5c**).

**Figure 5:**
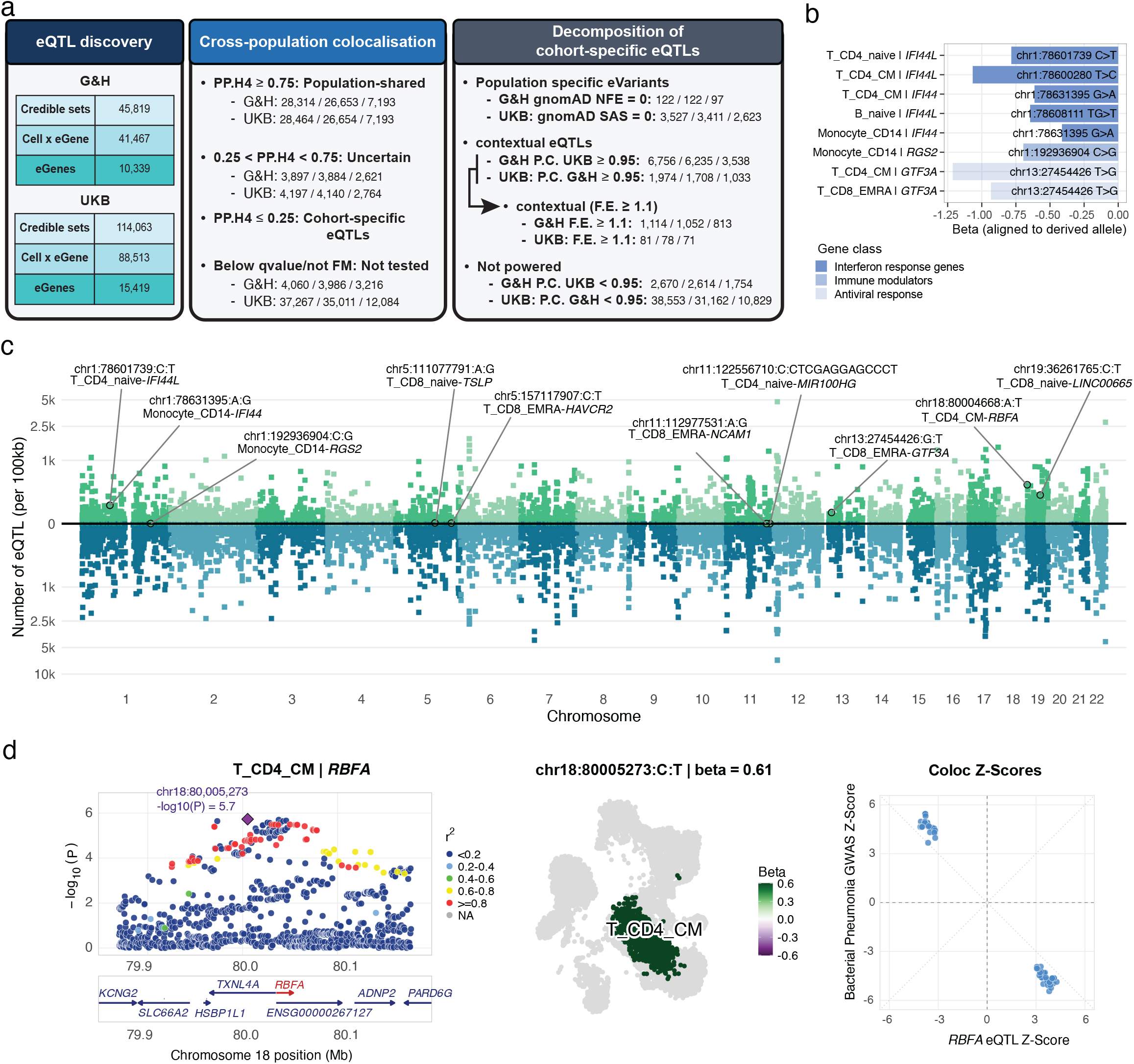
Allelic differences in gene regulatory architecture between populations. **a**, Diagram illustrating the overall eQTL discovery in the 2 cohorts, the criteria used for the selection of cohort-specific signals and the number of resulting population-specific eVariants and contextual eQTLs. Triplet numerical values represent mapping to: Credible Sets / Cell Type x eGene Intersections / Unique Distinct eGenes. **b**, G&H eVariant-associated eGenes with annotated roles in the interferon response, with effect sizes aligned to the transition from ancestral to derived alleles. **c**, Brisbane plot showing the density of eQTL fine-mapped signals in the G&H (upper part) and UKB (lower part) cohorts. Interesting examples of variants showing a G&H population-specific effect are highlighted. **d**, A bacterial pneumonia risk locus (chr18:80004668:A:T) colocalising with an eQTL for *RBFA* (chr18:80005273:C:T) in T CD4 CM cells. The inverse relationship between disease risk and *RBFA* expression is shown using z-scores. Abbreviations: FM = Fine-mapped, P.C. = Power Calculation, FrEXP = Fraction of Expression

We next examined cohort-specific eQTLs to determine whether they reflected ancestry-related genetic differences between the two cohorts or differences in cohort context. A subset of cohort-specific eQTLs could be explained by marked differences in allele frequency between European and South Asian ancestry, as assessed using independent reference cohorts ^65^. These included 122 G&H-specific variants and 3,527 UKB-specific variants (**Fig. 5a-c**). Among the G&H-specific variants, several affected genes involved in interferon-mediated immunity, including *IFI44* ^66^*, IFI44L* ^67^, *RGS2*^68,69^, and *GTF3A*^70^, with the non-ancestral allele consistently associated with reduced expression (**Fig. 5b**). This suggests population-specific modulation of interferon signaling, a pathway known to be under strong evolutionary constraint and recurrent pathogen-driven selection differentiating South Asian from European ancestries^71^. Another interesting example was 18:80005273:C:T, rs115449055, a variant that is rare in Europeans (MAF < 0.01%) but more common in South Asians (MAF > 3%), colocalising with increased bacterial pneumonia risk and reduced *RBFA* expression in CM CD4⁺ T cells (**Fig. 5d**). As *RBFA* regulates mitochondrial ribosome biogenesis ^72^, reduced expression may impair the metabolic fitness of CM CD4⁺ T cells during infection, highlighting the potential for functional diversification due to genetic variation acting in ancestrally divergent cohorts. Additional examples are given in **Supplementary Table 23**; **Supplementary Table 24**.

Strikingly, we also identified a second class of eQTLs with comparable allele frequencies and sufficient predicted power for detection in both cohorts, yet observed exclusively in G&H or UKB (G&H- and UKB-contextual eQTLs; **Fig. 5a; Supplementary Fig. 16; Methods**). Because eQTL discovery depends on sample size with measurable gene expression, we hypothesised that these signals reflected cohort differences in the prevalence of target-gene expression. Accounting for allele frequency and statistical power therefore prioritised signals whose population specificity may reflect context-dependent gene regulation. In support of this hypothesis, for a subset of these contextual eQTLs we identified a positive skew in the ratio of donors expressing the gene in G&H compared to UKB (contextual eQTLs with fraction of expression [F.E.] ≥ 1.1, **Methods**; **Fig. 5a, Supplementary Fig. 16, Supplementary Table 24**).

Gene Ontology (GO) enrichment analysis revealed no substantial differences in immune-related functions between cohort-specific and cohort-shared eQTLs (**Fig. 6a**). However, G&H-contextual (F.E ≥ 1.1) eQTLs were enriched in genes upregulated in conditions more prevalent in this cohort (**Fig. 6b**), and were associated with the trait- and cell-type-specific programmes showing the greatest divergence between G&H and UKB (**Fig. 6c**). These findings suggest that cohort differences in phenotypic exposures or disease prevalence may contribute to context-dependent gene regulation. Both G&H- and UKB-contextual (F.E ≥ 1.1) eQTLs were also enriched for long non-coding RNAs (lncRNAs; **Fig. 6d**), consistent with their emerging role in context-dependent immune and environmental responses^73^. Relative to shared eQTLs, G&H-contextual (F.E ≥ 1.1) eQTLs were additionally enriched in subsets of CD8⁺ T cells and B cells (**Supplementary Fig. 17a**). Notable examples included a naïve CD4 T cell eQTL (rs1862418475-G) for *MIR100HG* implicated in periodontitis^74^, and a naïve CD8⁺ T cell eQTL (rs73040148-T) for *LINC00665* (**Supplementary Fig. 17b–c**), whose downregulation improves cell viability in LPS-induced inflammation^75^. Among eQTLs affecting protein-coding genes, we identified 11:112977531:A:G (rs1940707) and 5:157117907:C:T (rs10057431) in EMRA CD8^+^ T cells, associated to *NCAM1* and *HAVCR2* genes, respectively, which are linked to the acquisition of NK-like and exhaustion phenotypes in EMRA CD8^+^ T cells under chronic CMV exposure^76^ (**Fig. 6e** and **Supplementary Fig. 17d**). Some G&H contextual (F.E ≥ 1.1) eQTLs appeared to reflect changes in immune cell composition associated with disease states or predicted viral exposure. For example, an eQTL regulating *TSLP* expression in naïve CD8⁺ T cells was associated with atopic dermatitis ^77^, a condition more prevalent in G&H (odds ratio = 3.59; FDR 5%, χ² test), and associated with expansion of this cell population **(Supplementary Fig. 17e–f).**

**Figure 6:**
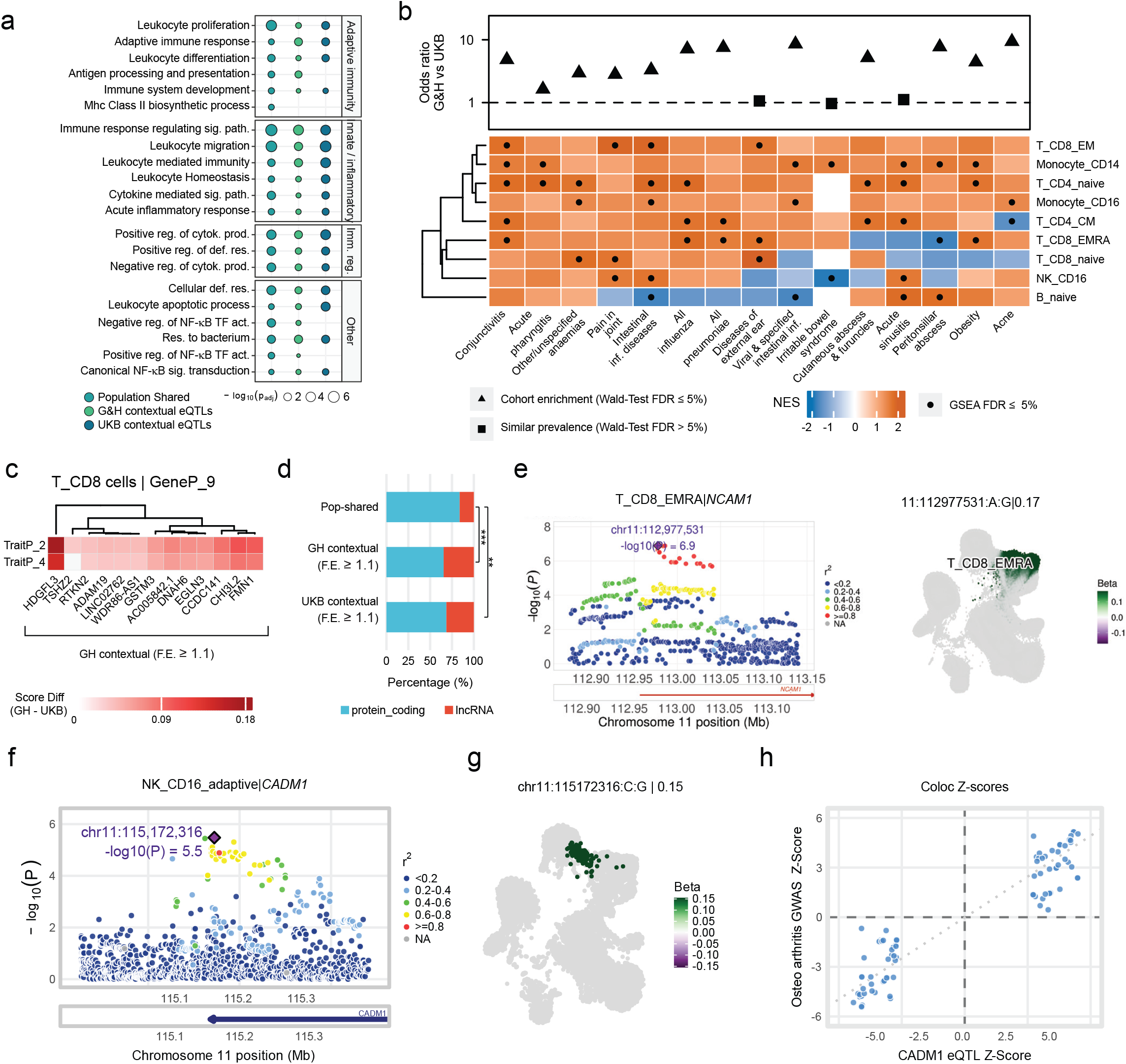
Context dependent genetic effects on gene expression and disease. **a**, Overrepresentation analysis of population-shared, G&H, and UKB contextual eQTLs for Gene Ontology (GO) Biological Process terms related to immune function. b, Heatmap showing normalised enrichment scores (NES) from cell type–matched gene set enrichment analysis (GSEA) of G&H contextual eQTLs F.E. *≥* 1.1 against differentially expressed genes resulting from various conditions. The corresponding relative odds ratio (G&H versus UKB) for each condition is displayed above the heatmap. **c**, Heatmap showing the GSEA of genes within the top 5% most divergent final scores (difference in G&H and UKB for the product of coupling score x gene loading) against G&H contextual eQTLs F.E. *≥* 1.1. **d**, G&H and UKB contextual eQTLs F.E. *≥* 1.1 eGenes are significantly enriched for long non-coding RNAs (lncRNAs) compared with population-shared eQTLs (** *<* 0.01, *** *<* 0.001, Fisher’s exact test). **e**, LocusZoom plot (left) and UMAP projection (right) illustrating the eQTL 11:112977531:A:G (rs1940707) associated with *NCAM1* expression in EMRA CD8+ T cells. **f**, **g**, LocusZoom plots (left) and corresponding effect size plots (right) for the eQTL 11:115172316:C:G (rs4938182) associated with *CADM1* expression in adaptive NK cells. **h**, Direction of effect for the significant Mendelian randomization association between *CADM1* expression and osteoarthritis risk, indicating that higher *CADM1* expression is associated with an increased risk of disease. Abbreviations: sig. pat. = signaling pathway, cytok. prod. = cytokine production, def. res. = defense response, Res. = response, reg. = regulation, TF act.= transcription factor activity, sig. = signal. Score Diff. = absolute difference in final score for G&H and UKB cohorts.

To further investigate whether chronic viral exposure may reveal context-dependent genetic regulation, we focused on adaptive NK cells, a CT3 cell state associated with CMV seropositivity ^78^. Mapping cis-eQTLs in UKB participants carriers of this cell state (40% of the cohort) identified 358 signals not detected in other PBMC populations or non-blood GTEx tissues, suggesting regulatory effects specific to this cellular context (**Supplementary Table 25**; **Methods**). Colocalisation with disease and trait GWAS loci followed by Mendelian randomisation identified 33 adaptive NK cell-specific signals with evidence of causal effects on disease not shared with other PBMC populations or GTEx tissues (**Supplementary Fig. 18a**; **Supplementary Table 26; Methods**). One notable example was an eQTL (11:115172316:C:G, rs4938182) regulating *CADM1* expression, which colocalised with osteoarthritis risk in both UKB and G&H cohorts. MR supported a causal association between increased *CADM1* expression and osteoarthritis risk **(Fig. 6f–h, Supplementary Fig. 18a)**. *CADM1* is not typically expressed in lymphocytes, but mediates cell adhesion through homophilic interactions, or heterophilic binding to *CRTAM*, *CADM3*, or *CADM4* on opposing cells, and has been implicated in immune synapse formation^79^. Its expression in adaptive NK cells is consistent with the increased chromatin accessibility at the *CADM1* locus in adaptive NK cells during CMV infection^80^. These results suggest a potential link between CMV-driven NK-cell adaptation and osteoarthritis susceptibility. Additional eQTLs specific to adaptive NK cells implicated *BID* in increased monocyte counts, and *FOXO3* in elevated circulating triglyceride levels (**Supplementary Fig. 18b-c**). We also identified disease-associated loci that colocalised with eQTLs restricted to adaptive NK cells among immune cells -but also detected in GTEx tissues-including *TRAF1* for rheumatoid arthritis, and *CXCR1* for knee osteoarthritis (**Supplementary Fig. 18d-e**).

Together, these findings support a model in which disease prevalence, immune-cell composition and transcriptional regulation interact to generate cohort-specific regulatory effects from largely shared genetic variation. More broadly, they suggest that integrating non-genetic determinants of immune-cell composition with genetic analyses is essential for understanding population differences in the regulatory architecture of complex traits.

## Discussion

The human immune system operates at the intersection of genetic and environmental factors, continuously adapting to lifetime exposures and influencing health and disease. Our study provides a population-scale view of how ancestry, environmental exposures, and disease jointly shape the architecture of human immunity^5,8,9^. By integrating single-cell transcriptomics with longitudinal health records and genetic data across ethnically distinct cohorts, we show that immune variation emerges from dynamic interactions between inherited genetic background, environmental exposures, and disease status to shape human health.

A striking observation was the asymmetry between conserved exposure-associated programmes and population-divergent disease signatures. Ageing, smoking, BMI, and biological sex produced highly reproducible immune programmes, suggesting the existence of shared axes of immune remodelling. In contrast, chronic cardiometabolic diseases engaged substantially different cellular architectures in the two cohorts, despite overlapping clinical phenotypes. These findings suggest that common diseases may converge clinically while arising through partially distinct immune trajectories shaped by population-specific exposure histories, environmental contexts and genetics.

Our dataset integrates population-scale single-cell transcriptomics, genetic information and detailed phenotyping from electronic health records across two cohorts of distinct ancestry. This design enables robust modelling of how genetic and non-genetic influences shape immune programmes and their contribution to disease. These analyses suggest that cardiometabolic disease progression is accompanied by staged non-genetic immune remodelling^81,82^. Early adiposity-associated states were characterised predominantly by inflammatory monocyte and dendritic-cell programmes, whereas obesity was associated with broader activation of cytotoxic and innate-like lymphocyte compartments ^53–56^. Established cardiometabolic disease states additionally engaged cholesterol-responsive monocyte programmes and differentiated effector adaptive immune states. Together, these observations support a model in which metabolic dysfunction progressively reorganises immune ecosystems from early innate inflammatory sensing toward chronic adaptive and cytotoxic remodelling.

A second major finding is that population-specific regulatory effects frequently arise despite highly conserved underlying genetic architecture. Although most cis-regulatory effects were shared across populations, a substantial subset of eQTLs emerged preferentially within one cohort in association with differences in immune composition, exposure prevalence, and disease-associated transcriptional states. These findings suggest that regulatory variant activity is conditioned not only by cell type identity, but also by environmentally induced immune states ^10,83^. In this framework, genetic effects become functionally active only within specific cellular and physiological contexts generated through exposure-dependent immune remodelling.

Chronic viral infection emerged as a particularly strong determinant of immune state organisation. CMV-associated remodelling profoundly altered cytotoxic T-cell and adaptive NK-cell compartments^17,49^ and was associated with distinct regulatory effects detectable preferentially within these exposure-associated states. These observations suggest that single-cell genomics is uniquely positioned to capture exposure-associated changes in immune-cell composition, and raise the possibility that persistent environmental exposures establish latent regulatory architectures that remain undetectable in conventional population studies lacking cellular resolution or exposure stratification.

Our multiethnic design allowed us to explore cohort-specific differences in both compositional traits and gene regulatory programmes that differentiate donors of European ancestry compared to South Asian ancestry living in the UK. For instance, we identified differentially enriched rare cell types, including CD5+ B cells drastically expanded in a small subset of donors restricted to UKB, representing monoclonal B cell lymphocytosis, a precursor stage to CLL^44,84^. This difference in prevalence of an ageing-associated premalignant cell state could explain the reported lower CLL incidence in Asian populations^46^, including the lower prevalence of CLL in British Pakistani/British Bangladeshi in UK^85^, and informs genetic mechanisms underlying the emergence of this state in ethnically stratified individuals.

More broadly, our findings suggest that population differences in immune-mediated disease biology may often reflect differences in immune system state shaped by environmental and/or demographic factors rather than genetic divergence. Populations differ in infection burden, metabolic exposure, socioeconomic environment, and disease prevalence, all of which can reshape immune composition and transcriptional state ^8,86–88^. In turn, these changes alter the cellular environments within which regulatory variants operate. This model may help explain why genetic associations often display variable penetrance and transferability across populations despite broadly shared genetic architecture.

Several design aspects of this study should be acknowledged. Because ancestry, environment, and disease prevalence are partially confounded in observational cohorts, the relative contribution of these factors cannot be completely disentangled in this setting alone. In addition, our analyses are cross-sectional and therefore cannot resolve temporal relationships between immune remodelling and disease progression. Well-controlled longitudinal and perturbational studies will be required to determine which of the identified immune states directly contribute to disease pathogenesis or instead reflect downstream consequences of chronic disease and exposure.

Overall, our results support a framework in which immune states represent dynamic interfaces linking environmental exposure, population history, and genetic regulation across the human life course. Population-scale single-cell atlases therefore provide not only descriptive catalogues of cellular diversity, but also a means to define how environmental and genetic forces intertwine to generate population-specific disease biology. This work establishes a foundation and a resource for the community for studying human disease through the joint analysis of immune state architecture, environmental exposure, and regulatory genetics across diverse populations.

## Methods

### Ethics

Recruitment descriptions: UK Biobank obtained approval from the North West Multi-centre Research Ethics Committee (MREC) UK NHS REC approval reference number: 21/NW/0157. Genes & Health obtained ethical approval from London South East NRES Committee of the Health Research Authority approval reference number: 14/LO/1240.

### Sample collection

#### Collection

UK Biobank samples were collected in accordance with protocol previously described^89^. East London Genes and Health cohort is described in previous publication^31^. Donors attended clinics at three UK Biobank imaging centres: Stockport, Newcastle and Reading and two clinics for Genes and Health: London and Bradford. Blood was collected by venipuncture into 2ml EDTA spray coated vacutainer for Genes and Health (Greiner Vacuette #454087) or 3ml EDTA for UK Biobank (Becton Dickinson #BD367838). Samples were kept refrigerated until transport. Details of the demographic of the two collections are given in **Supplementary Table 1.**

#### Shipment

Samples were transferred at the end of clinic day to a pre-chilled Credo CubeTM reusable container. These were then couriered overnight to a central consolidation facility before forwarding onto the Wellcome Sanger Institute (Cambridge, UK) for processing the following morning. The large majority of samples (98.9%) were first processed within 24 hours of venepuncture. The number of donor samples received from each site was; 1,921 from Stockport, 1,390 from Reading, 1,813 Newcastle, 1,204 London (with 24 donors providing a repeat measurement ∼7 months apart) and 311 from Bradford. See **Supplementary Table 3**.

### Single-cell sequencing

#### PBMC Extraction

Working in a Class II biological safety cabinet 150µl of blood for each donor was transferred to a 96-well-semi-deep well plate (Thermo Fisher #249944). Donor positions within the arrayed plate were stored within laboratory information management systems (LIMs). All liquid and cell handling steps from this point on were carried out on a custom Hamilton Star liquid handler (Hamilton Robotics). The PBMC extraction was performed using reagents and a modified protocol from Stem Cell Technologies PBMC Easy Sep Direct kit (#19654). In brief; blood array plate was transferred to a chilled station on the PBMC Star deck. Antibody and magnetic beads were arrayed in Eppendorf Twin Tec full skirted plates and placed on a chilled station. 15µl of EasySep direct antibody was added to blood on a chilled station and incubated for 5 minutes. 200µl of EasySep Buffer (Stem Cell Technologies #20144) was added followed by 15µl of Rapid Sphere magnetic beads with subsequent gentle mixing by repeat aspiration. Each plate was transferred to the magnetic station (Alpaqua Magnum FLX) and held for 5 minutes. 250µl of supernatant containing purified PBMC was transferred to a fresh semi deep well plate and a further 15µl of Rapid Sphere was added. This was followed by a further two 5 minute magnet incubation and supernatant aspiration steps to ensure removal of all bead / red blood cell complexes. Cells were then centrifuged (500rcf) and washed twice in PBS/BSA 0.04% and re-suspended in a final volume of 75µl.

#### Assay Controls

Over the course of the experiment we included two immortalised monocyte-like myeloid leukaemic cell lines (THP-1 and U937), as monitoring controls. These were obtained from the European Collection of Authenticated Cell Cultures (ECACC) and validated free of Mycoplasma contamination. They were passaged according to supplier recommended conditions in RPMI 1640 (Thermo Fisher #61870010), 10% FBS (Thermo Fisher #10500056) and 1% Penicillin-Streptomycin (Thermo Fisher #15070063). A single batch of each line was expanded to desired volume, then re-suspended in Recovery freezing media (Thermo Fisher #12648010) then split into individual aliquots. These were then placed in controlled rate freezing containers in -80°C, before transfer to liquid nitrogen until required. In total 24 replicates of THP-1 and 25 replicates of U937 were included over the duration of the experiment. Frozen stocks were retrieved from liquid nitrogen, rapidly defrosted and washed in pre-warmed RPMI 1640 (Thermo Fisher #61870010) with 10% FBS (Thermo Fisher #10500056) as described in 10x Genomics protocol (CG00039). Dead cell removal was carried out using a Dead Cell Removal kit (Miltenyi Biotech #130-090-101) following manufacturer’s instructions. Cell lines were added to a reserved well on the extracted PBMC plate and included in the process from here on.

#### Cell counting

To determine cell viability and concentration we used a Nexcelom Cellaca MX high throughput cell counter (Revvity). Haemocytometer chambers (Nexcelom #CHM24-B100) were prepared using the Hamilton Star. A 1 in 4 dilution consisting of 25µl PMBC, 25µl PBS/0.04%BSA and 50µl Acridine Orange/Propidium Iodide (Nexcelom #CS2-0106) was prepared in a separate Eppendorf 200µl TwinTec plate. For all extractions, median PBMC concentration was 1.22×10^6^ and 1.00×10^6^ for PBMC extracted after 24 and 48 hours respectively. Only one cell count per donor was obtained and this counts file was uploaded into LIMs. Median viability for PBMC was 98% (50-100%) and 99% (83-100%) for PBMC extracted 24 and 48 hours after venipuncture respectively. Median viability for cryopreserved PBMC was 74% (5-100%). In general PBMC that were below 50% viability were not included in subsequent 10x reactions.

#### Pooling

Samples were sequenced in 634 pools composed of 9 to 12 samples. In order to resolve potential centre batch effects we implemented donor randomisation with a rule that each clinic was equally represented across all 10x reactions performed on a given day. LIMs scripts would calculate the pipetting volume required for each destination well to receive 20,000 cells per donor. Cells were then centrifuged and concentrated to achieve a concentration of ∼6,400 cells per microlitre, and target a capture of 25,000 cells per channel.

#### Single Cell RNA library preparation

Single-cell encapsulation, cDNA and Illumina library preparation took place on a 10x Genomics Chromium Connect using Single Cell 3’ Library and Gel Bead Kit (10x Genomics #1000141) and Automated Chip G (10x Genomics #1000136). The majority of libraries were prepared using dual index kit (#1000213), however there were a small minority prepared with single index (#1000212). 12 PCR cycles of first round cDNA amplification and 10 PCR cycles of Illumina index amplification was performed for all reactions.

#### Sequencing

Bioanalyzer High Sensitivity assay (Agilent #5067-4626) was used to determine the library concentration. Weighted pools for sequencing were constructed based on predicted cell capture and desired mean read per cell coverage. These were then diluted to 2.8nM prior to loading. Sequencing was performed on NovaSeq 6000 (Illumina) with an average 113 donors and 200,000 cells run across one S4 flow cell and sequenced across 62 runs with parameters; 2⨉150 cycle flow cells (26bp (Read 1), 8bp (Index), and 98 bp (Read 2)), generating a total of approximately 620 billion reads.

### Sysmex CBC measurement

A complete blood count was performed from the EDTA blood sample using a Sysmex XN-1000 haematology analyser to generate an extended profile of blood cell indices. Once whole blood aliquots for PBMC extraction were made, vacutainers were loaded into Sysmex and run at ambient temperature. We assayed 90.3% of the collected cohort, of which 98.7% were run on the Sysmex within 24 hours of venepuncture. Samples were then returned to 4 degrees centigrade where they were kept for up to three days before disposal. Symex was externally benchmarked using an independent external quality assurance sample for duration of study.

### Cryopreservation

PBMC extraction for the cryopreservation protocol was the same as before with the main difference that 2×150µl blood aliquots per donor were made and consolidated into a single cryovial. Once PBMC extraction was complete cells were centrifuged down and re-suspended in 125µl of CryoStor per extraction (Stem Cell technologies #7952). 250µl of pooled cells were transferred to 0.26ml 3d barcoded vials (Azenta Life Sciences #68-0303-01) before being placed and Cool Cell (Corning) in a -80°c freezer then moved to -190°c LN2 for long term storage. Composition of tube racks was tracked using a Ziath Cube barcode scanner (Azenta Life Sciences).

### Computational methods

#### Data processing and quality control

##### Single-cell RNA sequencing processing

Raw reads were processed using https://github.com/wtsi-npg/bambi to generate CRAM files and tag decoding. The resulting CRAMs were demultiplexed in each pool by using samtools split and converted to FASTQ using samtools fastq. Single-cell 3′ FASTQ files were then processed using 10x Genomics Cell Ranger v6.1.1. Reads were aligned to the human reference genome GRCh38 (GENCODE v32/Ensembl98), excluding intronic reads, and gene–cell count matrices were generated for each pool. YASCP pipeline v1.7 (https://github.com/wtsi-hgi/yascp) was used to process raw count matrices and generate donor level single-cell counts annotated with relevant sample metadata. The pipeline processing steps include ambient RNA removal, doublet detection using 5 different methods, donor deconvolution using Vireo ^37^, automated cell type assignment for each sequencing pool and retrieval of relevant sample metadata (see following sections). Specifically, ambient RNA removal was implemented using CellBender (v0.2.1)^33^, using the parameters described in^90^.

##### Donor Deconvolution

For donor deconvolution, the YASCP pipeline framework uses cellSNP (v1.2.2)^91^ to extract single-cell allele counts and then Vireo (v0.5.6)^37^ in genotype-aware mode to assign cells to specific samples, relying on expected donor genotypes as priors to assign cell barcodes. For this analysis, we curated a pool-specific list of SNPs derived from merged UKB and G&H genotypes. This panel prioritised high-quality common variants (INFO ≥ 0.9, MAF ≥ 0.01), focusing on exonic and UTR regions while excluding sex chromosomes and the MHC to avoid analytical complexities. Each pool was further supplemented with additional variants specific to the expected donors (MAF ≥ 0.0001, applying the same quality filters), thereby improving the resolution of rare alleles. Cells labeled as unassigned or as doublets by Vireo, as well as cells assigned to unexpected donors or to the same donor multiple times within a pool, were excluded from the analysis.

##### Pool- and donor-level quality assessment

Libraries with fraction of reads in cells < 70% were excluded, in line with the 10x Genomics Cell Ranger guidelines ^92^. After donor deconvolution, donors with fewer than 500 unique molecular identifiers (UMIs) were excluded, as they represented outliers in the per-cohort distribution and likely reflected deconvolution errors or issues in sample processing or pooling.

##### Evaluation of donor deconvolution robustness

To control for potential sample swaps occurring during experimental protocols, we applied Vireo in genotype-unaware mode. This approach provides an unbiased validation of the assignments, as it does not use the genotypes of the expected donors as priors, thereby ensuring that assignment probabilities are not inflated by expected genotypes when the actual match was low. For this analysis, we used the ∼7.4 million common SNP panel provided by cellSNP^91^. To validate the accuracy of cell-to-donor assignments, we compared, within each pool, the barcodes of the expected donors deconvoluted with Vireo in genotype-aware mode to those of all donors deconvoluted with Vireo in genotype-unaware mode. We retained the donor for further analysis when the majority of a donor’s barcodes in the genotype-aware modality matched the same donor’s barcodes in the genotype-unaware modality.

We further assessed the robustness of the deconvolution at the cell level using a jackknife cross-validation. For each pool, deconvolution in aware-mode was repeated ten times, each using a random subset comprising 60% of the variants in the deconvolution panel. Cells were excluded from downstream analysis if their donor assignment changed in any iteration. Additionally, deconvoluted donors in which more than 10% of cells were labeled as ‘jumping’ were excluded from further analysis.

Additionally, we compared each cell’s allele counts, as determined by the cellSNP algorithm, to the imputed genotype of the corresponding donor. For this analysis, we first selected candidate sites in each cell that met the following criteria: (i) covered by at least one read, (ii) not A>G variants, to reduce bias from RNA editing, and (iii) homozygous in the imputed genotype data, as single-cell coverage is insufficient to reliably assess heterozygous sites. Within these candidate sites, reads carrying an allele discordant with the assigned donor’s genotype were considered discordant. For each cell, we calculated a discordance ratio, defined as the total number of discordant reads divided by the total number of reads covering the evaluated sites. Cells with a discordance ratio greater than 0.3 were excluded. For each cell, we also computed the discordance ratio against every donor expected in the pool and cells were discarded if the lowest discordance ratio did not correspond to the assigned donor.

##### Doublet detection

To identify and remove potential cell multiplets, we relied on three independent, transcriptome-based computational methods implemented in the YASCP v1.7 workflow: Scrublet (v0.2.3)^35^, using expected multiplet rate of 0.1 and a reference distribution of 100,000 simulated doublets generated from raw counts; scDblFinder (v1.16.0)^36^, run with default parameters on raw counts; and scds (v1.18.0)^34^, which was applied using a hierarchical combination of its internal algorithms. Cells classified as multiplets by at least two of these three methods were flagged as doublets and excluded from all downstream analyses.

#### Normalisation

We normalised CellBender corrected counts for each cohort separately using the Normalization And Data Aggregation (NADA) pipeline (https://github.com/HTGenomeAnalysisUnit/nada_smk), developed for this study. The pipeline implements a Proportional Fitting + log1p + Proportional Fitting (PFlog1pPF) normalization algorithm. We applied this method by scaling the raw counts of each cell by the total read count across the entire cohort. We selected this approach for its high performance in terms of scalability and variance stabilization compared to alternative normalization methods, as reported previously^93^.

#### Donor and cell-level quality control

We applied a two-step filtering strategy to remove low-quality cells. Following cell type annotation, we first filtered out any donors with fewer than 100 assigned cells. Subsequently, we removed outlier cells using a robust, cell-type adaptive filter based on the Median Absolute Deviation (MAD) of several key quality control (QC) metrics. These metrics included the log10-transformed number of UMIs per cell, the log_10_-transformed number of genes detected per cell, the proportion of UMIs in the top 50 most expressed genes, and the percentage of mitochondrial reads. The distribution of these metrics in processed cells is represented in **Supplementary Fig. 19a**. In our cell type-specific MAD filter, cells were removed if their values fell outside a 5 MAD tolerance from the median of their assigned cell type (at the CT2 annotation level). We also applied a hard filter to exclude cells with a mitochondrial read percentage exceeding 20%. The comparison of filtering impact on each cell type when using our strategy (cell type-specific MAD), the same approach applied globally on all cells, or canonical hard-filters, showed that our strategy results in more uniform cell selection, without penalizing rare / specialised cell types (**Supplementary Fig. 19b**). Finally, we removed cells marked as likely artefacts during manual curation of cell clusters and cells not having a consistent cell-type annotation along the four granularity levels. The overall results of cell QC filtering is summarised in **Supplementary Fig. 2.** In addition, we prepared a restricted dataset where we ensured uniform ancestry in the two cohorts and removed duplicated samples mapped to the same individual. Starting from the QCed dataset we applied the following procedure: first, all frozen samples were removed. Second, ancestry filtering was applied based on principal component analysis, retaining only individuals of European ancestry for the UKB cohort, and of Pakistani or Bangladeshi ancestry for the G&H cohort (see Ancestry analysis section). Finally, for individuals represented by multiple samples, only the sample with the highest number of quality-control–passing cells was retained to avoid duplication. This resulted in a dataset including 6.73M cells from 4,452 UKB individuals with inferred European ancestry and 2.25M cells from 1,225 G&H individuals (497 British Pakistani, 728 British Bangladeshi). The restricted dataset was used in analysis of the cell compositional traits (see Cell compositional traits variation along biological and risk factors), MixMIL analysis (see MixMIL), gene expression variance decomposition (see corresponding section), and eQTL discovery (see Expression quantitative trait loci (eQTLs)). The other analyses are based on the QCed dataset (see **Supplementary Table 6**).

#### Annotation Strategy and Reference Atlas Integration

##### Hierarchical Cell Type Annotation

We assigned cell types using a hybrid approach that combined automated prediction tools with manual annotation based on unsupervised clustering and marker gene expression. Automated predictions served as seeds for manual curation, which was refined using clustering results and marker-based interpretation. These automated annotations were generated using the YASCP pipeline (v1.7) and applied to all cells by sequencing pool, regardless of whether they passed QC filters. This choice informed our cell type-guided filtering strategy described in paragraph Donor and cell-level QC.

The model used included Azimuth (’Human - PBMC’)^39^ and CellTypist with several references (’COVID19_Immune_Landscape’, ‘COVID19_HumanChallenge_Blood’, ‘Immune_All_Low’, and ‘Immune_All_High’)^40,41^. A core set of cells from UKB was then selected for annotation refinement, namely confidently assigned singlet cells (see deconvolution and doublet detection sections) from samples processed from fresh cell material. For these cells, we used scVI^94^ embeddings generated both across the dataset and within specific immune compartments (T/ILC, B/plasma cells, monocytes), re-computing highly variable genes (n=2,000) and neighbours for each subset. Leiden clustering was applied across a range of resolutions within each subset, and cluster identities were assigned based on canonical marker gene expression, with support of automated label transfer predictions. This strategy enabled high-confidence cell type annotations by integrating automated predictions with manual inspection of marker genes and clustering results. Within this core set, annotations were structured hierarchically across five levels to reflect biological lineage and cell type granularity: L1: Lineage (e.g., Lymphoid, Myeloid, Progenitor), L2: Lineage subtype (e.g., B/plasma, T cells), CT1: Major cell type (e.g. B cell, CD4^+^ T cells, CD8^+^ T cells) CT2: Subtype/State (e.g. naïve B cells, Effector Memory (EM) CD4^+^ T cells, naïve CD8^+^ T cells) (**Supplementary Table 4**). These core annotated cells formed the basis for building a reference atlas.

##### Integration reference model

To generate a high quality reference atlas we systematically evaluated multiple integration strategies, including Harmony^95^, scVI^94^, scANVI^38^, scPoli^38,96^ and compared them with PCA (**Supplementary Fig. 3**). Following a first hard-filter cutoff for single cell quality, only high-quality fresh cells from the UKB donors were used as a core reference (see Hierarchical Cell-type annotation section). We selected the top 5,000 highly variable genes, excluding mitochondrial, ribosomal, and sex-linked genes as well as BCR/TCR repertoire genes, to minimize integration artifacts. Integration models were trained using sequencing pool as the batch covariate. Performance was assessed using the scIB benchmarking suite^97^, which evaluates both batch mixing and biological conservation. Among the tested approaches, scANVI consistently achieved the highest scores across metrics, and was therefore selected as the final integration method for constructing the reference atlas. Overall, population-scale single-cell atlases define the landscape of cellular variation underlying health trajectories and disease risk. Unlike small datasets that sample only extreme conditions, large-scale atlases enable robust identification of cell states, gene programs, and donor-level variation across a biological continuum. Constructing such atlases requires robust, error-free protocols and careful choice of experimental and data processing steps. For instance, in this context careful single-cell integration was critical to avoid artificially merging distinct manifolds while preserving meaningful variation. Our strategy, based on a high-quality core reference and transfer learning to embed additional samples, was shown to well balance batch correction with biological conservation, minimising technical and donor-driven artifacts without correcting away subtle population-specific or ageing-associated signals, and providing a stable yet flexible framework for discovering reproducible cellular states. The scANVI pre-trained model is available for download to annotate curated celltype in new PBMC datasets.

##### Reference-based label transfer

We used the core reference to annotate the full dataset via label transfer, leveraging the hierarchical annotation structure (L1: lineage; L2: broad cell type; CT1 and CT2: increasingly refined subtypes). Label transfer was performed using the trained scANVI ^38^ reference model and scArches framework^42^. The query datasets included cells excluded by the initial hard filters, additional sequencing batches processed at later stages, and cells from the G&H cohort. These datasets were embedded into the reference space by training scArches adapters following established best practices, and annotations were transferred independently for each level of the hierarchy. Notably, the core reference included a subset of cells deemed low-quality after manual refinement due to their mixed marker genes expression profiles. The inclusion of these cells allowed for the transfer of both high-confidence cell type labels and low-quality QC labels to the query cells via weighted k-nearest neighbor propagation on the shared embedding, facilitating the subsequent lineage-specific filtering using MAD-based cutoffs. By modeling the distribution of QC metrics (e.g., % mitochondrial reads, number of genes, number of UMIs) within each annotated lineage, we could stringently remove these low-quality cells while preserving rare or borderline populations. To assess label transfer quality, we inspected uncertainty scores, compared marker gene expression between reference and query, and verified consistency across annotation levels. Since annotations were transferred independently at each level, we applied an additional consistency filter to remove cells with discordant labels across the hierarchy (e.g., cells assigned to the lymphoid lineage at L1 but classified as CD14⁺ monocytes at CT2). A total of 495,447 (4%) of cells showed such inconsistencies and were excluded from downstream analysis due to high uncertainty scores (>0.5) and poor marker concordance.

##### Definition of cell states

Cell type annotation at the CT3 level (cell states) was performed on the full dataset containing both UKB and G&H cohorts in a cell compartment-wise fashion. Based on the cell type labels obtained using label transfer with scANVI and scArches, cells were subsetted to include cells only from one L2 level (B/plasma, HSC MPP), CT1 level (e.g. NK, ILC, MAIT, γδ T, Basophil) or CT2 level (CD4^+^ Treg, CM CD4^+^ T cells, naïve CD4^+^ T cells, CM CD8^+^ T Cells, EM CD4^+^ T cells, naïve CD8^+^ T cells, EMRA CD8^+^ T cells), based on the heterogeneity and number of cells in the compartments. Cells were then clustered in Scanpy v1.9.6 ^98^ with Leiden clustering using the scANVI embeddings, and clusters annotated based on a combination of CellTypist and Azimuth predictions (using models listed above) and manual annotation based on differentially expressed genes between clusters (**Supplementary Fig. 5**). A dictionary of the high-resolution cell states, their functional description and marker genes is available in **Supplementary Table 4**.

Differences between cell type composition between the UKB and G&H cohorts at CT3 level were tested using Mann-Whitney U-test on CT3 fractions out of total cells per donor, followed by Benjamini-Hochberg adjustment for multiple testing.

##### Blood composition archetypes and subtypes

To identify subgroups of donors based on the composition of co-varying cell types, a donor-by-cell-type matrix with each feature representing the proportion of a given CT3 cell type within the donor’s cells was generated. Neighbours and graph-based Leiden clusters of donors reflecting similar cell type compositions were defined using Scanpy with default parameters. Cell composition clusters enriched in similar cell types were merged to obtain blood subtypes termed archetypes. The archetypes were visualised using a UMAP embedding computed using the neighbours in the cell composition space. To identify characteristic cell types for the archetypes, differences between cell type composition across the archetypes were tested using Mann-Whitney U-test on CT3 fractions out of total cells per donor, followed by Benjamini-Hochberg adjustment for multiple testing. Enrichment of cohort (UKB vs G&H) membership, predicted CMV serostatus and recent illness prior to sampling date based on questionnaire data from G&H participants were tested across the archetypes using Fisher’s exact test. For evaluating enrichment of CD5+ B cells between age-matched donors between cohorts, the numbers of donors with at least 0.1% or 1% B_CD5 out of total cells were quantified in UKB and in G&H restricted to the UKB age range of 52-85 years, followed by a Fisher’s exact test.

##### CMV serostatus classification

Cytomegalovirus (CMV) serostatus was inferred from plasma antigen measurements for 93 donors with available data in UKB and subsequently predicted from cell type composition for the full Cardinal cohort. Donor-level CMV reactivity was quantified using measurements for multiple CMV antigens included in the serology panel (pp150-N, pp52 and pp28), which were aggregated to obtain a single CMV reactivity score for each donor. CMV serostatus was defined by fitting a two-component Gaussian mixture model to the distribution of donor CMV reactivity scores using the *GaussianMixture* implementation from the Python package scikit-learn. Donors were assigned to the component with the higher mean reactivity as CMV seropositive and to the lower-mean component as CMV seronegative.

Cell type composition for each donor at CT3 level was organised into a donor-by-cell-type matrix, with each feature representing the proportion of a given cell type within the donor’s cells. Donors were randomly divided into training (80%) and held-out test (20%) sets using stratified sampling implemented with the *train_test_split* function from scikit-learn, preserving the proportion of CMV-positive and CMV-negative donors between partitions.

A random forest classifier was trained to predict CMV serostatus from donor-level cell-type fractions using the *RandomForestClassifier* implementation in scikit-learn. Hyperparameters were selected using nested cross-validation within the training set. In the inner loop, a grid search across the number of trees, minimum number of samples per leaf, and the number of features considered at each split was performed using the *GridSearchCV* function with stratified k-fold cross-validation. The area under the receiver operating characteristic curve (AUC) was used as the optimization metric. In the outer loop, model performance was evaluated on held-out validation folds to estimate expected predictive performance and to record optimal hyperparameters across folds. The most frequently selected hyperparameter configuration across folds was used to train the final model on the full training set.

Model performance was assessed on the independent test set using the area under the receiver operating characteristic curve calculated with the *roc_auc_score* function from scikit-learn. Classification probabilities for CMV seropositivity were obtained from the fitted random forest model and thresholded at 0.5 to generate binary predictions. The trained model was subsequently applied to the full Cardinal cohort, including both UKB and G&H cohorts, by constructing sample-by-cell-type fraction matrices with the same feature set and ordering as used during model training to generate predicted CMV probabilities and binary classifications for each donor.

#### Medical record data

Electronic demographic and health record data were accessed through UKB-RAP and the Genes & Health TRE, under approved research protocols, which provide secure access to individual-level data.

Disease definitions were derived from the MVP–FinnGen–UK Biobank meta-analysis available at https://mvp.finngen.fi/, which provides phecodes for 330 diseases. Phecodes were employed as they aggregate individual ICD codes into clinically coherent categories, thereby reducing data sparsity and increasing statistical power in association analyses. Comparability of disease definitions across UKB and G&H was ensured through harmonised phecode mappings: each phecode was mapped to ICD-10, Read2, and CTV3 codes, (ICD10 and SNOMED for G&H) facilitating the extraction of diagnostic information from multiple sources, including Hospital Episode Statistics (HES), cause of death records, cancer registries, and general practitioner (GP) clinical data. For UK Biobank, diagnoses were extracted using the *ukbrapR* package (https://github.com/lcpilling/ukbrapR), and the mapping between phecodes and diagnostic codes was obtained from the UK Biobank Read-to-Phecode repository https://github.com/spiros/ukbiobank-read-to-phecode/tree/master^99^. For the Genes & Health dataset, three- and four-digit ICD-10 codes were used.

To address incompleteness in disease representation, additional autoimmune and mental health disorders were incorporated, resulting in a total of 350 diseases. Three diseases were subsequently excluded from the analysis due to duplication (essential hypertension) or issues with diagnostic ascertainment (glomerular disorders in diseases classified elsewhere and kidney transplant status).

Implausible diagnosis dates (e.g., those recorded prior to birth) were removed. The medical records prevalence is reported in **Supplementary Table 2**.

#### Cell compositional traits variation along biological and risk factors

##### Computation of single-cell complete blood counts (scCBCs)

Single-cell complete blood counts (scCBCs) were calculated by integrating quantitative measurements from the Sysmex haematological analyser with single-cell profiles generated using the 10x Genomics platform. Total lymphocyte and monocyte counts were obtained from Sysmex data, and the relative proportions of each immune cell subset within these compartments were estimated from the 10x dataset.

Absolute abundances for each cell type were derived as:

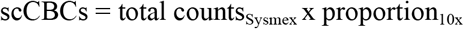

with calculations performed separately for lymphoid and myeloid lineages. This transformation removed the apparent negative correlations between cell type proportions due to compositional constraint (**Supplementary Fig. 6**).

scCBCs were computed for 4,094 UK Biobank donors and 985 Genes & Health donors. Cohort selection criteria and outlier removal procedures are described in Zanotti et al. ^26^, where cohort demographics are also reported (Table 1). All downstream analyses involving scCBCs were conducted using this cohort.

##### Associations between scCBCs and major axis of biological variation

We modelled cell-type abundance as a function of key biological and environmental factors, including age, sex, ancestry, body mass index (BMI), smoking status, and the index of multiple deprivation (IMD). Age was defined at PBMC collection, and BMI and smoking status were taken from the most recent available assessment. In G&H, smoking status and BMI measurements were recorded for a smaller subset of participants (*N* = 773 and 770, respectively), resulting in a reduced effective sample size. IMD, a composite measure of socioeconomic status, was standardised within England, Scotland, and Wales before being combined into a single continuous variable; in G&H, IMD was available only for North East London participants (N = 772).

For each immune cell type, linear models were fitted to assess associations with biological variables: UKB:

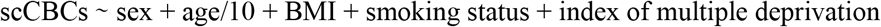

G&H:

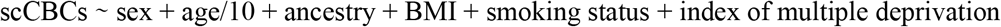

Prior to modelling, scCBCs were variance-scaled while maintaining their original mean values to preserve meaningful differences in cell type abundance. Continuous covariates (BMI and IMD) were standardised to z-scores, whereas categorical variables (sex, ancestry, smoking status) were encoded as binary or multi-level factors. Age remained unstandardised to retain units of years and facilitate interpretability.

##### Associations between scCBCs and disease diagnosis

Associations between single-cell complete blood counts (scCBCs) and disease status were assessed using generalised additive models (GAMs), which allow non-linear associations between covariates and immune cell abundances. Analyses were limited to diseases represented by at least 35 cases. Separate models were fitted for each cell type, as specified below:

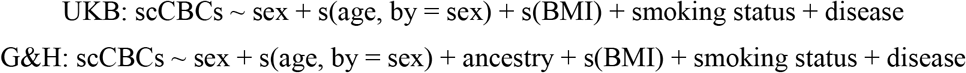

where *s*(⋅) denotes smooth functions estimated using B-spline basis expansions. Covariates were processed in accordance with prior analyses: age and BMI were standardised to z-scores, and sex, smoking, ancestry, and disease status were encoded as categorical variables. To ensure comparability across models, scCBCs were variance-scaled within cell types while preserving mean biological differences. In G&H, smoking data and BMI information were available for N=773 and 770 participants, respectively. Control cohorts were stringently defined as individuals without diagnoses belonging to the same disease category (e.g., controls for type 2 diabetes excluded all metabolic disorders), mitigating potential misclassification and class imbalance. Participants with malignant neoplasms were excluded from both case and control groups to isolate disease-specific immune variation from oncological effects. The p values for disease effects were adjusted for multiple testing using the Benjamini–Hochberg false discovery rate procedure. All models were fitted in *R* using the mgcv package ^100^ (v1.9-1), with smoothness parameters estimated via restricted maximum likelihood (REML). More details are reported in^26^.

#### MixMIL (Cell states - disease association)

We employed an attention-based Multiple Instance Learning (MIL) method, MixMIL^51^, to perform donor-level label prediction using as inputs the 50 scANVI single cell features (Integration Reference Model paragraph in Methods) and donor level covariates (sex, age, BMI, smoking status). We trained the model to predict case-control status using a 3-fold cross-validation approach for UKB and 5-fold for G&H, downsampling controls to match the number of cases for each disease. The disease diagnoses were selected among those with at least 100 cases in UKB (at least 50 cases for G&H), comprising 29 and 32 diseases category in UKB and G&H, respectively.

The model outputs donor-level predictions, single-cell attention weights, and feature importance. To obtain a cell-type level score (for each donor and for each trait predicted by MixMIL), we used an attention enrichment score, computed as the ratio between the proportion of attention given to a certain cell type and the proportional abundance of the same cell type. More details are reported in a separate manuscript^29^.

#### Gene expression variance decomposition

##### Pseudobulk gene expression preprocessing and sample selection

For each cohort and each of the eight CT1 cell types (Monocytes, CD8^+^ T cells, CD4^+^ T cells, NK, B cells, DC, MAIT, γδ T cells), we averaged PFlog1pPF normalised gene expression per donor (see Normalisation section) and transformed each gene using a rank-based inverse-normal transform. We restricted the analysis to the same donor and sample inclusion criteria as the eQTL pipeline (see Expression quantitative trait loci (eQTLs) section) but analysed British Pakistani and British Bangladeshi individuals from the G&H cohort jointly. Genes were retained when they had non-zero expression in at least 10% of donors.

##### Variance component model

For each gene, the cis-genetic kernel *K^g^* was computed as a linear kernel over the standardised cis SNPs within ±1 Mb of the transcription start site. Analogously, the EHR kernel *K_EHR_* is linear in the EHR features: one-hot-encoded phecode diagnoses retained at ≥20 cases per cohort (see Medical record data), three-level smoking status (current / previous / never), and BMI. We accounted for covariates *F*, namely sex, age and 10 genetic PCs. Therefore the variance estimates are to be interpreted as a fraction of total variance after accounting for sex and age.

For each gene × cell type pair’s gene expression *gex* we fitted the variance decomposition model:

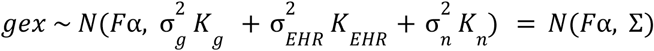

using *limix*^58,101^, A genetics-only null model:

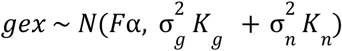

was used to compute the likelihood-ratio statistic:

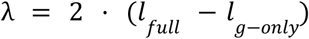

quantifying the additional variance captured by *K_EHR_*.

##### False discovery rate control

To control FDR empirically, for each gene we permuted the rows of the EHR feature matrix once and refitted the full model, yielding a null LRT per gene (λ_0_). Within each cell type, we estimated FDR at threshold t as:

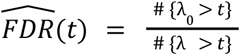

and called genes significant at the most permissive *t* with *FDR*(*t*) ≤ 0. 05^102^.

##### Best linear unbiased predictor and trait association

To quantify the donor level expression attributed to EHR, we derived the best linear unbiased predictor (BLUP) of the EHR component:

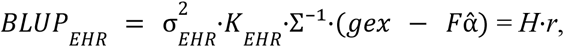

where Σ⁻¹ is the inverse of the full covariance matrix. To remove in-sample effects we further derived the leave-one-out BLUP^103^:

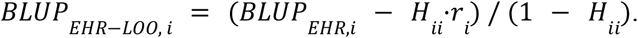

This single-kernel form from^103^ is an approximation to the exact multi-kernel leave-one-out BLUP. We then computed for each gene x cell type combination Pearson’s r between the *BLUP_EHR_*_−*LOO*_ and each EHR trait across donors. These correlations are the input to the downstream latent-program and tensor-decomposition analyses.

##### Split-half robustness analysis

To assess stability of the variance fractions and BLUPs, we split UKB Monocyte randomly into two halves and reran the full pipeline independently on each half. We report reliability metrics across the two splits in **Supplementary Fig. 9**.

#### Latent programs discovery

Variance decomposition results from the UKB and G&H cohorts were integrated to construct a unified feature tensor (cell types X genes X traits). First, to establish a consensus set of associations, we took the union of significant cell-gene pairs across both cohorts, restricted to pairs assayed in both datasets. We then identified significant cell-gene-trait combinations using a cell-type-specific Bonferroni significance threshold (*P* < 0. 05 / [*N_genes_ X N_traits_*]). Any combination passing this threshold in at least one cohort was retained, provided the trait was evaluated in both cohorts. For all retained cell-gene-trait combinations, the corresponding effect size estimates (Pearson’s r) were extracted to populate the data structure. To prevent ubiquitous translation-related signals from disproportionately driving the decomposition, genes encoding standard and mitochondrial ribosomal proteins were systematically excluded from the final input tensor.

The association effect sizes were structured into a four-dimensional tensor defined by 1003 Genes, 8 Cell Types, 85 Traits, and two Cohorts to identify shared and condition-specific multidimensional interaction modules. Tucker tensor decomposition was performed utilizing the Higher-Order Orthogonal Iteration (HOOI) algorithm implemented in the rTensor R package^104^. Optimal tensor ranks were evaluated using a two-step approach: initial rank boundaries were estimated via the elbow method on cumulative variance scree plots derived from the singular value decomposition (SVD) of each unfolded tensor mode. These estimates were refined through parallelised grid-search cross-validation, wherein 5% of non-zero tensor elements were randomly masked. The final optimal rank configuration (Genes=12, Cell types=6, Traits=6, Cohorts=2) was selected by minimizing the out-of-sample root mean square error (RMSE) upon tensor reconstruction (**Supplementary Fig. 20**). To elucidate the biological functions of the extracted gene modules, genes were ranked by their continuous loading weights within each dimensionally reduced program. Gene Set Enrichment Analysis (GSEA) was executed on these ranked lists using the fgseaMultilevel algorithm (50,000 permutations) from the fgsea R package (v1.33.1)^105^ against human gene sets retrieved via msigdbr (v26.1.0). Pathway collections assessed included Reactome, GO Biological Processes (GO-BP), MSigDB Hallmark, and ImmuneSigDB, with significantly enriched biological functions defined by a Benjamini-Hochberg adjusted p-value < 0.05) (**Supplementary Fig. 13**; **Supplementary Table 17**; **Supplementary Table 18**; **Supplementary Table 19**; **Supplementary Table 20**).

To test whether contextual eQTL (F.E. ≥ 1.1) genes were enriched among genes whose program contribution diverged most between cohorts, we investigated their distribution among genes with highly divergent activation between G&H and UKB cohorts. We first computed a cohort gene coupling score as the sum of the core tensor value for the gene program, trait program, cell type program combination weighted by the cohort matrix loadings and aligned to the dominant cell type direction. We then computed for each gene a cohort-specific activation score across all gene-celltype-trait program combinations by multiplying the gene loading in the gene program by the corresponding cohort gene coupling. Finally, we took for each gene in each combination the absolute difference between the cohort-specific activation score (final_score_diff). For each gene-program × trait-program × cell-type combination, genes were ranked by final_score_diff (highest divergence first) and tested with fgsea (v1.36.2); pre-ranked GSEA, minSize = 10, maxSize = 500, 10,000 permutations, eps = 0) for enrichment of cell-matched, G&H- or UKB-specific contextual (F.E. ≥ 1.1) eGenes toward the high-divergence end of the ranking.

#### Differential Gene expression analysis

Starting from the QCed dataset (see Donor and cell-level quality control section), pseudobulk RNA-seq counts were obtained by summing raw single-cell expression values per donor for each CT2 level cell type, using the NADA pipeline (refer to Normalization section for details). Samples with less than 10 cells per cell type and duplicated samples from the same donor were removed, keeping the replicate with the highest cell count. Frozen samples were excluded to avoid batch effects. Samples were filtered to retain donors matching the cohort’s primary ancestry (Caucasian for UKB, South Asian for G&H), with ancestry outliers removed.

Differential gene expression analysis was performed independently for each cell type using the limma-voom ^106^ workflow. For each pseudobulk count matrix, genes were filtered using edgeR *filterByExpr* ^107^, which performs model-aware filtering to retain genes with adequate counts given the experimental design, normalization factors were calculated with edgeR calcNormFactors, and counts were transformed to log_2_ counts per million with precision weights using voom. Weighted linear models were fitted including covariates and differential expression statistics were computed using lmFit followed by empirical Bayes moderation with eBayes. Individual covariates included sex, ancestry, scaled age, scaled BMI, smoking status (current smokers) and the first 4 gene expression PCs to account for technical variation. These PCs were calculated separately for each cell type using prcomp on unfiltered log gene pseudo-bulk matrices normalised with calcNormFactors followed by log2 counts-per-million normalization. Samples with missing covariate values were excluded prior to modeling. Associations of gene expression with each covariates were extracted from the full model, thereby accounting for all other covariates. For disease/trait testing, we first selected traits with an overall minimum of 50 cases and then, for each cell type subset, required a minimum of 5% cases and 40 samples in total for G&H, or a minimum of 2 cases and 2 controls for UKB. Disease status was then added to the model one trait at a time. A summary of the number of genes, individuals, and significant results at Benjamini–Hochberg false discovery rate (FDR) < 0.05 is shown in **Supplementary Table 27** and **Supplementary Table 28**.

Gene set enrichment analysis was performed with the GSEA function from clusterProfiler package ^108^ (v4.2.2). Cell matched G&H contextual (F.E. ≥ 1.1) eGenes (min. 4 genes, max. 290 genes) were used as gene sets to test enrichment in upregulated or downregulated genes from the differential gene expression analysis ordered by log Fold Change.

### Genetic analyses

#### Common variant genotype preparation

We obtained imputed genotypes in GRCh37 coordinates for the UK Biobank dataset under project access 69938. These genotypes are imputed based on HRC, UK10K and 1000G European reference panels as described in ^30,109^. For Genes&Health cohort, genotypes are determined using Illumina GSAv3EAMD chip and then imputed genotypes in GRCh38 coordinates were obtained from the TOPMed imputation server using the TOPMed reference panel r3 ^110^. This enabled analyses of common variants (MAF > 0.1%), analyses of rare variants from sequencing was not performed in this manuscript.

From each imputed dataset, we first extracted only the samples also sequenced in the single-cell dataset. We then removed multi-allelic variants and variant IDs were renamed as chr:pos:A1:A2, where A1 and A2 are REF and ALT alleles sorted in alphabetical order. Since this may create conflicting IDs for some INDELs we further removed any variant where the renaming created a duplicated ID. Variants were filtered and saved to PGEN format using plink2 according to the following criteria:

- MACH-R2 (similar to INFO score) ≥ 0.5 (--mach-r2-filer 0.5)
- HWE test p-value < 1 x 10-8 (--hwe 1e-8 ‘midp’ ‘keep-fewhet’)
- MAF ≥ 0.001 (--maf 0.001)

The resulting variants for the UKB dataset were then converted to GRCh38 coordinates. Using the bcftools liftOver plugin^111^ and the hg19ToHg38.over.chain.gz chain file from UCSC genome browser [https://hgdownload.soe.ucsc.edu/goldenPath/hg19/liftOver/hg19ToHg38.over.chain.gz] we mapped original coordinates, REF and ALT to GRCh38 genome build. We then applied a series of additional checks on successfully converted variants to ensure coordinate conversion consistency:

- Removed any variant were the new chromosome is not equal to the original one
- Removed variants were the ALT allele could not be identified univocally, resulting in multiple possible ALT alleles
- Removed new duplicated or multi-allelic variants generated when the liftover processed remapped two originally distinct variants on the same position in the GRCh38 build

These filters removed approximately 15,000 variants. When REF and ALT alleles were swapped as a result of liftOver we updated the genotype matrix accordingly using plink2 *--ref-allele force*. The final working datasets included 17,223,908 and 15,437,811 autosomal variants for the UKB and G&H dataset, respectively.

#### Ancestry analysis

To characterise the genetic ancestry of CARDINAL participants, we performed principal component analysis (PCA) on cohort-level genotype data. For each cohort (UKB and G&H), genotypes were processed separately, retaining only directly genotyped variants (i.e., excluding imputed SNPs). These were then merged with the gnomAD ^112^ subset of the 1000 Genomes Project (1KGP) ^113^ and Human Genome Diversity Project (HGDP) ^114^ reference panel, restricting to overlapping sites with concordant allele coding. After quality control (genotype missingness < 0.01) and LD-based pruning, we computed PCA using FlashPCA2 ^115^, first on a worldwide reference set and then on subsets restricted to South Asian and European populations to increase resolution of fine-scale structure within these groups. CARDINAL samples were projected onto the reference PCA space using pre-computed SNP loadings. Ancestry proportions were estimated using the PANE framework ^116^, which applies non-negative least squares (NNLS) regression on the first 10 principal components to infer the proportion of genetic relatedness of each individual to a set of reference sources. CARDINAL individuals were used as targets, while 1,837 individuals from 21 reference populations spanning six macro-regions (Africa, America, Asia, Europe, Middle East, and Oceania) drawn from the HGDP and 1KGP served as sources, selected to capture the major axes of genetic diversity within each macro-population. Each CARDINAL individual was assigned to an ancestry group when more than 50% of their inferred ancestry derived from a single macro-population. Additionally, G&H participants were further sub-grouped into British Pakistani and British Bangladeshi ancestry by comparing their position within the South Asian PCA cluster to PCA-based ancestry estimates available from the original Genes & Health cohort.

#### Variant annotation

Genetic variants used in eQTL analysis were annotated using Variant Effect Predictor (VEP v111) and vcfanno^117^ to obtain gene level consequences and other useful information, including gnomAD v4.0^65^ global, Non-Finnish European (NFE), and South Asian (SAS) AF, LOFTEE^118^ LoF impact predictions, GREEN-DB v2.5 regulatory regions^119^. The eGene biotype was annotated based on GENCODE v32 definitions. Based on the combination of gene consequences reported by VEP and additional annotations we defined simplified variants consequence labels as follows: 3_prime_utr, 5_prime_utr, coding_sequence, downstream_gene (5kb downstream of a gene feature), intronic, loss-of-function (LoF, including stop_loss, stop_gain, start_loss, frameshift indels, splice_donor, splice_acceptor), LoF-HC (LoF variant classified as high-confidence by LOFTEE), no_consequence (none of the other classes is applicable, hence the variant has no direct consequence on the eGene), non_coding_transcript_exon, prot_changing (including missense and inframe indels), splice_region (near splice site, namely 1-3 bases of the exon or 3-8 bases of the intron, or annotated by VEP as splice_donor_5th_base, splice_polypyrimidine_tract), synonymous, upstream_gene (5kb upstream of a gene feature).

#### Expression quantitative trait loci (eQTLs)

##### eQTL association mapping

Cis-expression quantitative trait loci (cis-eQTL) association analyses were conducted on the restricted dataset (see Donor and cell-level quality control) using TensorQTL^120^ as implemented in the QTLight pipeline (https://github.com/wtsi-hgi/QTLight/tree/main). As phenotypes, we used the inverse rank normalised pseudo means estimated in each of the available CT2 cell types. We considered all SNPs within a ± 1Mb window from each gene transcription start site (TSS), and with minor allele frequency (MAF) greater than 0.1% (0.001) and minor allele count (MAC) of at least 10. All associations were run separately for the three main ethnic groups in each cohort: UK Biobank Europeans (N = 4452), G&H British-Pakistani (N = 497) and G&H British-Bangladeshi (N = 728). The normalised UMI counts were aggregated at the individual level by taking the mean across all cells of a given cell type (pseudo-mean) and the inverse rank normal transformation (IRNT) was then applied to the resulting pseudo-mean values. Association analyses were corrected for 20 genetic principal components (gPCs) to account for population structure and 20 expression principal components (ePCs) to account for within cell type differences in cell type composition between donors, and sex at birth. These analyses were run on all cell types in which at least 100 donors had at least 10 cells of the same cell type, which lead to 23 analysed cell types in UKB Europeans, 19 in G&H British-Pakistani and 20 in G&H British-Bangladeshi. Finally, for each cell type, we only tested genes expressed in at least 20% of the individuals, with an additional cutoff of at least 50 individuals. For the G&H cohort, summary statistics from ancestry-specific analyses were subsequently combined by the shared cell type (19) using inverse-variance–weighted fixed-effects meta-analysis. The exact sample size, number of genes tested and number of significant eGenes in each cell type is reported in **Supplementary Table 21**. We construct SNP IDs as chrom:position:a1:a2, with a1 being the first allele in alphabetical order and treated as the effect allele.

To account for multiple testing, we applied the Aggregated Cauchy Association Test (ACAT) ^121^ for each gene–cell type pair to obtain a gene-level p-value. These p-values were subsequently adjusted for multiple testing across all tested gene–cell type pairs using Storey’s q-value approach. Gene–cell type pairs with q-values ≤ 0.05 were considered statistically significant.

##### Fine-mapping and identification gene expression modules across genes and celltypes in UKB and G&H cohorts

Starting from eQTL results of G&H and UKB cohorts, we used the FLANDERS framework ^27^ to perform fine-mapping for each celltype and then identify gene expression modules. Briefly, we first used the Locusbreaker algorithm ^122^ to identify genomic loci associated with gene expression and these were then fine-mapped separately in each population using the Sum of Single Effects (SuSiE) model ^123^ and in-sample LD. Fine-mapped credible sets were grouped into co-regulated modules defined within FLANDERS as groups of gene expression signals across genes and/or cell types that were consistent with sharing the same causal variant, as described in detail in ^27^. Joint fine-mapping was then performed for each module to define a 99% joint credible set underlying all traits included in the module. The variant with the highest posterior inclusion probability (PIP) within each joint credible set, was selected as most likely causal SNP, for comparing effect sizes and directions across traits.

##### Colocalisation of eQTL credible sets between G&H and UKB cohorts

To assess the sharing of genetic regulatory effects across populations, we performed colocalisation analysis between eQTL credible sets across G&H and UKB cohorts, restricted to matching cell type–gene pairs using the FLANDERS pipeline ^27^. This allowed us to identify credible sets likely to share an underlying causal variant between the two cohorts.

##### Colocalisation of co-regulated gene expression modules with traits and diseases in G&H cohort

We used colocalisation analysis to investigate the relationship between modules of co-regulated gene expression traits and complex trait- and disease-associated loci in the G&H cohort. First, we performed fine-mapping on 529 ICD10-defined disease endpoints and 107 quantitative traits GWAS from the full G&H cohort ^110,124^ using the FLANDERS pipeline as described above, but using fixed locus definition thresholds. Specifically, the locus breaker limit threshold was set to 1 x 10^-5^, while the significance threshold was set to 5 x 10^-8^. G&H gene expression modules were then colocalised with finemapped traits using the FLANDERS framework and positive colocalisation events were defined as those with posterior probability of shared causality (PPH4 ≥ 0.75). These are reported in **Supplementary Table 29**.

#### Cross-population comparison

##### Categorisation of eQTL signals

After the colocalisation between G&H and UKB eQTL signals, credible sets were classified based on their posterior probability of a shared causal variant (PP.H4): signals with PP.H4 > 0.75 were classified as population-shared, those with 0.25 ≤ PP.H4 ≤ 0.75 as uncertain, and those with PP.H4 < 0.25 as non-colocalising. A credible set was considered non-colocalising only if it failed to colocalise (PP.H4 < 0.25) with all credible sets for the same gene in the other population. Credible sets that did not meet the q-value threshold or could not be fine-mapped before the colocalisation step were classified as ‘Not tested’.

Non-colocalising credible sets were further subdivided into three categories. Population-specific eVariants were defined as signals driven by a variant monomorphic in the other cohort at the CARDINAL cohort-level. Contextual eQTLs were defined as signals for genes that, despite being expressed in the other cohort, were not an eQTL in that population for the same cell type. In addition, we considered contextual eQTLs credible sets that failed to meet the criteria for eGene consideration in the colocalisation step (gene x cell type expressed in at least 20% of the cohort individuals) in one cohort but not in the other. A subset of contextual eQTLs were further classified as contextual eQTL with fraction of expression [F.E.] ≥ 1.1 when two conditions were met: (i) the ratio of the fraction of donors expressing the eGene in the discovery population to that in the other population exceeded 1.1, and (ii) the eQTL had high statistical power (> 0.95) to be detected in the other cohort, as estimated using the powerEQTL package ^125^. Power was computed via simple linear regression (powerEQTL.SLR), using the top SNP MAF and sample size from the other cohort, the observed effect size, and a family-wise error rate of 2 × 10^-5^. Signals meeting the power threshold but not the expression ratio criterion were thus attributed to genuine regulatory divergence rather than differential detectability.

##### Cell type enrichment for G&H contextual eQTLs F.E. ≥ 1.1

To assess cell type enrichment of G&H contextual eQTLs F.E. ≥ 1.1, we estimated baseline PBMC cell type abundances from G&H single-cell composition data by averaging cell counts per donor and converting them into cohort-wide proportions. For each eQTL category (population-shared, G&H eVariants, and G&H contextual eQTLs F.E. ≥ 1.1), enrichment was calculated as the ratio between the observed and expected number of credible sets per cell type and log2-transformed. Differential enrichment was defined as the difference between the log2 enrichment of each population-specific category and that of population-shared eQTLs within the same cell type. Statistical significance was assessed using Fisher’s exact test on cell type-specific contingency tables, followed by multiple-testing correction.

##### Gene-set enrichment analysis of population shared and population specific eGenes

Over-representation analysis (ORA) of Gene Ontology Biological Process (GO BP) terms was performed using the enricher function from the clusterProfiler package package (v4.2.2) ^108^, with the background gene set defined as the union of all eGenes identified across the UKB and G&H cohorts. GO BP terms were restricted to those containing between 10 and 500 genes. Immune-related GO BP terms were selected by searching terms containing immune-relevant keywords and identifying the set of eGenes captured within these annotations; terms were added until reaching saturation. ORA was then performed on this curated set of immune-related terms for three eGene groups (Pop-shared, G&H-specific, and UKB-specific), with p-values adjusted for multiple testing using the Benjamini-Hochberg procedure (FDR ≤ 0.05).

#### Identification of CD16^+^ adaptive NK-specific cis-eQTLs in UKB and their colocalisation with diseases

Cis-eQTL signals were identified at the CT3 resolution for CD16^+^ NK and CD16^+^ adaptive NK cells cells using SaigeQTL ^126^ as implemented in the QTLight pipeline (https://github.com/wtsi-hgi/QTLight/tree/main) and gene expression raw counts. cis-eQTL analysis was performed following the same procedure described above for CT2 level. Briefly, variants within ±1 Mb of each gene transcription start site (TSS) with MAF > 0.1% and MAC ≥ 10 were tested in UK Biobank Europeans (N = 1789) participants. Models were adjusted for sex, 20 genetic principal components, and 20 expression principal components. Analyses were restricted to donors contributing ≥10 cells and to genes expressed in at least 50 individuals and ≥50% of donors. Multiple testing correction was performed using ACAT to derive gene-level p-values, followed by Storey’s q-value adjustment across all gene–cell type pairs. Associations with q-value ≤ 0.05 were considered significant. Finemapping of significant eGenes and colocalisation analysis was performed using the FLANDERS pipeline as described above (see Resolving cis acting pleiotropy across populations, cell types and genes section).

To define CD16^+^ adaptive NK-specific signals in UKB individuals, we systematically excluded signals shared with other cellular and tissue contexts based on colocalisation analyses. First, CD16^+^ adaptive NK signals were compared with those identified in CD16^+^ NK cells at the same resolution (CT3 level). Second, CD16^+^ adaptive NK signals were compared against all other cell types at CT2 resolution (excluding CD16^+^ NK cells). Third, colocalisation was performed between CD16^+^ adaptive NK signals and bulk tissue GTEx eQTLs credible sets downloaded from Open Targets v.25.09 ^127^. In all comparisons, shared signals were defined as those with a posterior probability of colocalisation (PP.H4) greater than 0.5. Signals meeting this criterion in any of the above comparisons were considered non-specific and excluded. The remaining signals were defined as CD16^+^ adaptive NK-specific cis-eQTL signals.

We subsequently tested such CD16^+^ adaptive NK-specific signals for colocalisation with genetic associations from multiple disease and trait datasets, namely fine-mapped GWAS summary statistics obtained from OpenTarget v25.09 ^127^ for GWAS Catalog (17,641 studies) and FinnGen R12 traits (1,248 studies). We also included a collection of 46 manually curated diseases (**Supplementary Table 30**) and 29 hematological traits measured in the full UKB cohort, for which we performed GWAS using REGENIE v4.1 software ^128^, TOPMed-imputed genotypes and the protocol described by Astle et al ^129^ (see data availability section). Shared signals between CD16+ adaptive NK-specific cis-eQTLs and disease/trait loci were defined using a threshold of PP.H4 ≥ 0.75. To ensure disease associations were specific to CD16+ adaptive NK biology, an additional filtering step was applied: all disease and trait loci were also tested for colocalisation with eQTL signals from all other UKB cardinal cell types at CT2 resolution (excluding CD16+ NK cells), and GTEx bulk tissue eQTLs. Disease associations showing evidence of colocalisation (PP.H4 ≥ 0.75) in any of these non-target contexts were considered non-specific and excluded from further analysis. Finally, we performed Mendelian randomisation analyses using the Wald ratio method to estimate the effect of CD16+ adaptive NK gene expression on trait and disease associations.

#### Mendelian randomisation of BMI on gene expression

To measure the effect of BMI on gene expression at the single cell level, we used Mendelian Randomisation, considering as instruments the 471 independent significant SNPs coming from the UK biobank GWAS. In order to reduce the multiple test burden as exposures we focused on the 956 cell type-gene pairs which resulted significantly associated to BMI from the variance decomposition analysis. For each gene, association testing between the BMI instrument SNPs and pseudobulk expression was performed using a similar approach to eQTL association mapping, but with cis-effect-corrected pseudobulk expression to remove local genetic signal prior to testing. Briefly, for each cell type, we regressed inverse rank normalised pseudobulk mean expression on imputed instrument dosage, correcting for 20 genetic PCs and 20 expression PCs. Analyses were performed with TensorQTL in trans mode via the cellink package (see code availability). After harmonising the two datasets we performed betaMR using the inverse variance weighted method. For each analysis we checked whether there was a detectable heterogeneity in the genetic instruments estimated by measuring Q.

## Acknowledgements

This research was funded in part by the Wellcome Trust [grant number 220540/Z/20/A], the Chan Zuckerberg Foundation [grant number 2020-224316] and by the British Heart Foundation (BHF) Cambridge Centre for Research Excellence RE/18/1/34212. Fondazione Human Technopole is supported by public funds awarded by the Italian Ministry of Economy and Finance, the Ministry of University and Research, and the Ministry of Health.

This work is supported by the project IMPACT (G.A.N.) 101201772 Funded by the European Union. Views and opinions expressed are however those of the author(s) only and do not necessarily reflect those of the European Union or grant authority (ERCEA). Neither the European Union nor the granting authority can be held responsible for them.

This project has received funding from the Innovative Medicines Initiative 2 Joint. Undertaking (JU) under grant agreement No 101007873 (EU-IMI2 101007873).

L.A. was supported by the Helmholtz Association, as part of the joint research school Munich School for Data Science (MUDS) and the Studienstiftung des deutschen Volkes (German Academic Scholarship Foundation).

This work was supported by the Deutsche Forschungsgemeinschaft (DFG) - Projektnummer 452881907 (grant CRC TRR 338).

E.R and F.J.T are funded by the European Union ERC, DeepCell - 101054957.

This research has been conducted using the UK Biobank Resource under application number [69938].

We thank Wellcome Sanger Institute scientific operations team members; Stefanie Lensing, Scott Thurston, Lesley Shirley, Sameena Iqbal, Katy Taylor, Cordelia Langford and Ian Johnston for automation, sample management and sequencing operations support. Wellcome Sanger Institute Human Genetics Department team members; Sally Bygraves, Nicola Corton, Kalia Dede, Guillaume Noel and Hannes Postingl.

Genes & Health is/has recently been core-funded by Wellcome (WT102627, WT210561), the Medical Research Council (UK) (M009017, MR/X009777/1, MR/X009920/1), Higher Education Funding Council for England Catalyst, Barts Charity (845/1796), Health Data Research UK (for London substantive site), and research delivery support from the NHS National Institute for Health Research Clinical Research Network (North Thames). We acknowledge the support of the National Institute for Health and Care Research Barts Biomedical Research Centre (NIHR203330); a delivery partnership of Barts Health NHS Trust, Queen Mary University of London, St George’s University Hospitals NHS Foundation Trust and St George’s University of London. Genes & Health is/has recently been funded by Alnylam Pharmaceuticals, Genomics PLC; and a Life Sciences Industry Consortium of AstraZeneca PLC, Bristol-Myers Squibb Company, GlaxoSmithKline Research and Development Limited, Maze Therapeutics Inc, Merck Sharp & Dohme LLC, Novo Nordisk A/S, Pfizer Inc, Takeda Development Centre Americas Inc. We thank Social Action for Health, Centre of The Cell, members of our Community Advisory Group, and staff who have recruited and collected data from volunteers.

We thank: Barts Health NHS Trust, NHS Clinical Commissioning Groups (City and Hackney, Waltham Forest, Tower Hamlets, Newham, Redbridge, Havering, Barking and Dagenham), East London NHS Foundation Trust, Bradford Teaching Hospitals NHS Foundation Trust, Public Health England, Discovery Data Service/Endeavour Health Charitable Trust (especially David Stables), Voror Health Technologies Ltd, NHS England - for GDPR-compliant data sharing backed by individual written informed consent.

Most of all we thank all of the volunteers participating in UK Biobank and Genes & Health.

## Author Contributions

G.I., D.v.S., E.L.C., F. Torabi., M.H., N. Subasinghe., L.D. and S.W. generated the data, with S.W. leading.

M.O., A.R., D.B., E.G., R.E., and V.I. performed data processing, with M.O., A.R., E.G. and V.I. co-leading.

E.G., S.W., M.O., A.C., and A.R. carried out data quality control and benchmarking, with M.O., A.R. and E.G. co-leading.

J.P.E., S.L., A.R., O.D. and F.C. conducted data integration, cell annotation and refinement, with F.C. leading.

M.O., A.L., S.S., A.R., A.S., B.A., E.H., M.J.B. and N.P. performed eQTL analyses, with M.J.B. and N.P. as co-leads. D.Z., S.W., N.P. and N.Soranzo analysed cell compositional traits. O.D., G.C., J.P.E., L.A., E.R., B.C., M.T., P.B., D.V., F.P.C., O.S., C.A.G. and N.Soranzo analysed cellular and gene programs. F.P.C. contributed to the design and interpretation of the variance decomposition analyses and co-supervised the corresponding analyses. E.G. performed factor analysis using tucker decomposition. A.R., performed ancestry analysis. P.B and B.C. performed pseudobulk differential gene expression analysis.

C.C., M.I.C., I.G., C.D.C., M.C., A.G., V.S., E.S. assisted with interpretation of results.

S.W., N.Staudt., V.I., S.A.T., M.J.B., O.S., D.A.v.H., N.P., and N.Soranzo designed and coordinated the project.

J.W., R.T., and S.F. provided research resources.

V.I., F.J.T., S.A.T., O.S., D.A.v.H., and N.Soranzo supervised the study.

E.G.*, O.D.*, A.R.*, J.P.E.*, M.T.*, D.Z.*, G.C.*, S.W.*, M.O.*, F.C., F.T., B.C., O.S., V.I., M.J.B., D.A.v.H.^#^, S.A.T.^#^, N.P.^#^ and N.Soranzo^#^ wrote the manuscript.

E.G.*, O.D.*, A.R.*, J.P.E.*, M.T.*, D.Z.*, G.C.*, S.W.*, M.O.* contributed equally to this work.

All authors reviewed and approved the final manuscript.

## Consortia

### Genes & Health Research Team

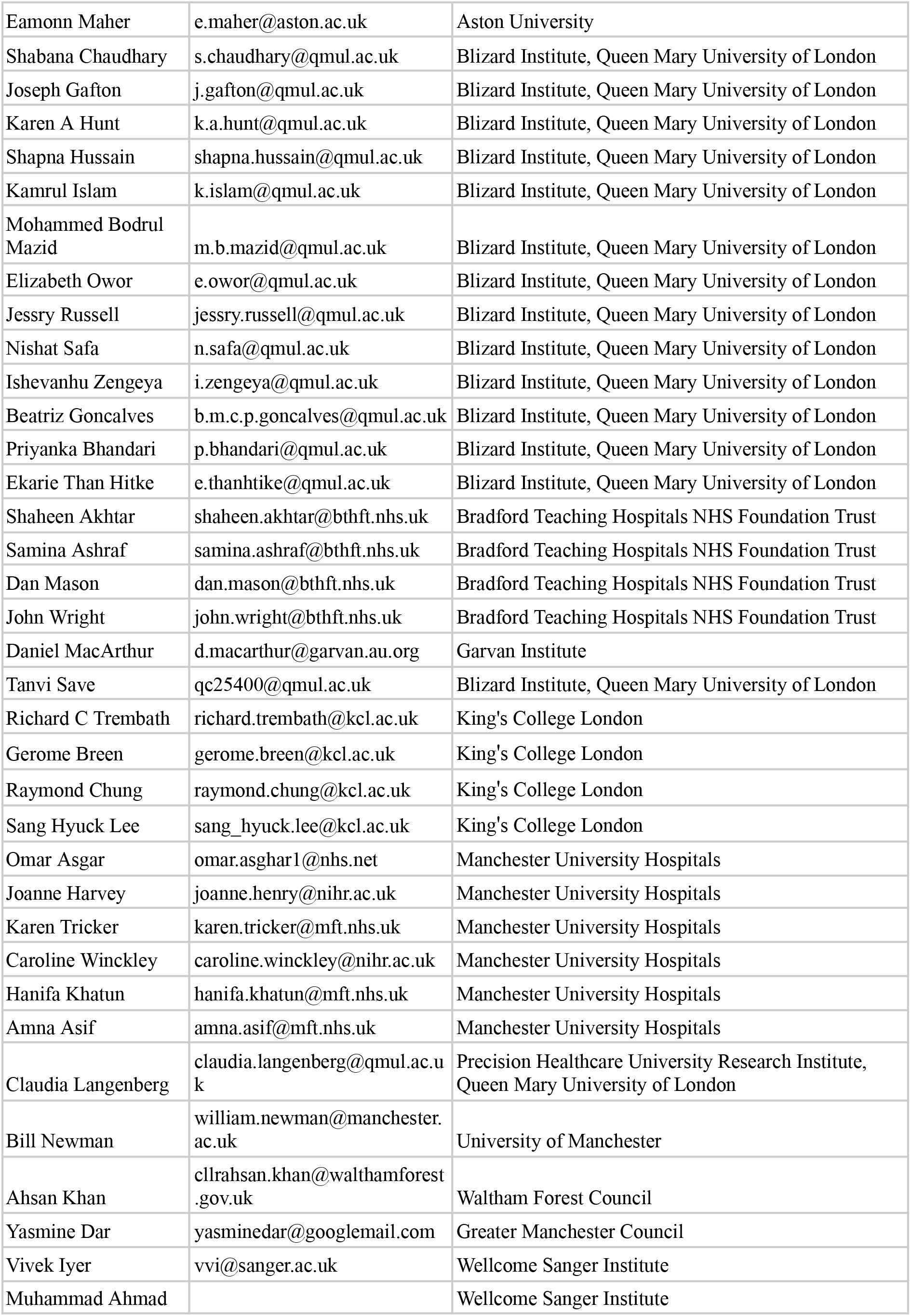
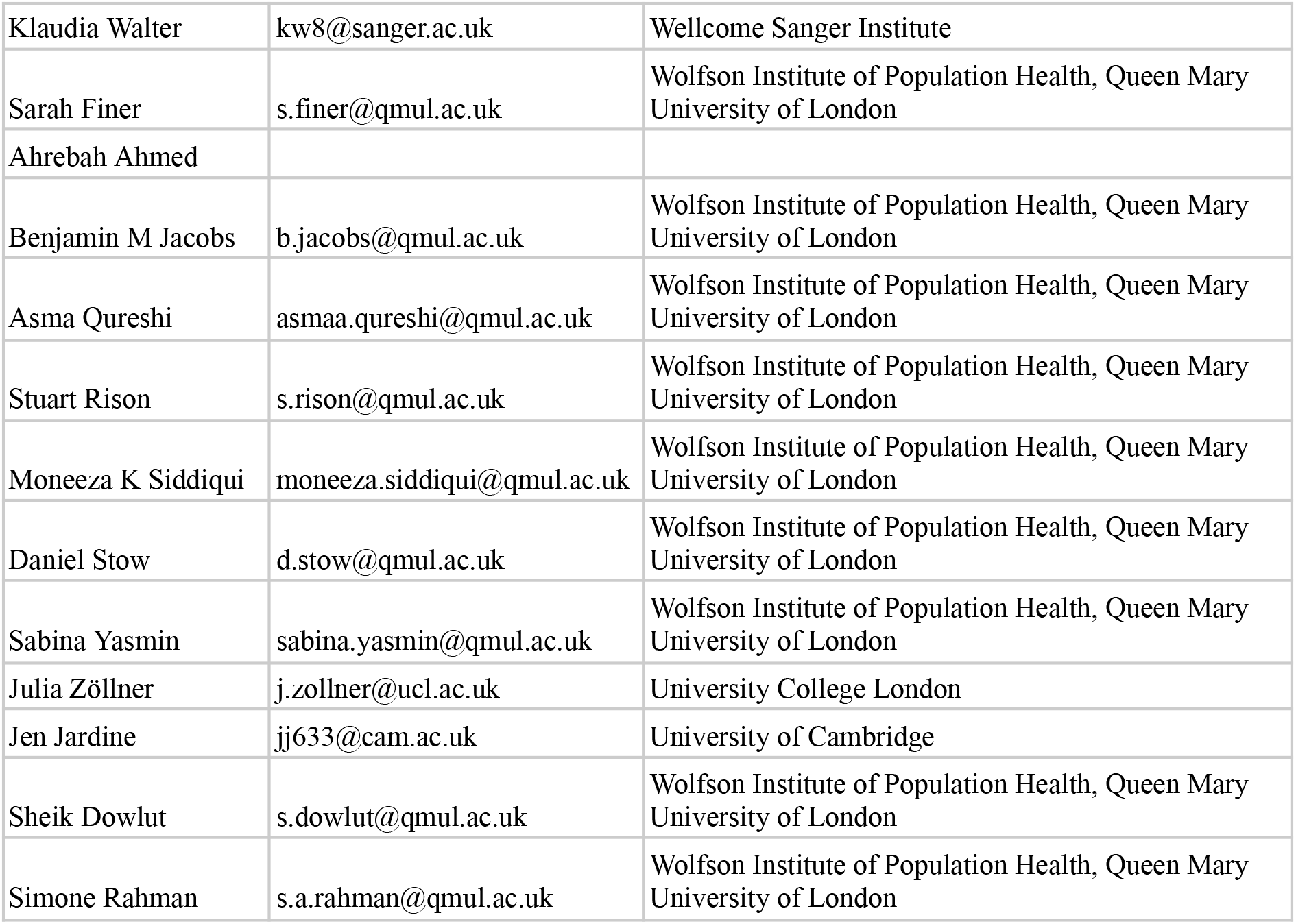

## Additional Information

Supplementary Information is available for this paper.

## Data availability

The GWAS summary statistics for the 29 hematological traits from UKB performed in this study are available in Zenodo (10.5281/zenodo.21723758). Single-cell eQTLs, fine-mapped eQTL credible sets, eQTL modules and colocalisation results generated using FLANDERS pipeline on the CD16+ NK adaptive cells are available in in Zenodo (10.5281/zenodo.21723758). The scANVI model pre-trained on the CARDINAL dataset is available in the CARDINAL data central repository (data will be released upon publication) and can be used for label transfer in new datasets. The Celltypist cell type prediction models for celltype 2 and celltype 3 labels are available in the CARDINAL data central repository (data will be released upon publication). Single-cell eQTLs, fine-mapped eQTL credible sets and eQTL modules generated using FLANDERS pipeline are available in the CARDINAL data central repository (data will be released upon publication). Full results for the EHR vs genetics variance decomposition and inferred latent programs are available in the CARDINAL data central repository (data will be released upon publication). Single-cell complete blood counts (scCBCs) for CARDINAL samples are available to bona fide researchers upon approved application to the UK Biobank Access Management System and Genes & Health Trusted Research Environment. The summary statistics for associations between the scCBCs and quantitative / disease traits are available in the CARDINAL data central repository (data will be released upon publication). Single cell attention values based on MixMIL analysis are available in the CARDINAL data central repository (data will be released upon publication). The gnomAD variant allele-frequencies annotations are available from gnomAD dataset portal v4.0 (https://gnomad.broadinstitute.org/). GREEN-DB v2.5 data on human regulatory regions was downloaded from Zenodo (doi: 10.5281/zenodo.5636209). The genotype and phenotype for the UKB cohort described in this study were obtained from the UK Biobank resource under approved application number 69938. These are available, together with raw and processed single-cell data, to bona fide researchers upon approved application to the UK Biobank Access Management System. The health and genetic data for the G&H cohort used in this study were obtained from the Genes & Health resource. These are available, together with raw and processed single-cell data, to registered researchers via the secure Genes & Health Trusted Research Environment following approval by the cohort’s data access committee. Fine-mapped credible sets for FinnGEN, GTEx, GWAS Catalog datasets were downloaded from OpenTarget Genetics v25.09 (https://platform.opentargets.org/).

## Code availability

The YASCP pipeline used to process 10X single-cell data and perform donor deconvolution is available on GitHub (https://github.com/wtsi-hgi/yascp). The NADA pipeline used to perform single-cell count normalization and aggregation is available on GitHub (https://github.com/davidebolo1993/nada_smk). The QTLight pipeline used to compute single-cell eQTLs is available in GitHub (https://github.com/wtsi-hgi/QTLight). Code for the FLANDERS workflow used to perform fine-mapping, to create cis-genetic modules and perform colocalisation is available in GitHub (https://github.com/Biostatistics-Unit-HT/Flanders). The cellink package is available on GitHub (https://github.com/theislab/cellink). A copy of the code used for specific analyses described in this paper is available in Zenodo (10.5281/zenodo.21723758).

