## Extended data figures 1-10 for "Single-cell dissection of immune-state architecture across diverse human population cohorts"

23 October 2026

### List of Figures


| | | |
|---|---|---|
| Extended Data Fig. 1: | Cell abundance by population | 2 |
| Extended Data Fig. 2: | Effects of infectious agents on cell composition | 3 |
| Extended Data Fig. 3: | Gene signatures from differentially expressed genes (DEGs) | 4 |
| Extended Data Fig. 4: | Decomposition of genetic and electronic health record (eHR) contributions to population-specific gene regulation across immune cell types | 5 |
| Extended Data Fig. 5: | Association of genes to eHR traits and exposures in G&H cohort | 6 |
| Extended Data Fig. 6: | Phenome-wide association study (PheWAS) plots illustrating cell type–specific disease associations across eHR traits in the G&H and UKB cohorts | 7 |
| Extended Data Fig. 7: | Mendelian randomisation of BMI on gene expression | 8 |
| Extended Data Fig. 8: | Variance decomposition programmes - Extended coupling | 9 |
| Extended Data Fig. 9: | Overview of associations for the top genes contributing to gene programs 2 and 3 | 10 |
| Extended Data Fig. 10: | eQTL signals identified across cell types | 11 |

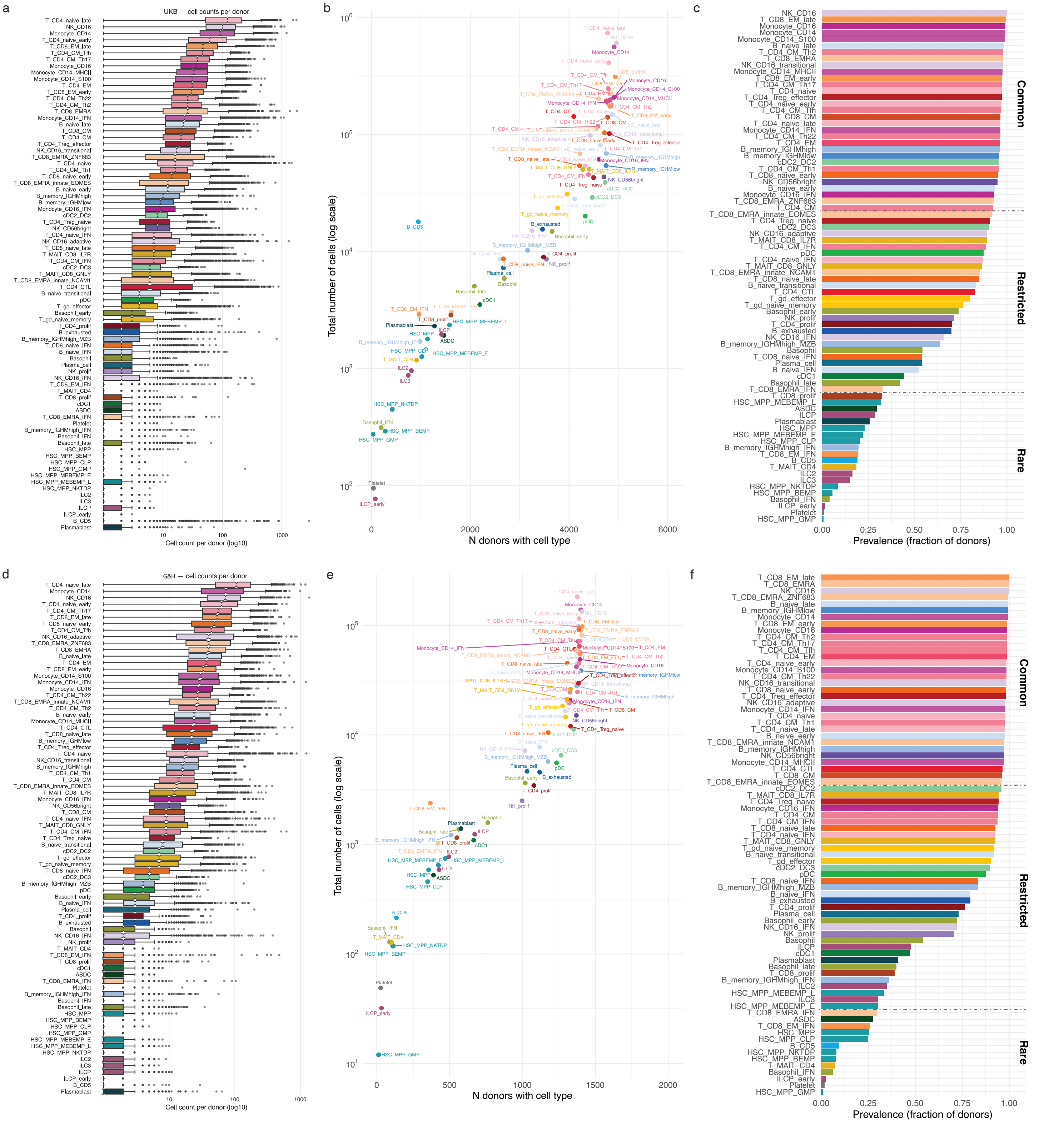


**Extended Data Fig 1: Cell abundance by population**

**a**, Distribution of number of cells per donor at CT3 level for the UKB cohort. **b**, Relative distribution of cell abundance vs number of donors within cell types for the UKB cohort. **c**, Cell type classification based on prevalence by donor for the UKB cohort. **d-f**, Same plots are reported for the G&H cohort.

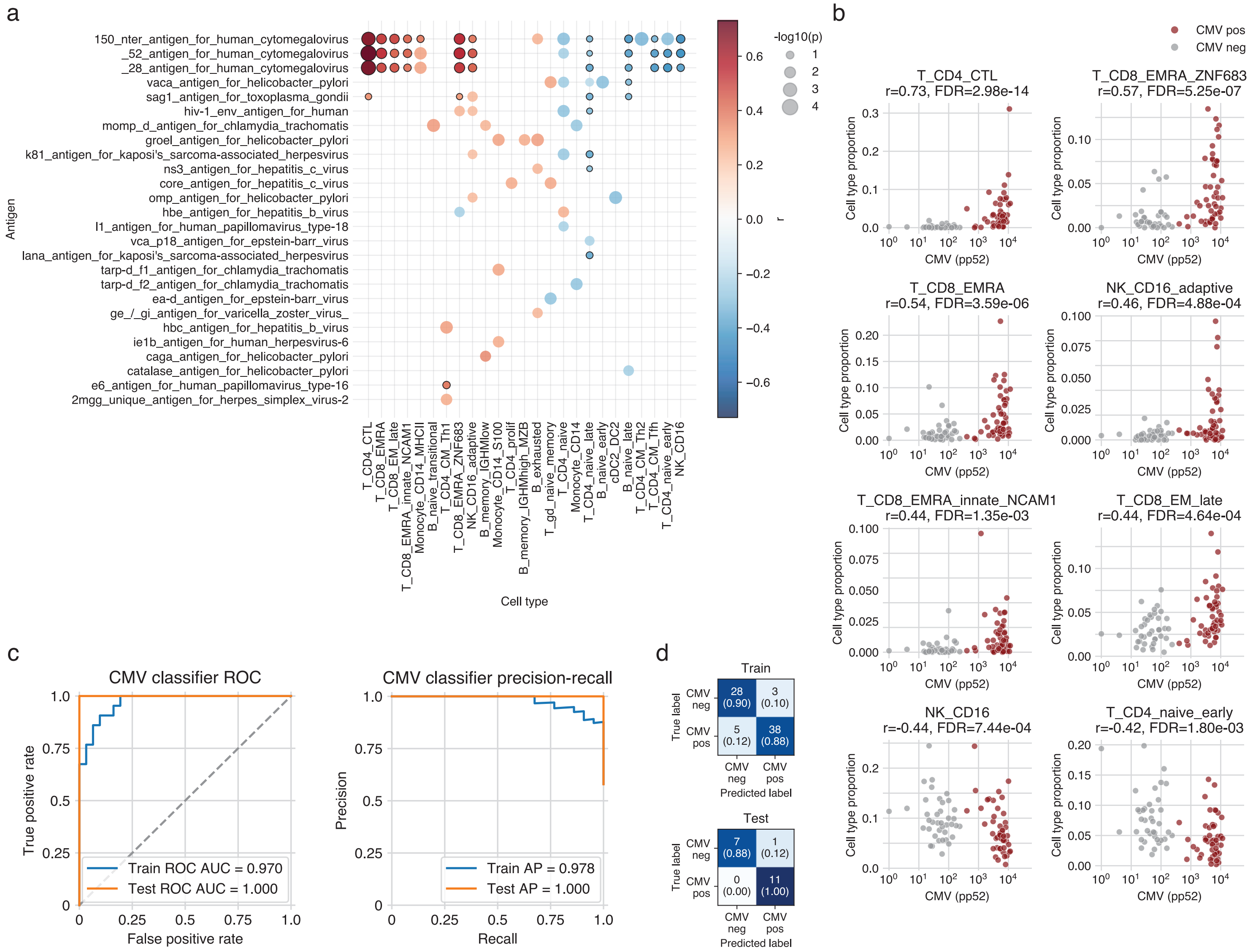


**Extended Data Fig 2: Effects of infectious agents on cell composition**

**a**, Correlation of cell proportions (CT3 level) with antigen serology in UK Biobank (n=90 donors). Pairs with FDR $< 0.2$ are shown and those with FDR $< 0.05$ are marked with a black border. **b**, Scatter plots of cell proportions (CT3 level) with antibody reactivity for CMV pp52 antigen for cell types with strongest correlations. CMV status inferred from serology data is shown as colour. **c**, Performance metrics (ROC AUC and precision-recall curve) for cell composition-based random forest classifier for CMV serostatus. **d**, Confusion matrices of the CMV classifications for train and test datasets.

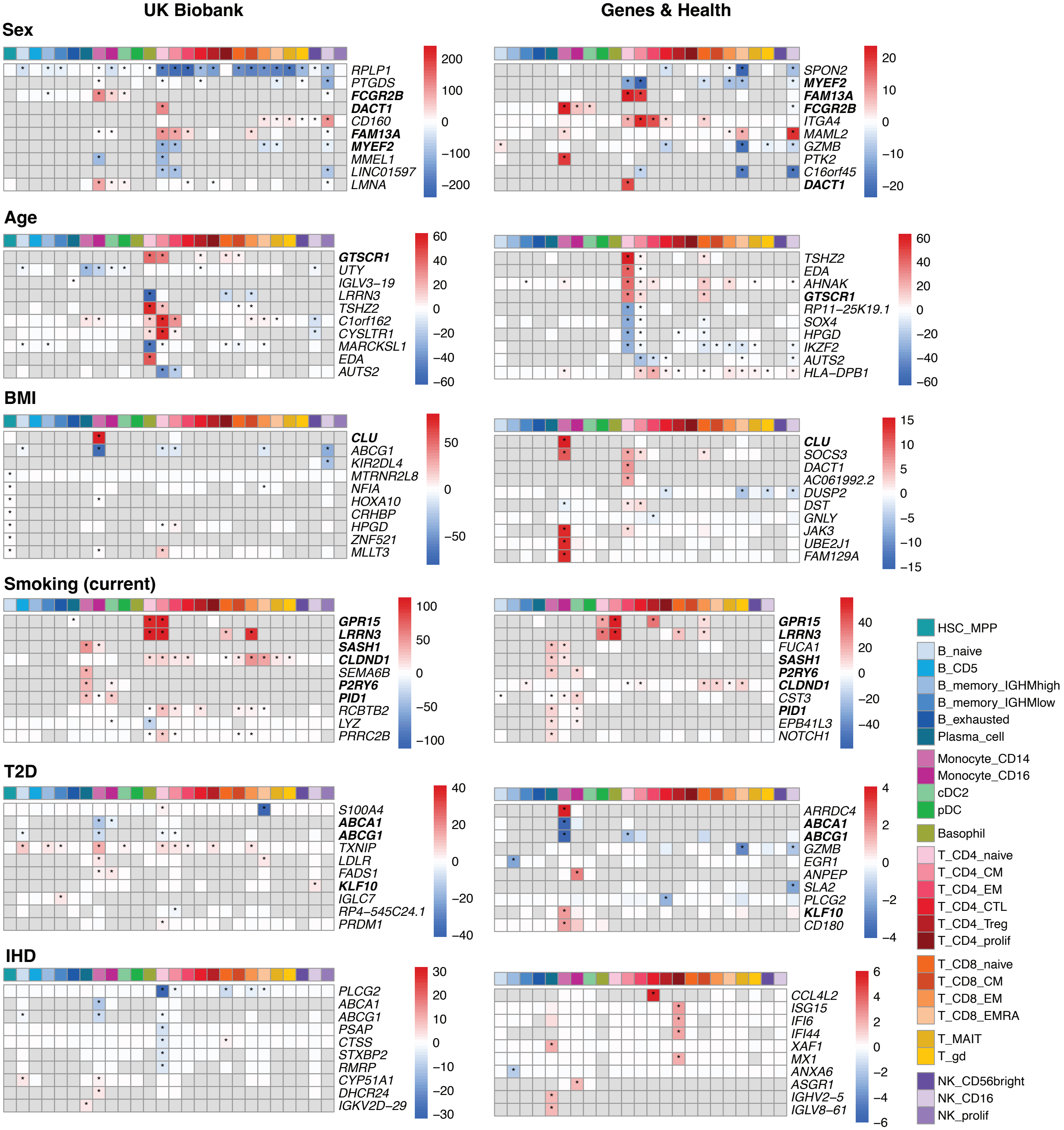


**Extended Data Fig 3: Gene signatures from differentially expressed genes (DEGs)**
Heatmaps showing for each trait - sex, age, BMI, current smoking, type 2 diabetes (T2D) and ischaemic heart disease (IHD) - and cohort the top 10 DEGs across all cell types. Values represent the $-log_{10}$ BH-adjusted P value, signed by log fold change. Asterisks denote BH-adjusted P value $<$ 0.05. Genes in bold are shared between cohorts as top 10 genes.

a

| Cell type | N samples | N genes | N cells |
|---|---|---|---|
| Monocytes | 4,436 | 18,480 | 1,324,151 |
| T CD8 | 4,394 | 18,042 | 1,295,243 |
| T CD4 | 4,387 | 19,565 | 2,684,422 |
| NK | 4,367 | 16,914 | 801,581 |
| B | 4,237 | 15,342 | 386,680 |
| DC | 3,499 | 14,065 | 69,262 |
| T MAIT | 2,650 | 11,883 | 59,290 |
| T $\gamma\delta$ | 1,590 | 10,731 | 20,055 |

b

| Cell type | N samples | N genes | N cells |
|---|---|---|---|
| Monocytes | 1,221 | 18,904 | 377,863 |
| T CD8 | 1,221 | 19,544 | 572,868 |
| T CD4 | 1,213 | 20,377 | 861,359 |
| NK | 1,208 | 17,714 | 276,906 |
| B | 1,198 | 17,290 | 211,074 |
| DC | 913 | 14,756 | 18,471 |
| T MAIT | 890 | 13,761 | 34,758 |
| T $\gamma\delta$ | 763 | 13,071 | 20,813 |

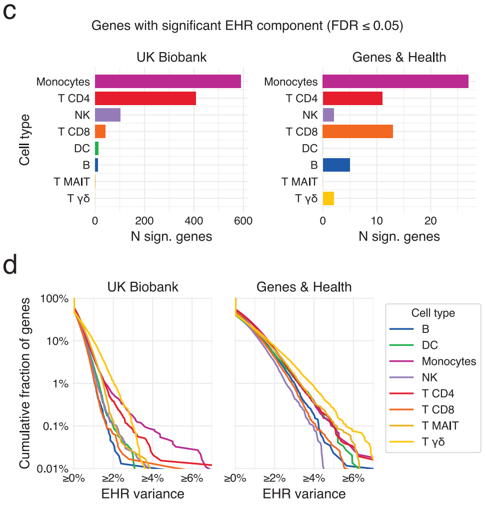


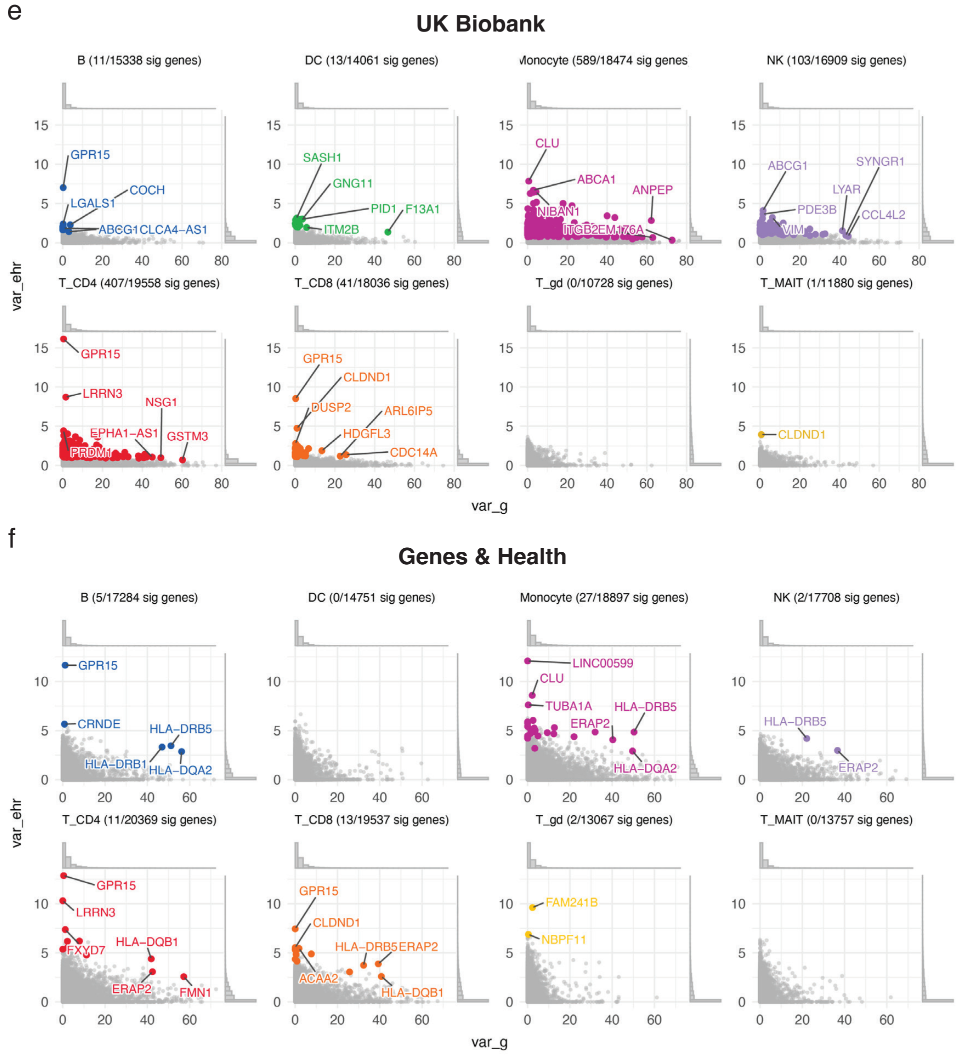


**Extended Data Fig 4: Decomposition of genetic and electronic health record (eHR) contributions to population-specific gene regulation across immune cell types**
**a**, **b**, Summary of pseudobulk single-cell datasets used for variance decomposition analyses in the two cohorts. Shown are the number of donors, genes tested, and cells profiled for each of the eight major immune cell types (Monocytes, CD8 T cells, CD4 T cells, NK cells, B cells, dendritic cells (DC), MAIT T cells, and $\gamma\delta$ T cells) in the UK Biobank (a) and Genes & Health (b) cohorts. **c**, Number of genes with a significant eHR-associated variance component (empirical FDR $\leq$ 0.05) across cell types in UK Biobank (left) and Genes & Health (right). Monocytes exhibited the largest number of eHR-associated genes in both cohorts. **d**, Cumulative fraction of genes with at least a given proportion of gene expression variance explained by EHR effects across cell types in UK Biobank (left) and Genes & Health (right). Each coloured curve represents one immune cell type. **e**, **f**, Gene-level variance decomposition results for each immune lineage in UK Biobank (e) and Genes & Health (f). Scatterplots show the proportion of expression variance explained by cis-genetic effects (x axis; var_g) versus EHR-derived effects (y axis; var_ehr) for all tested genes within each cell type. Genes with significant EHR variance components (empirical FDR $\leq$ 0.05) are highlighted and labelled. Titles indicate the number of significant genes relative to the total number tested for each lineage. Histograms along the axes show the marginal distributions of genetic and EHR variance components (log-scale). While cis-genetic effects explained a larger fraction of transcriptomic variability overall, a subset of genes displayed substantial EHR-associated variance, often in a cell-type- and cohort-specific manner. Representative examples include *CLU*, *ABCA1*, and *ANPEP* in monocytes from UK Biobank, and *HLA-DRB5*, *ERAP2*, and *LINC00599* in monocytes from Genes & Health.

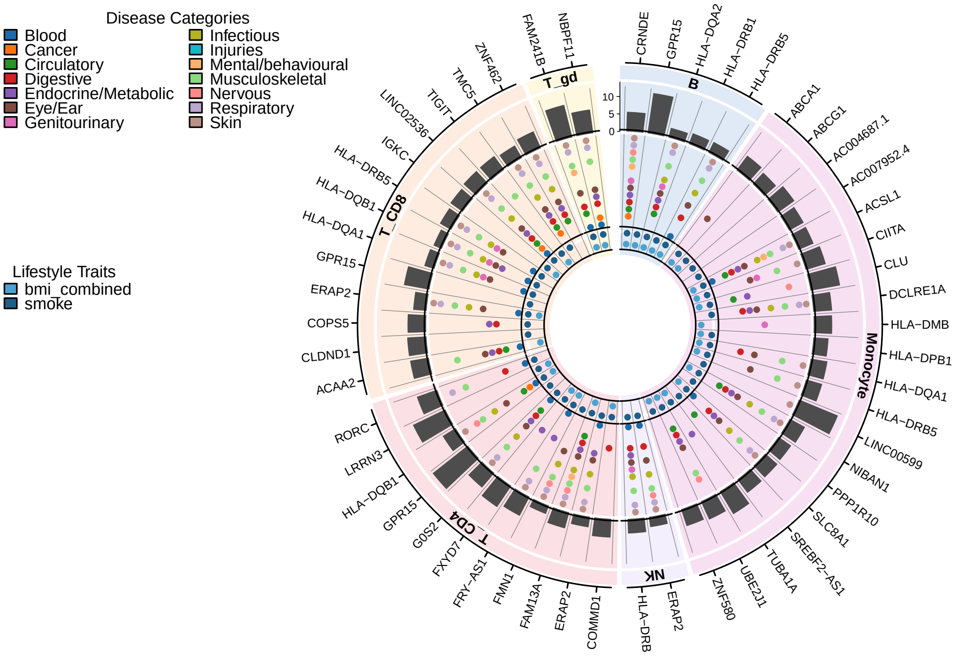


**Extended Data Fig 5: Association of genes to eHR traits and exposures in G&H cohort**

Circos plot summarizing, for the G&H cohort, the associations between gene expression and exposures (BMI, smoking) or EHR terms, as determined by variance decomposition analysis. Only the top 20 genes with Pearson's r $\geq$ 0.2 and the largest EHR-explained variance are shown. The outer ring represents the fraction of gene expression variance explained by the EHR component. The two inner rings represent significant signals for diseases and lifestyle traits (BMI, smoking), respectively.

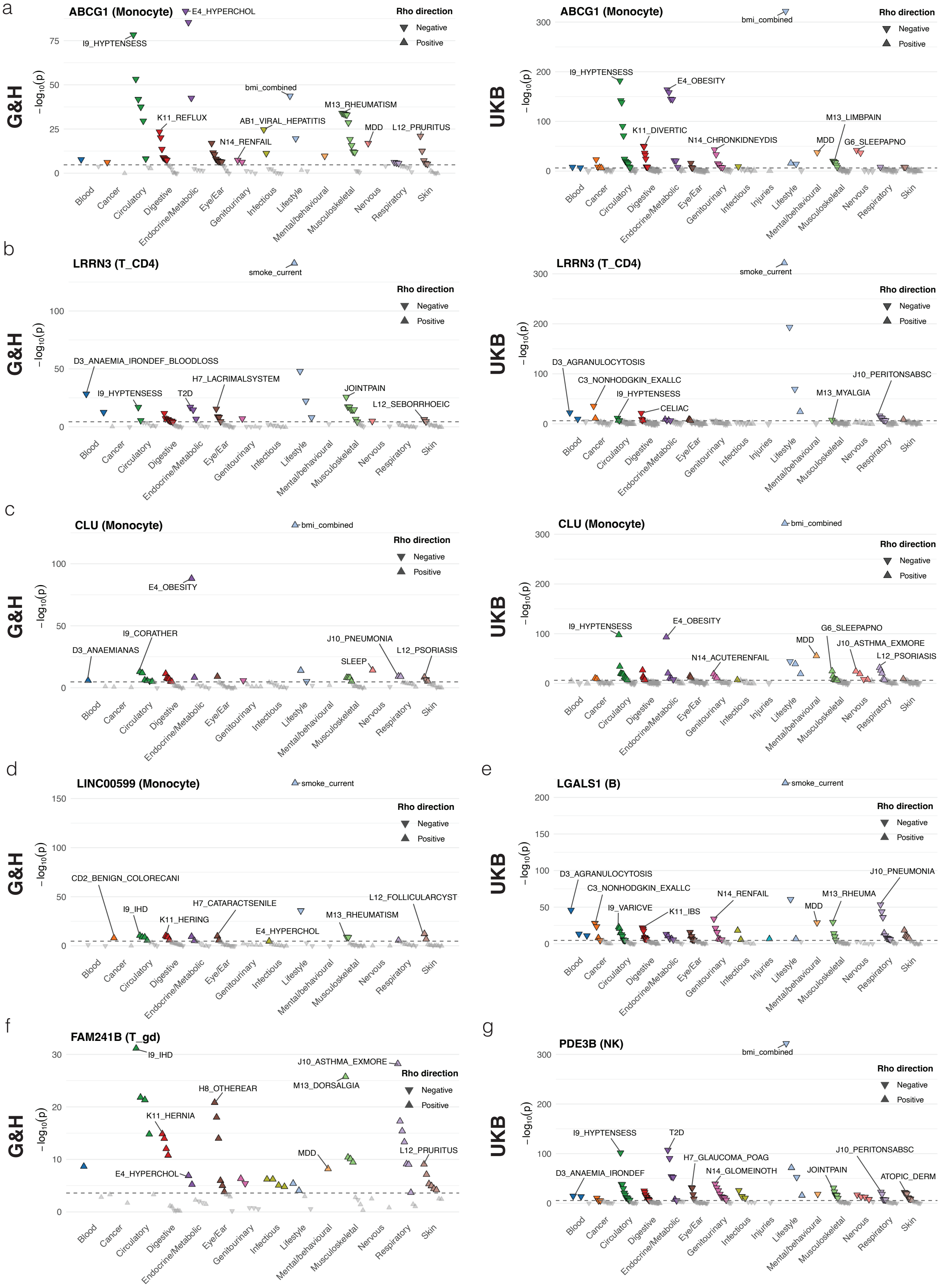


**Extended Data Fig 6: Phenome-wide association study (PheWAS) plots illustrating cell type–specific disease associations across eHR traits in the G&H and UKB cohorts**

Panels show PheWAS results for representative genes and cell types: (a) *ABCG1* (Monocyte), (b) *LRRN3* ($CD4^{+}$ T cells), (c) *CLU* (Monocyte), (d) *LINC00599* (Monocyte), (e) *LGALS1* (B), (f) *FAM241B* ($\gamma\delta$ T cells), and (g) *PDE3B* (NK cells). Left and right columns correspond to the G&H and UKB cohorts, respectively. Each point represents the association between the expression component predicted from EHR and a disease phenotype grouped by clinical category along the x-axis. The y-axis shows the association significance as $-\log_{10}(p)$. Upward and downward triangles indicate positive and negative effect directions (Pearson's r), respectively. Points above the significance threshold are colored and selected top phenotypes are annotated. Dashed horizontal lines indicate the significance cutoff after multiple-testing correction.

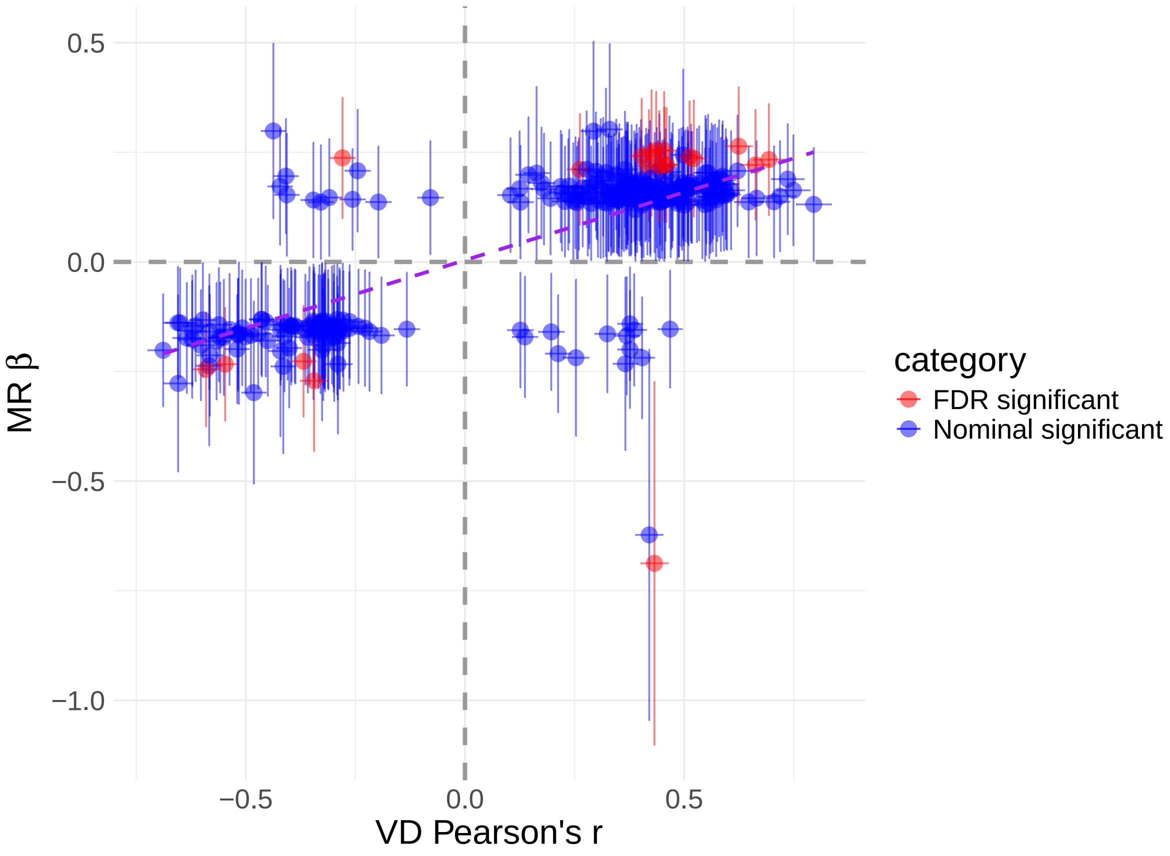


**Extended Data Fig 7: Mendelian randomisation of BMI on gene expression**
Mendelian randomisation analysis of BMI-associated gene–cell type pairs. The scatter plot compares the Pearson correlation coefficient from variance decomposition (VD Pearson's r) with the MR effect estimate (MR $\beta$), across 956 gene–cell type pairs. Red points indicate associations significant at FDR $< 0.05$, whereas blue points indicate nominally significant associations ($P < 0.05$). Error bars represent 95% confidence intervals.

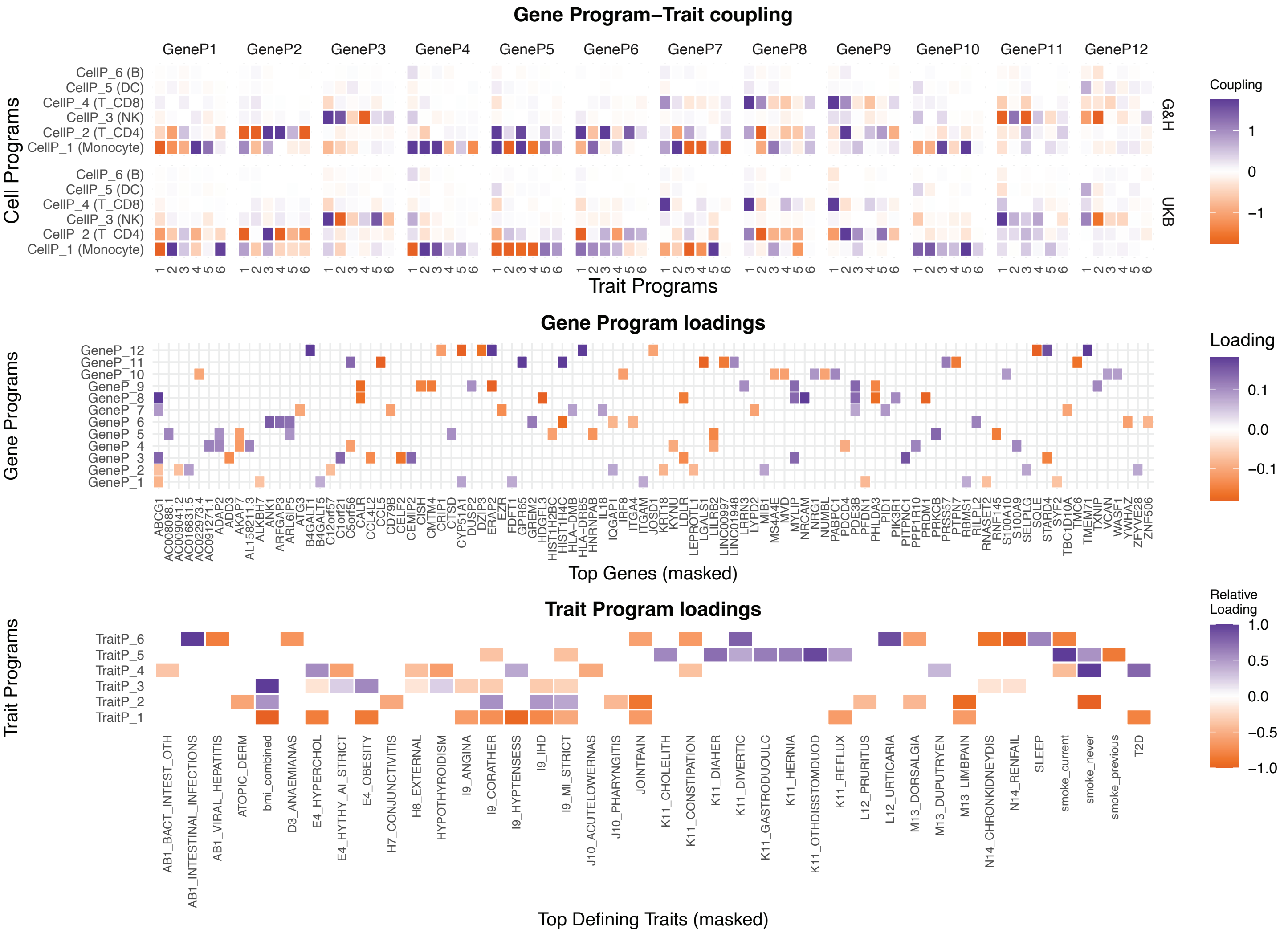


**Extended Data Fig 8: Variance decomposition programmes - Extended coupling**

Gene programs coupling with traits. Heatmap on the top row shows the coupling of gene programs with trait programs across the celltype programs. The signs are aligned to the dominant celltype in each celltype program. The loading matrix for the gene program and the trait program are reported below, showing the top 10 and the top 12 contributors, respectively. Genes / traits not in the top contributors are masked in each gene / trait program.

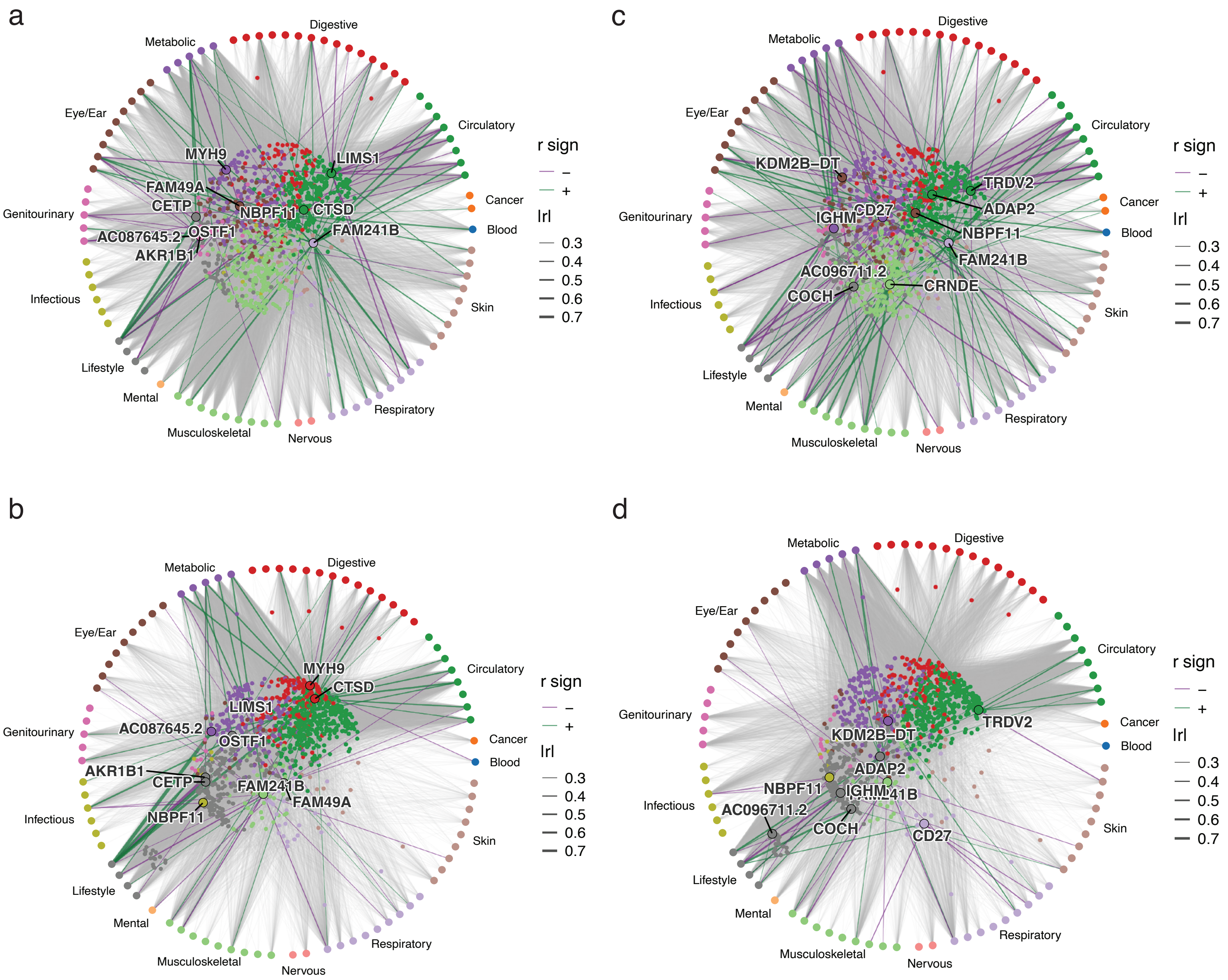


**Extended Data Fig 9: Overview of associations for the top genes contributing to gene programs 2 and 3**

**a**, **b**, The networks represent the pleiotropic connections for the top 10 genes in Gene Program 3 across all tested traits for G&H (a) and UKB (b). **c**, **d**, Similar networks for the Gene Program 2 for G&H (c) and UKB (d) cohort. The color of the lines indicates the direction of association, while the line width is proportional to Pearson's r value. The color of the gene dots represents the main trait category they are associated with, based on the number and strength of single trait associations.

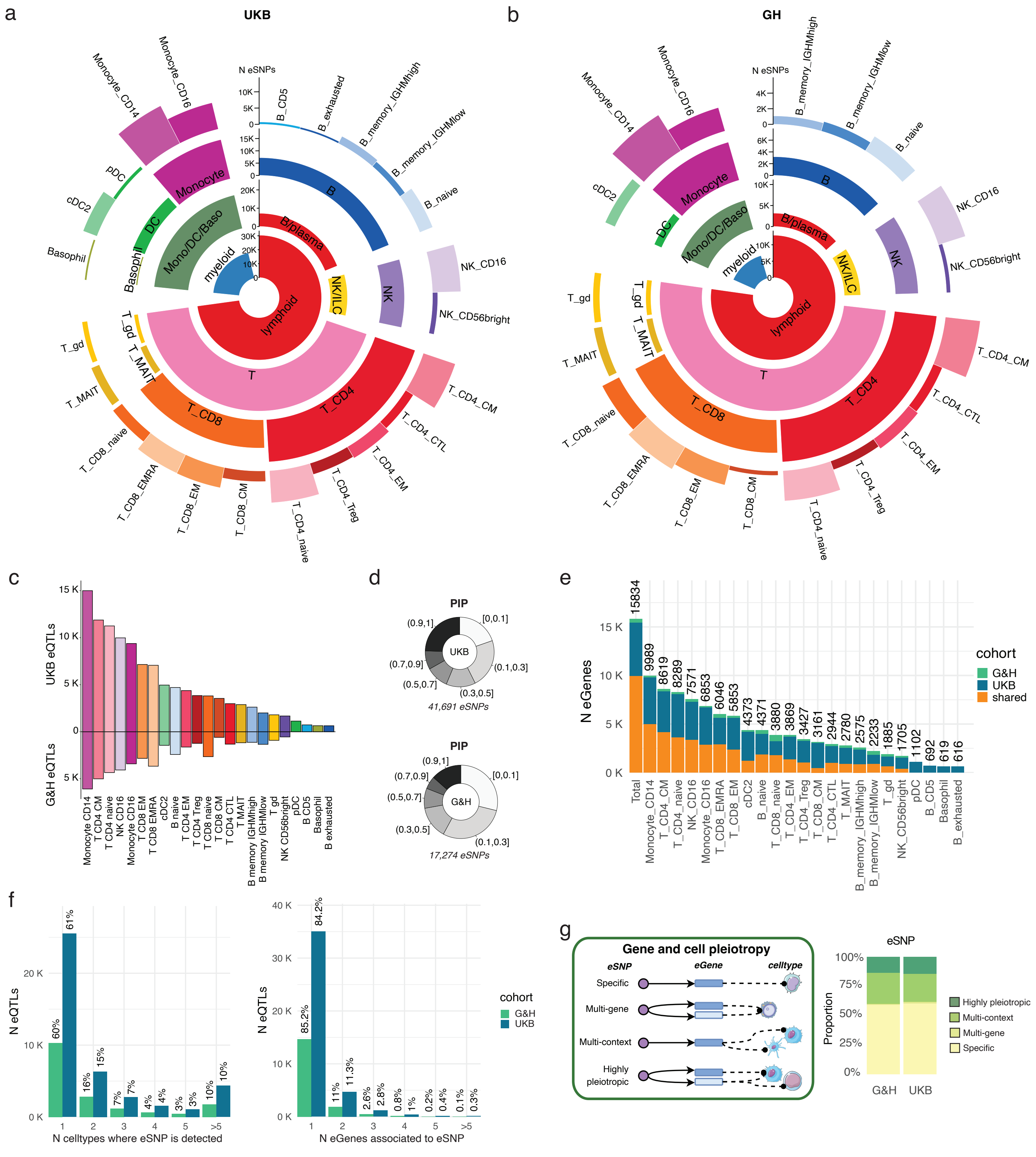


**Extended Data Fig 10: eQTL signals identified across cell types**

**a, b**, Number of independent eSNP signals identified by FLANDERS framework across cell types for the UKB (a) and G&H (b) cohort. The circos layers visualize counts across the 4 cell type granularities, ordered from inner (broad) to outer (specific): lineage 1, lineage 2, cell type 1 (CT1) and cell type 2 (CT2). **c**, Number of fine-mapped signals discovered in the UKB and G&H cohorts across CT2 cell types. d, Distribution of PIP values across eSNPs as determined by FLANDERS pipeline in the UKB and G&H cohorts. **e**, Distribution of eGenes discovered in the 2 cohorts, reporting the number of cohort-specific and cohort-shared eGenes across CT2 cell types. f, Distribution of eSNP signals in terms of number of cell types and number of associated eGenes. **g**, Diagram summarising the pleiotropy classification used for eSNPs in the study, based on their action across genes and cell types. The barplot reports the distribution of the eSNPs across the pleiotropy categories in the G&H and UKB cohort.
