## Supplementary Figures 1-20, Supplementary Tables Legends for "Single-cell dissection of immune-state architecture across diverse human population cohorts"

### Supplementary Information

26 October 2026

This document contains supplementary information associated with the manuscript:
*"Single-cell dissection of immune-state architecture across diverse human population cohorts"*

### Supplementary Figures


| | | |
|---|---|---|
| Supplementary Fig. 1: | Cardinal experimental and data processing workflow | 4 |
| Supplementary Fig. 2: | Pool-level quality control | 5 |
| Supplementary Fig. 3: | Integration metrics | 6 |
| Supplementary Fig. 4: | Cell type 2 marker genes | 7 |
| Supplementary Fig. 5: | Marker genes and UMAPs for CT3 cell states | 8 |
| Supplementary Fig. 6: | Single-cell Complete Blood Counts | 9 |
| Supplementary Fig. 7: | MixMIL attention weights for selected programmes | 10 |
| Supplementary Fig. 8: | Cross-cohort heatmap of cell type–specific disease-associated differential expression signatures | 11 |
| Supplementary Fig. 9: | Benchmarking and robustness assessment of variance decomposition analyses across cohorts and immune cell types | 12 |
| Supplementary Fig. 10: | Variance decomposition effects matrix for UKB Biobank | 13 |
| Supplementary Fig. 11: | Variance decomposition effects matrix for Genes&Health | 14 |
| Supplementary Fig. 12: | Tucker decomposition loadings matrix | 15 |
| Supplementary Fig. 13: | GSEA analysis results for gene programs inferred from Tucker decomposition | 16 |
| Supplementary Fig. 14: | Main factors influencing eQTL discovery power | 17 |
| Supplementary Fig. 15: | Characterization of eSNPs | 18 |
| Supplementary Fig. 16: | Cross-population comparison of eQTL effect sizes, allele frequencies, and gene expression for all the subsets of variants highlighted in Figure 5. | 19 |
| Supplementary Fig. 17: | Characteristics and examples of contextual eQTLs F.E. $\geq$ 1.1 | 20 |
| Supplementary Fig. 18: | Mendelian randomisation in NK_CD16_adaptive. | 21 |
| Supplementary Fig. 19: | Cell-level quality control | 22 |
| Supplementary Fig. 20: | Optimal ranks for Tucker decomposition | 23 |

#### Supplementary Table Legends


| | | |
|---|---|---|
| Supplementary Table 1: | Sample demographics summary on the QCed dataset | 24 |
| Supplementary Table 2: | Medical record prevalence on the restricted dataset | 24 |
| Supplementary Table 3: | Pool and donor information for RNA sequencing samples | 24 |
| Supplementary Table 4: | Cell annotation gene markers | 24 |
| Supplementary Table 5: | Number of cells and number of samples per cell type | 24 |
| Supplementary Table 6: | Summary of donors and cells used in analyses | 24 |
| Supplementary Table 7: | Compositional signatures for UK Biobank and Genes & Health | 24 |
| Supplementary Table 8: | MixMil signatures for UK Biobank and Genes & Health | 24 |
| Supplementary Table 9: | Variance decomposition – genes with significant EHR component (UK Biobank) | 24 |
| Supplementary Table 10: | Variance decomposition – genes with significant EHR component (Genes & Health) | 24 |
| Supplementary Table 11: | Variance decomposition – gene to trait significant correlations for EHR terms (UK Biobank) | 24 |
| Supplementary Table 12: | Variance decomposition – gene to trait significant correlations for EHR terms (Genes & Health) | 25 |
| Supplementary Table 13: | MR results for BMI | 25 |
| Supplementary Table 14: | Gene loadings from the VD latent factors | 25 |
| Supplementary Table 15: | Trait loadings from the VD latent factors | 25 |
| Supplementary Table 16: | Celltype loadings from the VD latent factors | 25 |
| Supplementary Table 17: | GSEA results on gene loadings – GOBP | 25 |
| Supplementary Table 18: | GSEA results on gene loadings – REACTOME | 25 |
| Supplementary Table 19: | GSEA results on gene loadings – Human Hallmark | 25 |
| Supplementary Table 20: | GSEA results on gene loadings – ImmuneSigDB | 25 |
| Supplementary Table 21: | eGene discovery across tested cell types | 25 |
| Supplementary Table 22: | Pleiotropy classification of independent causal genetic variants (eSNPs) in UKB and G&H | 25 |
| Supplementary Table 23: | Summary of population-shared, cohort-specific, and contextual eQTLs for the UK Biobank cohort | 25 |
| Supplementary Table 24: | Summary of population-shared, cohort-specific, and contextual eQTLs for the G&H cohort | 25 |
| Supplementary Table 25: | Credible sets for eQTL identified in CD16+ adaptive NK, cell-type specificity and colocalisation results | 25 |
| Supplementary Table 26: | Colocalisation and Mendelian randomisation results for CD16+ adaptive NK cell-specific eQTL signals with trait and disease associations | 26 |
| Supplementary Table 27: | Summary of differential gene expression analysis for Genes & Health at CT2 | 26 |
| Supplementary Table 28: | Summary of differential gene expression analysis for UK Biobank at CT2 | 26 |
| Supplementary Table 29: | Colocalisation of co-regulated gene expression modules with traits and diseases in G&H cohort | 26 |
| Supplementary Table 30: | Data sources for GWAS summary-statistics for the 46 manually curated disease datasets | 26 |

a

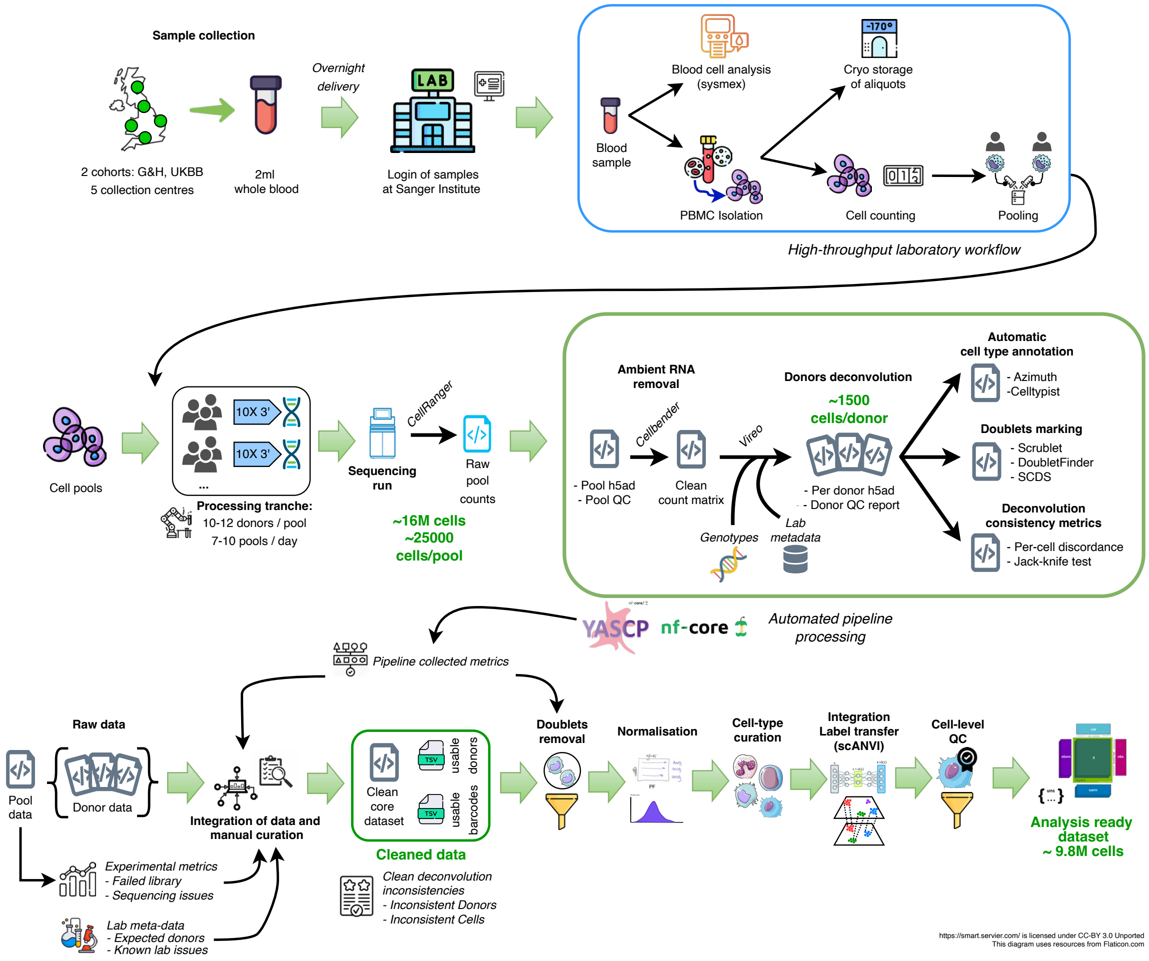


b

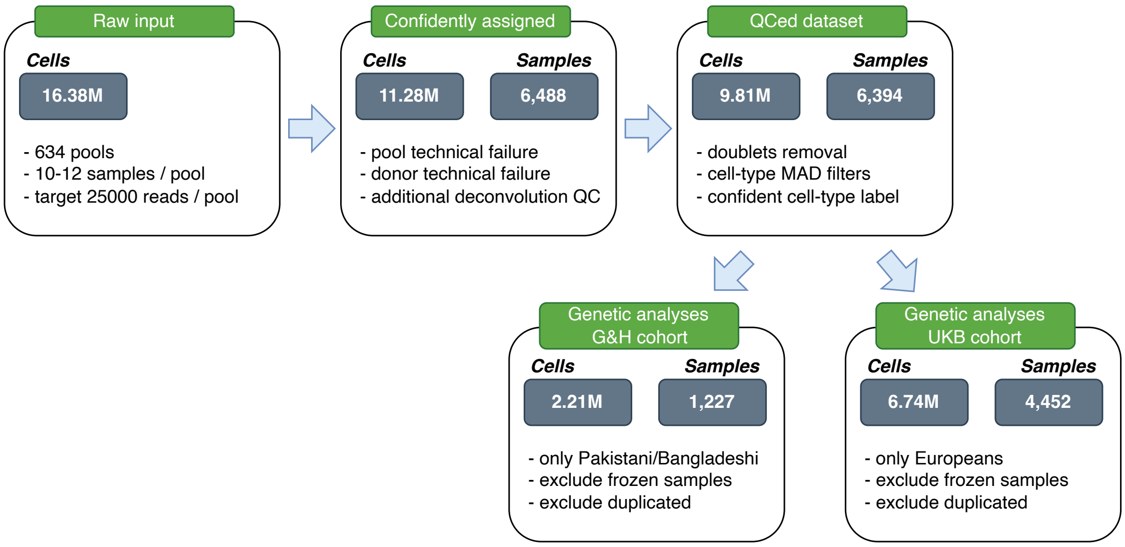


**Supplementary Fig. 1: Cardinal experimental and data processing workflow**
**a**, Overview of sample recruitment, data preparation, and processing steps. **b**, Flowchart illustrating the number of cells and individuals after each processing and QC step. Raw data from each sequencing pool was separated into single samples using genotype-based deconvolution (confidently assigned cells), and low-quality cells were then removed using an adaptive filtering strategy to generate the QCed dataset. Finally, the QCed cell dataset was split between the Genes&Health (G&H) and UK Biobank (UKB) cohorts, and the 2 groups were considered separately in subsequent analyses.

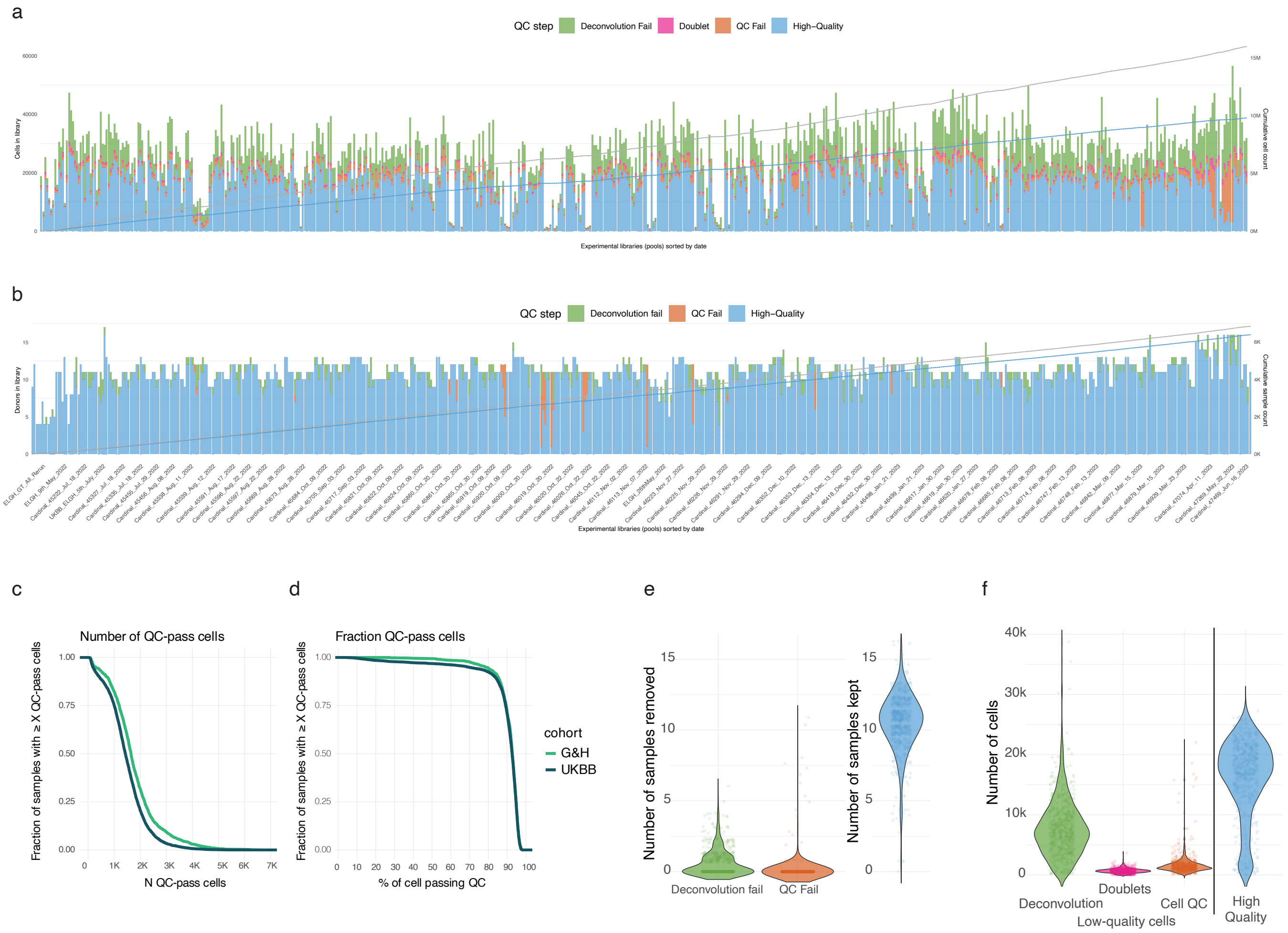


**Supplementary Fig. 2: Pool level quality control**

**a**, Number of cells per pool, with pools on the x-axis sorted by collection date. Bars represent cell counts per pool, and the overlaid cumulative curve indicates the total number of cells accumulating through time. **b**, Number of donors per pool, with pools on the x-axis sorted by collection date. Bars represent donor counts per pool, and the cumulative curve indicates the total number of donors accumulating through time. **c, d**, Cumulative distribution of the number and fraction of high-quality (QC-pass) cells per sample in G&H and UKB cohorts. **e, f**, Distribution of the number of samples and number of cells discarded and kept per pool across the major filtering steps.

a

| Method | Bio conservation: KMeans NMI | Bio conservation: KMeans ARI | Bio conservation: Silhouette label | Batch correction: Silhouette batch | Batch correction: iLISI | Batch correction: KBET | Batch correction: Graph connectivity | Batch correction: PCR comparison | Aggregate score: Batch correction | Aggregate score: Bio conservation | Aggregate score: Total |
|---|---|---|---|---|---|---|---|---|---|---|---|
| **scanvi_50** | 0.68 | 0.45 | 0.61 | 0.87 | 0.16 | 0.74 | 0.94 | 0.51 | 0.64 | 0.58 | 0.61 |
| **scanvi_50_esvl** | 0.65 | 0.43 | 0.60 | 0.87 | 0.16 | 0.74 | 0.94 | 0.53 | 0.65 | 0.56 | 0.60 |
| **harmony** | 0.62 | 0.37 | 0.56 | 0.91 | 0.15 | 0.82 | 0.77 | 0.93 | 0.72 | 0.52 | 0.60 |
| **scpoli** | 0.63 | 0.40 | 0.62 | 0.80 | 0.16 | 0.73 | 0.96 | 0.62 | 0.65 | 0.55 | 0.59 |
| **scanvi_large** | 0.63 | 0.35 | 0.60 | 0.88 | 0.15 | 0.73 | 0.95 | 0.43 | 0.63 | 0.53 | 0.57 |
| **scanvi_large_es_lb** | 0.63 | 0.35 | 0.60 | 0.88 | 0.15 | 0.73 | 0.95 | 0.43 | 0.63 | 0.53 | 0.57 |
| **scanvi_50_lhidden** | 0.62 | 0.36 | 0.59 | 0.89 | 0.15 | 0.70 | 0.95 | 0.20 | 0.58 | 0.52 | 0.55 |
| **scanvi_50_lhidden_esvl** | 0.62 | 0.36 | 0.58 | 0.89 | 0.15 | 0.70 | 0.94 | 0.18 | 0.57 | 0.52 | 0.54 |
| **scvi_50_lhidden_esvl** | 0.62 | 0.40 | 0.52 | 0.93 | 0.14 | 0.73 | 0.87 | 0.00 | 0.54 | 0.51 | 0.52 |
| **scvi_large** | 0.57 | 0.37 | 0.52 | 0.95 | 0.15 | 0.81 | 0.84 | 0.00 | 0.55 | 0.49 | 0.51 |
| **scvi_large_es_lb** | 0.59 | 0.31 | 0.51 | 0.94 | 0.14 | 0.72 | 0.89 | 0.00 | 0.54 | 0.47 | 0.50 |
| **scvi** | 0.58 | 0.33 | 0.53 | 0.93 | 0.14 | 0.64 | 0.90 | 0.00 | 0.52 | 0.48 | 0.50 |
| **scvi_50_lhidden** | 0.58 | 0.30 | 0.52 | 0.94 | 0.15 | 0.75 | 0.85 | 0.00 | 0.54 | 0.47 | 0.50 |
| **scvi_50_esvl** | 0.57 | 0.29 | 0.53 | 0.93 | 0.14 | 0.73 | 0.91 | 0.00 | 0.54 | 0.46 | 0.49 |
| **pca** | 0.60 | 0.36 | 0.56 | 0.91 | 0.13 | 0.56 | 0.79 | 0.00 | 0.48 | 0.50 | 0.49 |
| **scvi_50** | 0.57 | 0.29 | 0.52 | 0.93 | 0.14 | 0.74 | 0.90 | 0.00 | 0.54 | 0.46 | 0.49 |
| **scvi_es_lb** | 0.55 | 0.31 | 0.51 | 0.93 | 0.14 | 0.64 | 0.90 | 0.00 | 0.52 | 0.46 | 0.48 |
| **scvi_es** | 0.50 | 0.27 | 0.51 | 0.93 | 0.14 | 0.67 | 0.90 | 0.00 | 0.53 | 0.43 | 0.47 |
| **scvi_large_es** | 0.45 | 0.23 | 0.52 | 0.92 | 0.14 | 0.66 | 0.92 | 0.00 | 0.53 | 0.40 | 0.45 |

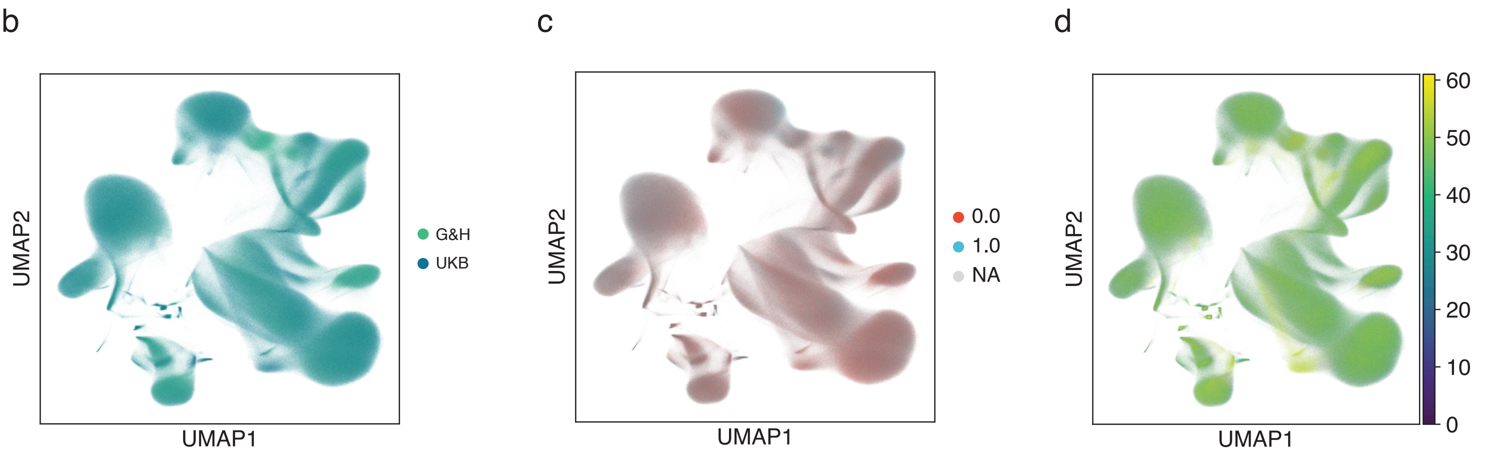


**Supplementary Fig. 3: Integration metrics**
**a**, Results of benchmarking of single-cell data integration methods considered in the study. **b-d**, After batch correction using scANVI, we verified the distribution of the main batch variable in the batch-corrected data, considering cohort **(b)**, sex **(c)**, and sequencing pool **(d)**.

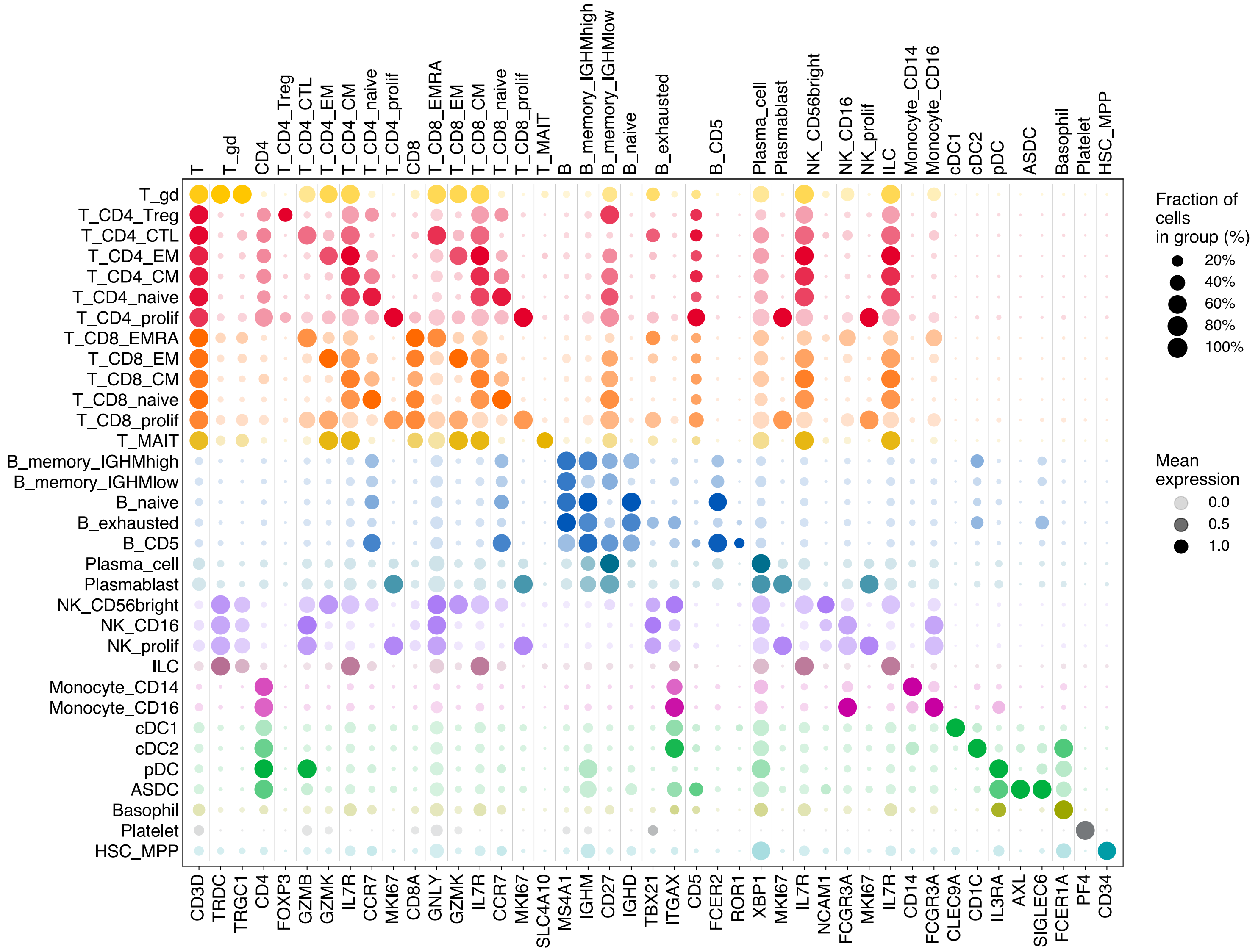


**Supplementary Fig. 4: Cell type 2 marker genes**

Cell type-specific expression of canonical marker genes for celltype 2 (CT2) cell types.

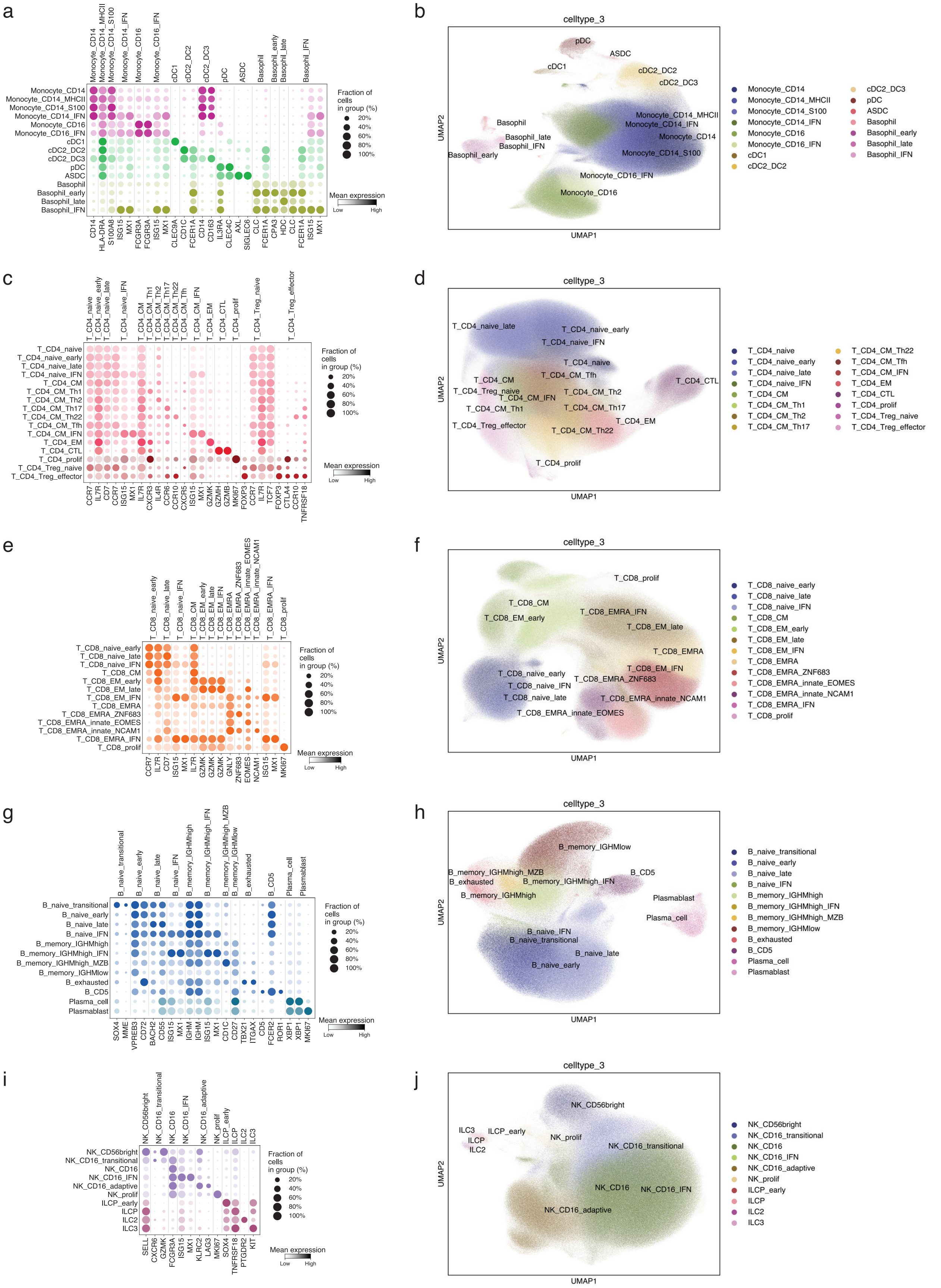

**Supplementary Fig. 5: Marker genes and UMAPs for CT3 cell states**

Cell type-specific expression of canonical marker genes and final UMAPs representing CT3 cell states across main cell lineages: **(a-b)** Monocytes and dendritic cells; **(c-d)** CD4$^{+}$ T cells; **(e-f)** CD8$^{+}$ T cells; **(g-h)** B and plasma cells; **(i-j)** Natural Killer (NK) and Innate Lymphoid Cells (ILCs).

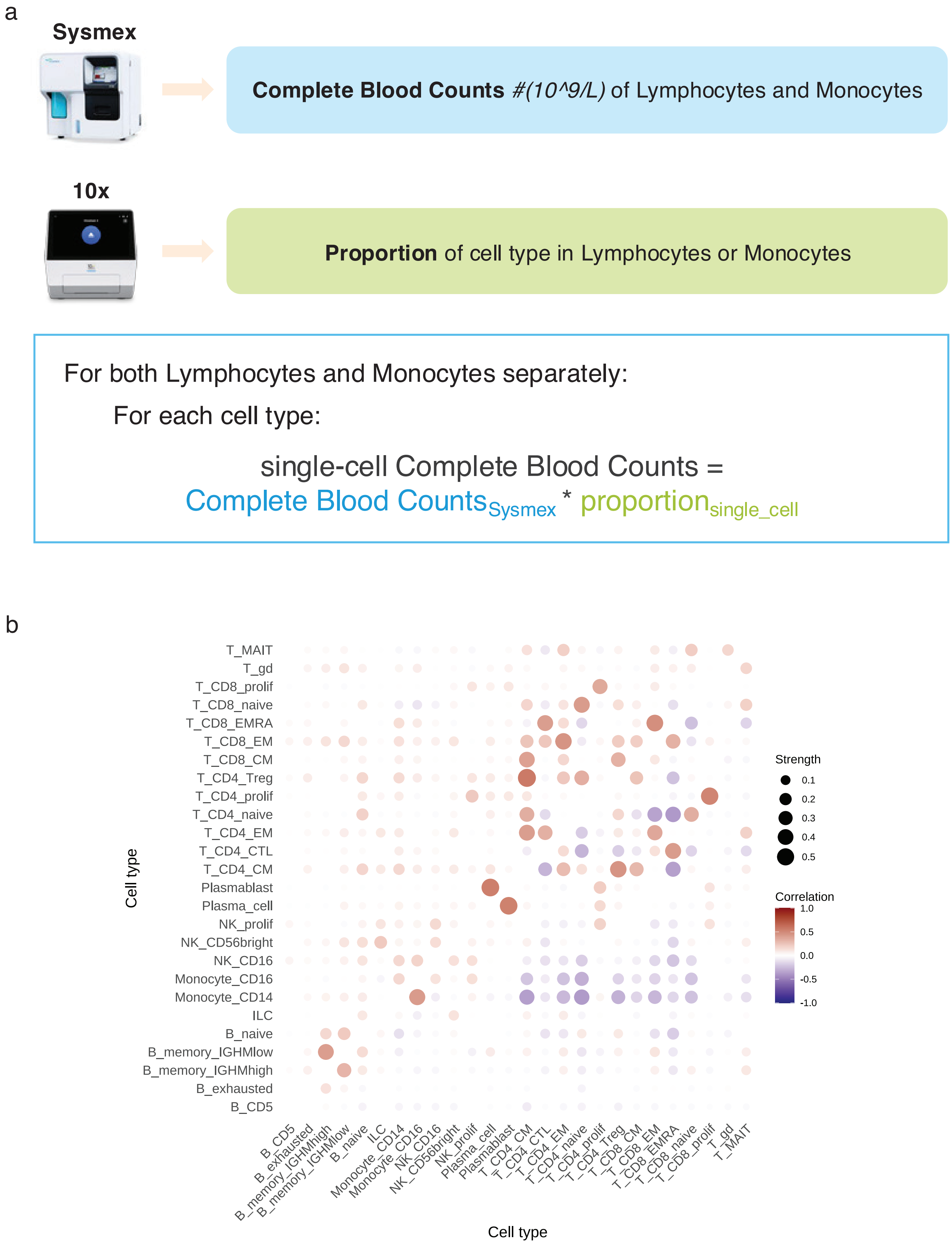


**Supplementary Fig. 6: Single-cell Complete Blood Counts**
**a**, Schematic of computation of single-cell Complete Blood Counts. **b**, Correlations between cell types computed using scCBCs (upper triangle) and proportions (lower triangle) for UKB cohort at CT2 level.

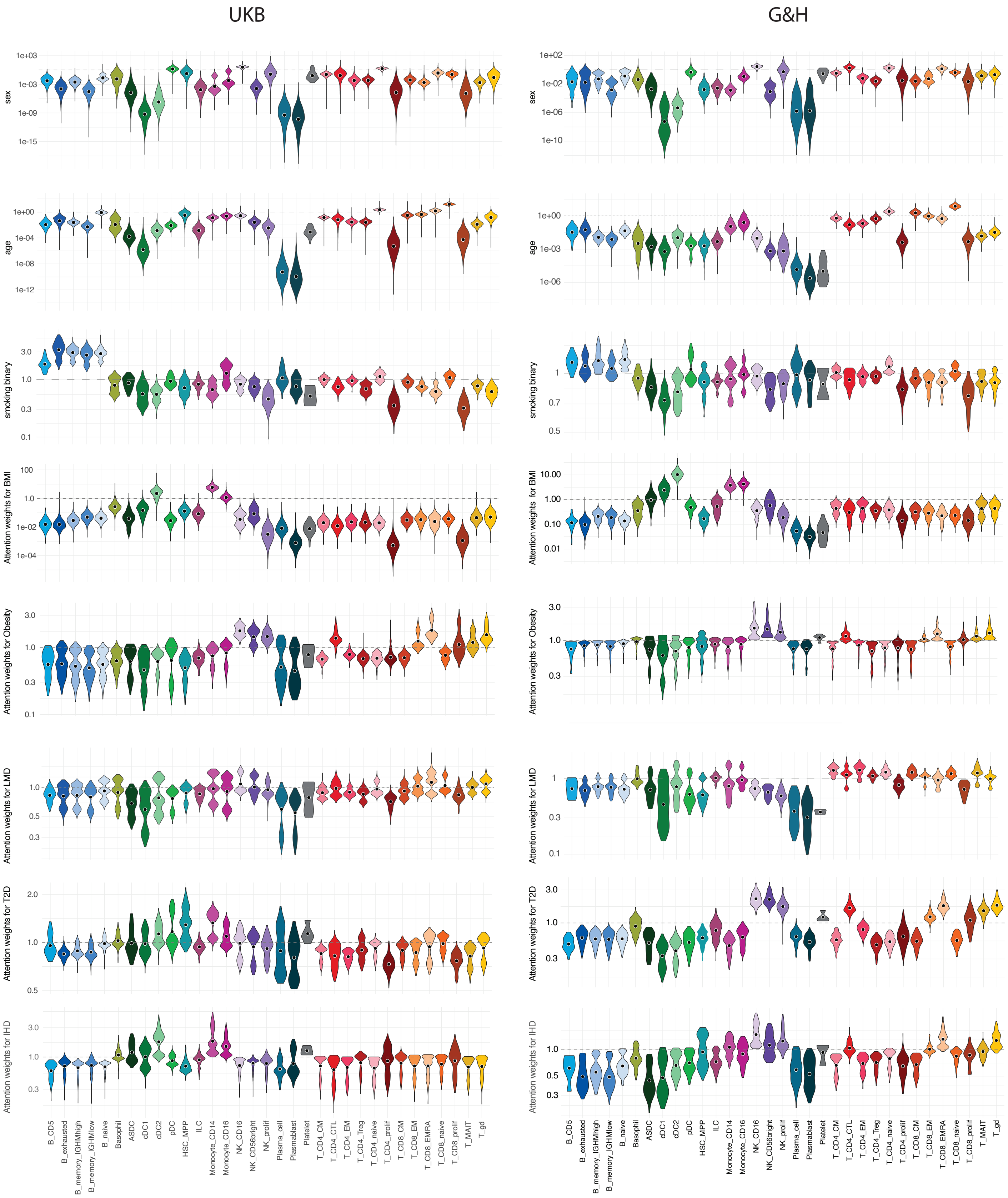


**Supplementary Fig. 7: MixMIL attention weights for selected programmes**
Violin plots illustrating the distribution of donor-specific attention scores across celltype 2 annotations for the UK Biobank (left) and Genes & Health (right) cohorts. The y-axis is shown on a $\log_{10}$ scale. Horizontal dashed grey lines denote the significance threshold (y = 1), above which cell types are considered informative for trait prediction. Black points represent the mean attention score for each cell type. The data relate to the predictive modeling described in **Figure 3**.

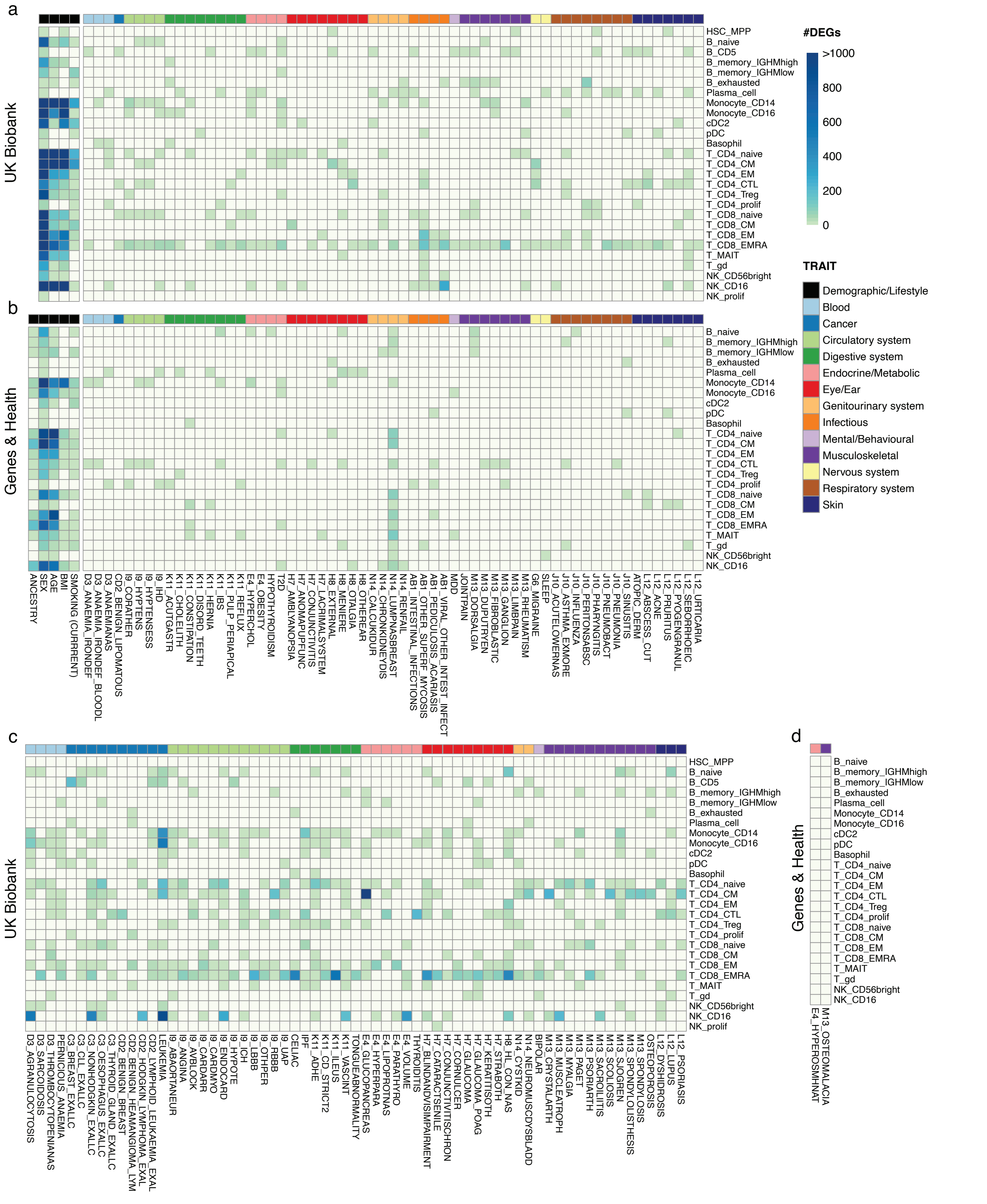

**Supplementary Fig. 8: Cross-cohort heatmap of cell type–specific disease-associated differential expression signatures**
Heatmaps showing the number of differential genes at FDR $< 0.05$ in each cell type as a function of demographic and lifestyle attributes (used as covariates) and contrasting cases and controls for available EHR. **a, b**, Results within the UK Biobank and Genes & Health cohorts, respectively, for 61 traits testable in both cohorts. **c**, Results for 65 out of 220 traits tested only in the UK Biobank, selected for having the largest number of DEGs. **d**, Results for 2 traits tested only in the Genes & Health cohort.

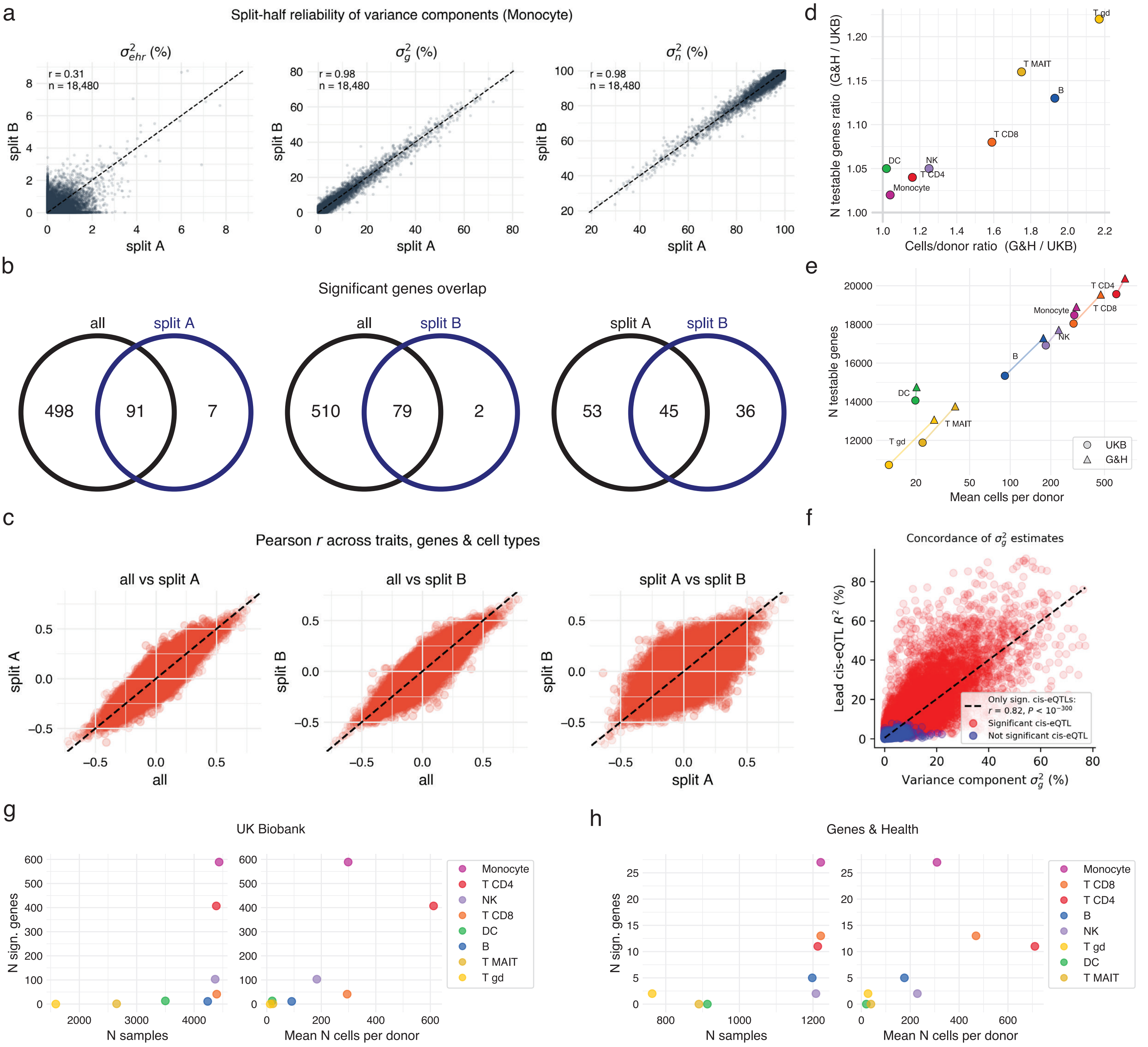


**Supplementary Fig. 9: Benchmarking and robustness assessment of variance decomposition analyses across cohorts and immune cell types**

**a**, Split-half reproducibility of variance component estimates in monocytes from the UK Biobank cohort. Donors were randomly partitioned into two equal subsets (split A and split B), and variance explained by EHR effects ($\sigma^2_{ehr}$), cis-genetic effects ($\sigma^2_g$), and residual variance ($\sigma^2_n$) was independently estimated in each subset. Each point represents one gene; Pearson correlation coefficients are indicated. Genetic and residual variance estimates were highly reproducible across splits, whereas EHR variance components showed lower but detectable concordance. **b**, Overlap of genes with significant EHR variance components (empirical FDR $\leq$ 0.05) comparing the full dataset ("all") and each split-half subset. Venn diagrams indicate the number of shared and unique significant genes. **c**, Concordance of trait-level EHR effects across analyses. Scatterplots show correlations between leave-one-out best linear unbiased predictors (LOO-BLUPs) derived from EHR covariates across all tested trait–gene–cell type combinations, comparing the full dataset with split A, and split B. Each point corresponds to one trait–gene–cell type association. **d**, Relative increase in statistical power in Genes & Health (G&H) compared with UK Biobank (UKB) as a function of cellular sampling depth. The x-axis shows the ratio of mean cells per donor between cohorts (G&H/UKB), and the y-axis shows the corresponding ratio of testable genes. Cell types with greater cellular coverage in GH yielded proportionally more testable genes. **e**, Relationship between mean number of cells per donor and the number of testable genes across immune cell types and cohorts. Circles denote UKB and triangles denote GH. **f**, Lead cis-eQTL $R^2$ versus cis-genetic variance (var_g) for each gene–cell type pair, coloured by whether the gene is a significant cis-eQTL (red) or not (blue). The lead-variant $R^2$ and the kernel-based var_g estimate are positively correlated (Pearson correlation, r = 0.82). **g**, **h**, Influence of cohort size and cellular sampling depth on the number of detected genes with significant EHR variance components in UKB (g) and G&H (h). Detection power was more strongly associated with cellular sampling depth than with donor number alone.

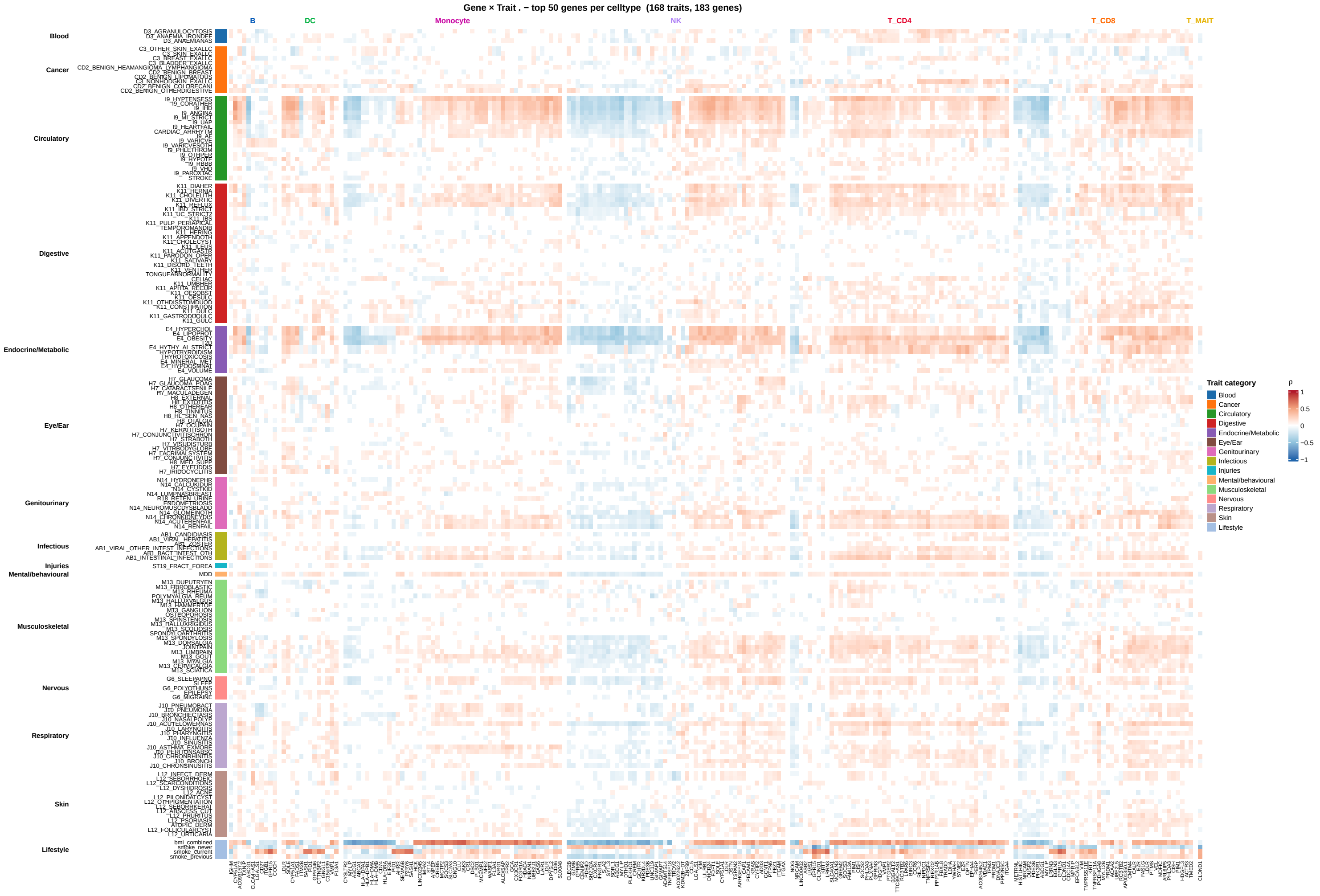


**Supplementary Fig. 10: Variance decomposition effects matrix for UKB Biobank**

The heatmap shows the top 50 genes per cell type, ranked by the variance explained by the EHR component, as computed in the variance decomposition analysis for the UKB cohort. Values represent Pearson's correlation coefficient (R) for each gene's correlation to each trait. Genes are clustered by cell type based on Euclidean distance computed on R values across all traits. Similarly, traits are clustered within each category based on Euclidean distance computed on R values across all genes.

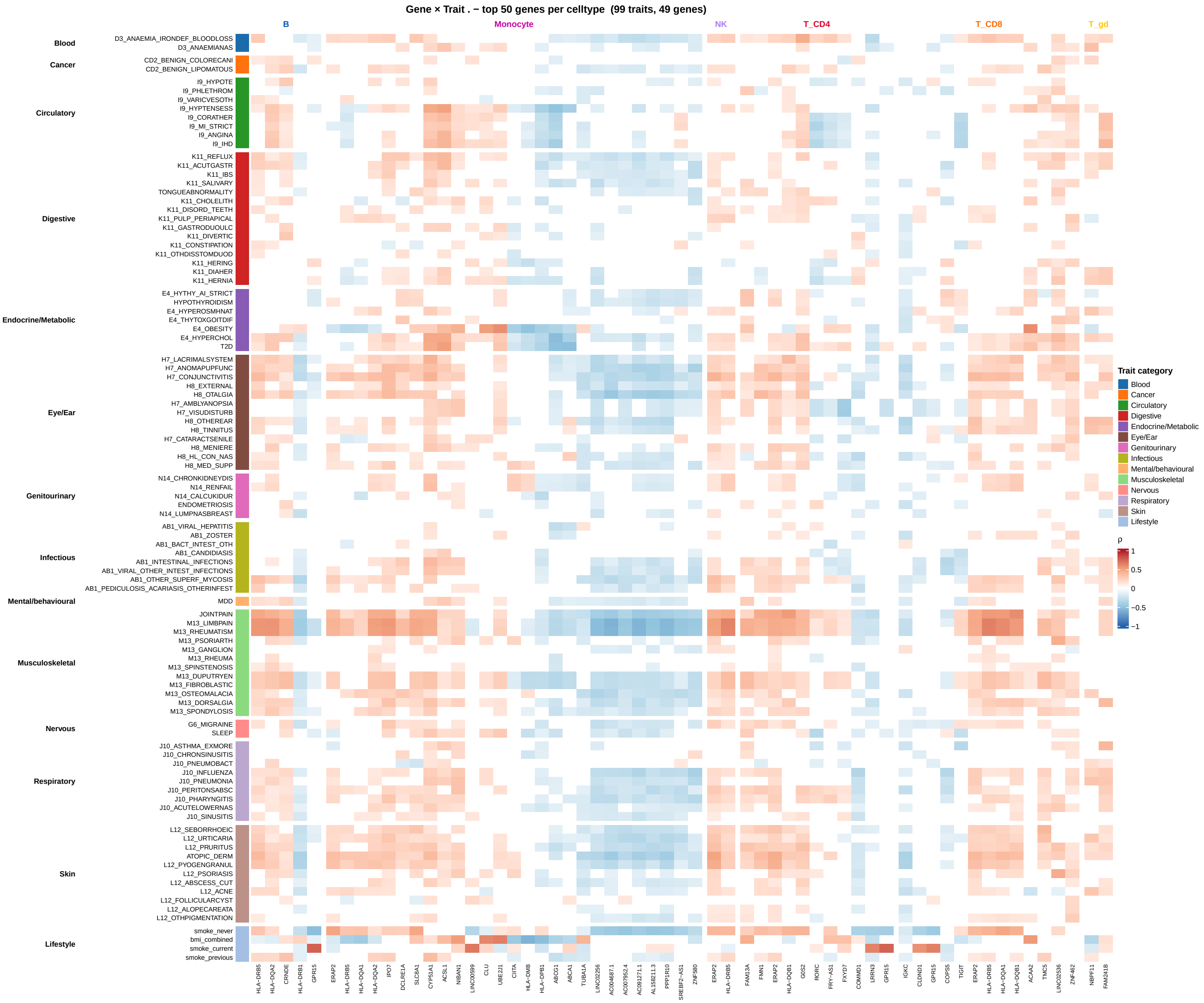


**Supplementary Fig. 11: Variance decomposition effects matrix for Genes&Health**

The heatmap shows the top 50 genes per cell type, ranked by the variance explained by the EHR component, as computed in the variance decomposition analysis for the Genes&Health cohort. Values represent Pearson's correlation coefficient (R) for each gene's correlation to each trait. Genes are clustered by cell type based on Euclidean distance computed on R values across all traits. Similarly, traits are clustered within each category based on Euclidean distance computed on R values across all genes.

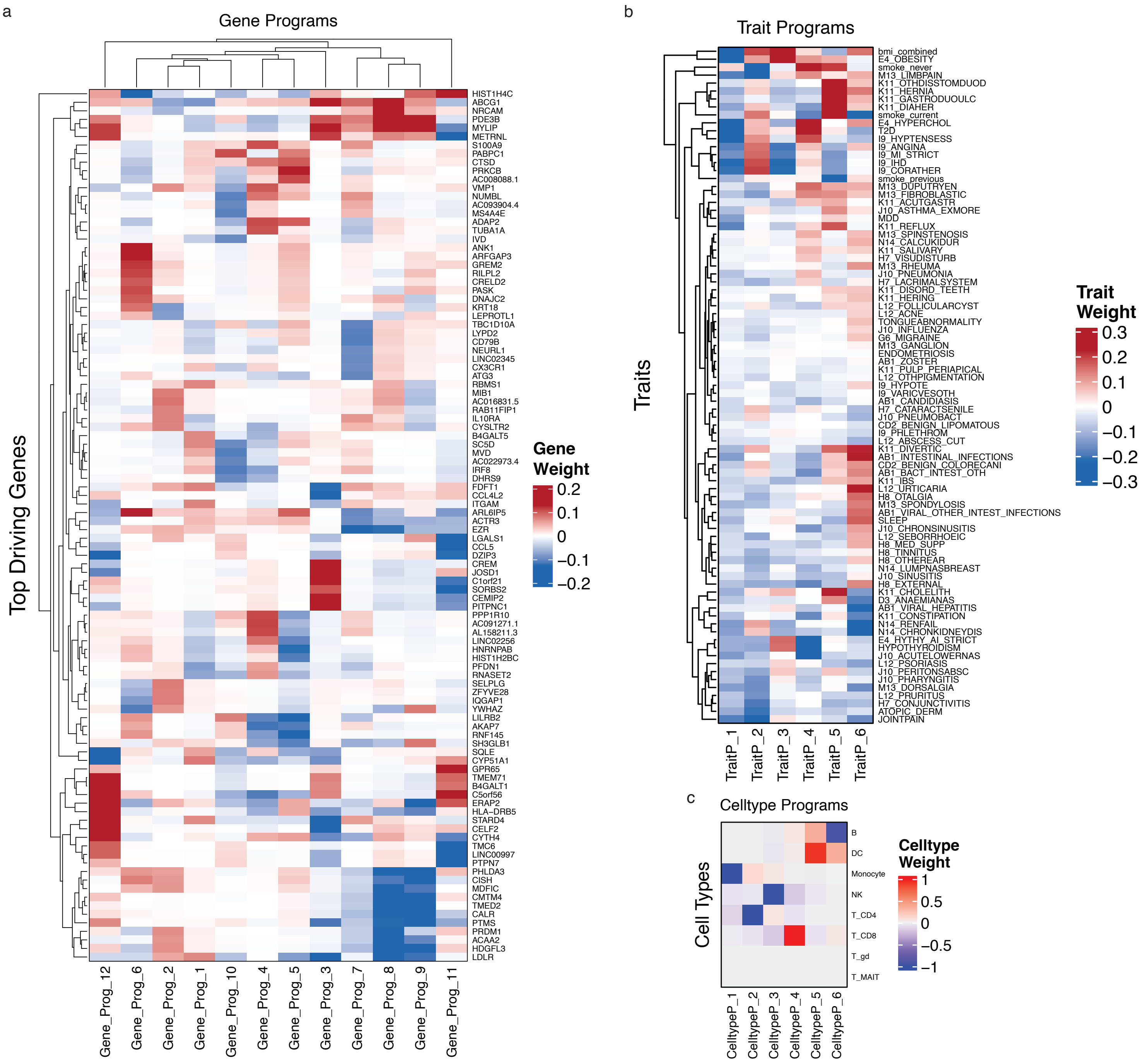


**Supplementary Fig. 12: Tucker decomposition loadings matrix**

Variance decomposition latent programs loadings. **a-c**, Heatmaps representing the loading matrix on the gene **(a)**, trait **(b)**, and celltype **(c)** dimensions. For genes, only the top 10 contributing genes for each gene program are shown.

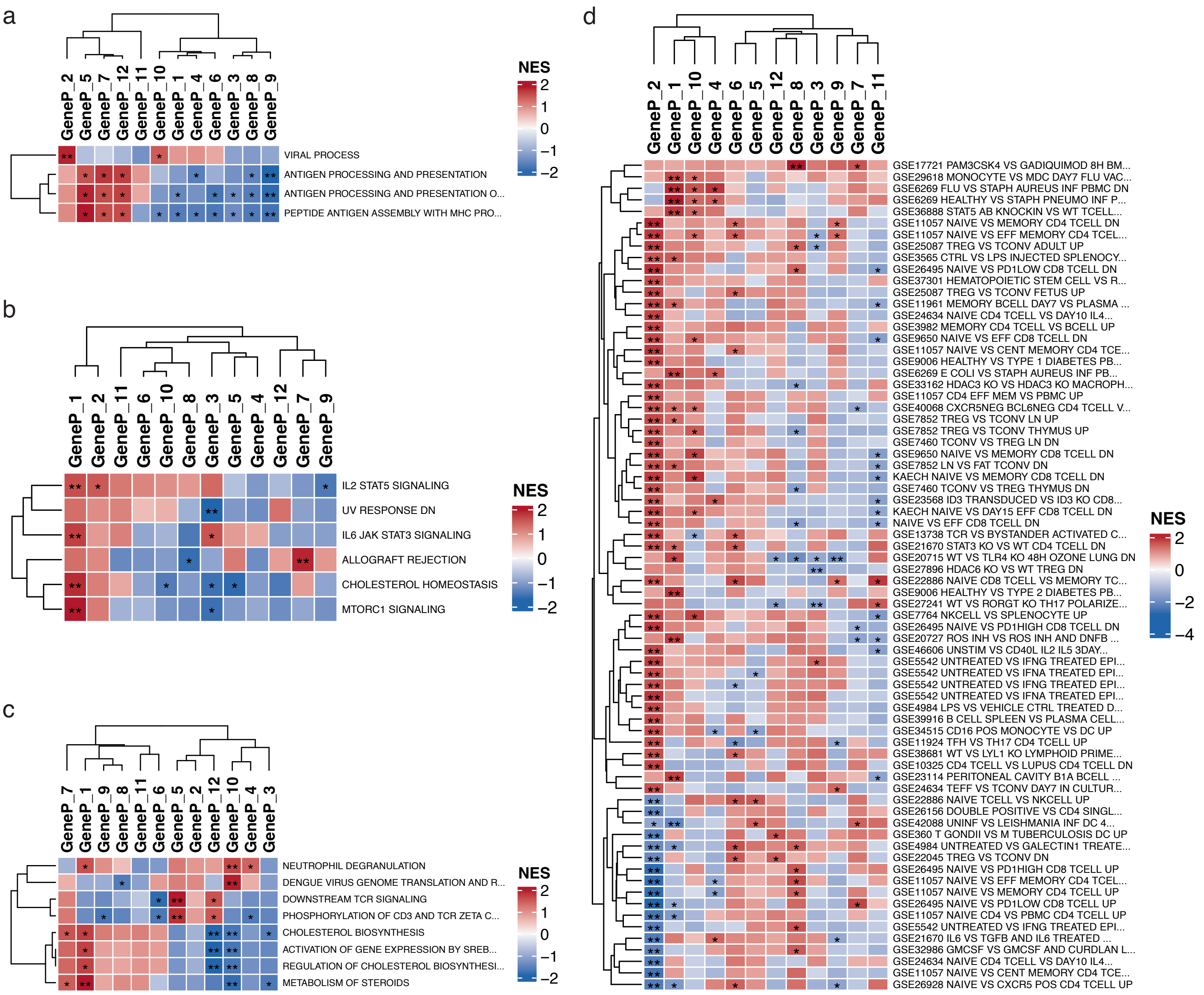


**Supplementary Fig. 13: GSEA analysis results for gene programs inferred from Tucker decomposition**

GSEA results on gene programs. To assist in the interpretation of the gene programs, we performed gene-set enrichment analysis across different gene sets from MSigDB. **a-d**, Heatmaps showing enrichment results for Gene Ontology Biological Processes **(a)**, human hallmark gene sets **(b)**, REACTOME pathways **(c)**, and ImmuneSigDB gene sets **(d)**. Enrichments with adjusted p-value $< 0.05$ or nominal p-value $< 0.05$ are marked by a double and a single star, respectively.

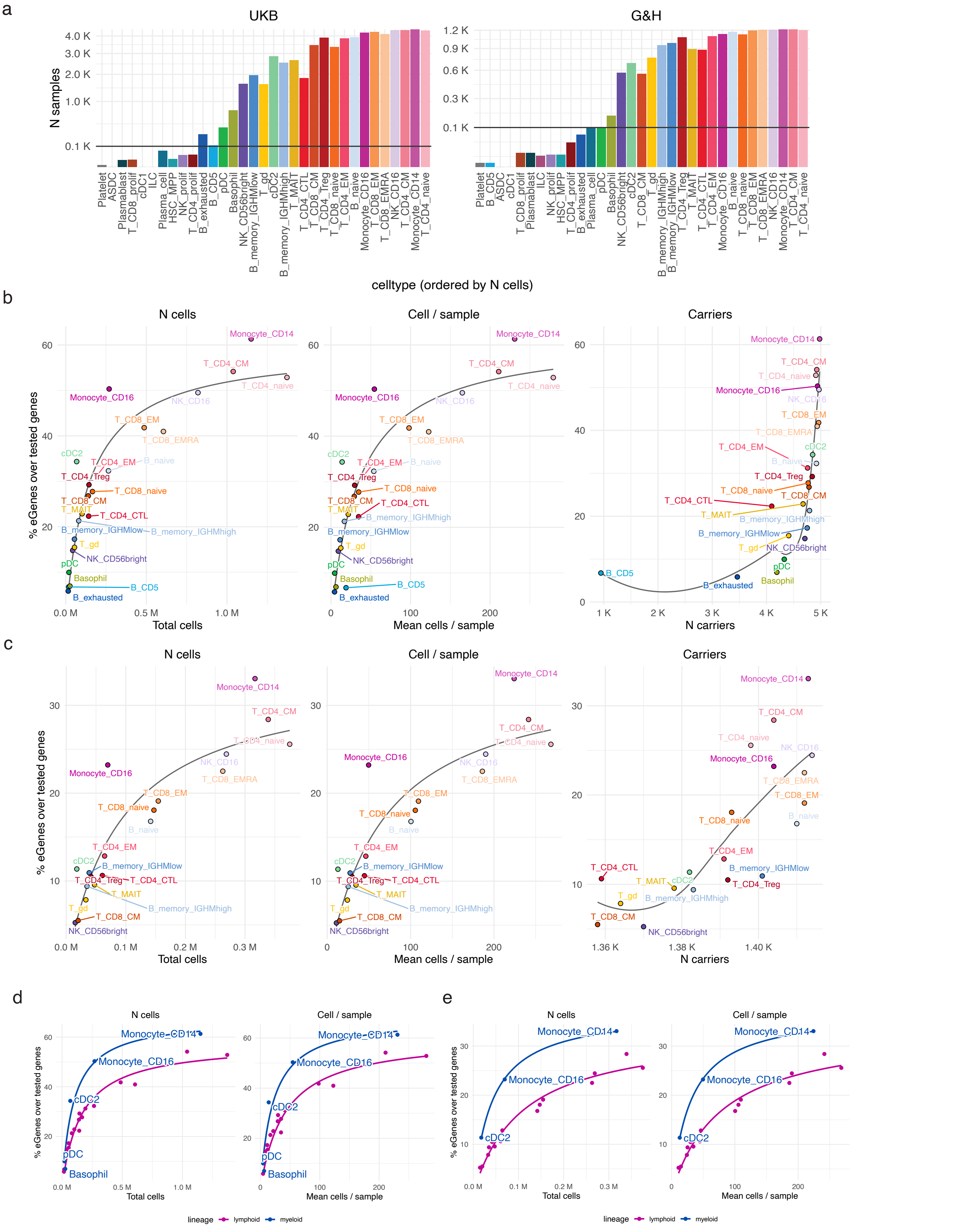


**Supplementary Fig. 14: Main factors influencing eQTL discovery power.**

**a**, Barplot representing the number of samples available for eQTL analysis in each cell type in the UKB and G&H cohort. The solid line represents the minimum sample threshold (100 samples) for a cell type to enter eQTL analysis. **b, c**, effect of total number of cells (N cells), cell per sample (Cell / sample) and number or sample carriers (Carriers) on eGene discovery, reported as the percentage of eGenes detected over total number of genes tested. The same plots are represented for UKB cohort (b) and G&H cohort (c). **d, e**, effect of total number of cells (N cells) and cell per sample (Cell / sample) on eGene discovery, stratified by myeloid or lymphoid cell types. This clearly shows distinct trends for myeloid and lymphoid cells, with myeloid cells yielding more eGenes at a comparable number of cells.

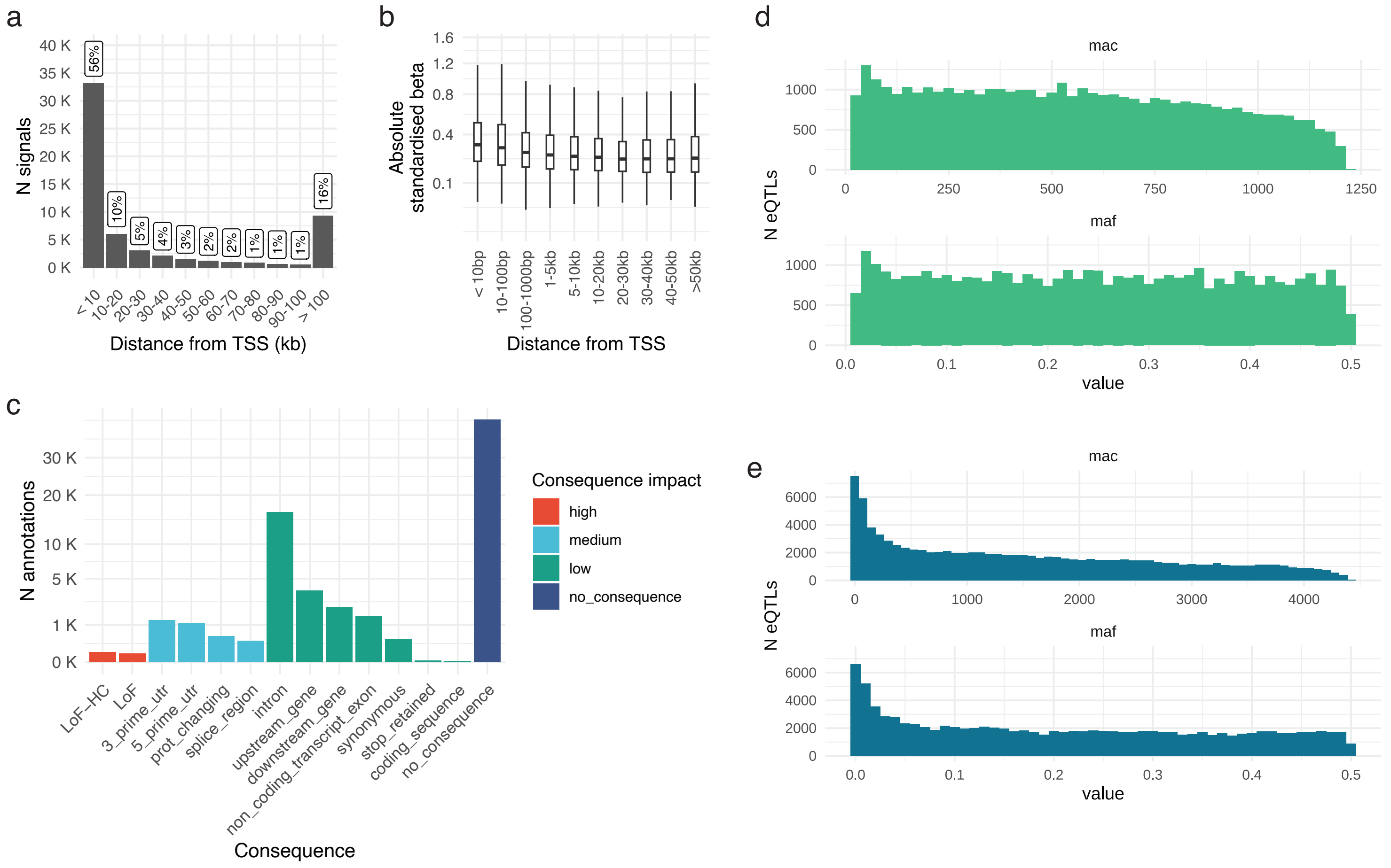


**Supplementary Fig. 15: Characterization of eSNPs.**
**a**, Distribution of eSNPs by distance from the eGene TSS. The percentages represent the fraction of total signals in a given bin. **b**, Distribution of eSNPs effect size across the TSS distance bins. Only signals with PIP $\geq$ 0.7 were considered in a and b. **c**, barplot representing the distribution of gene consequences for the identified eSNPs. Consequences are determined based on the associated eGene and grouped based on their putative disruptive impact into high, medium, low or no consequence. The no consequence group represents variants without any direct impact on the corresponding eGene. **d, e**, Distribution of eSNPs across MAC and MAF values in the G&H (d) and UKB (e) cohort.

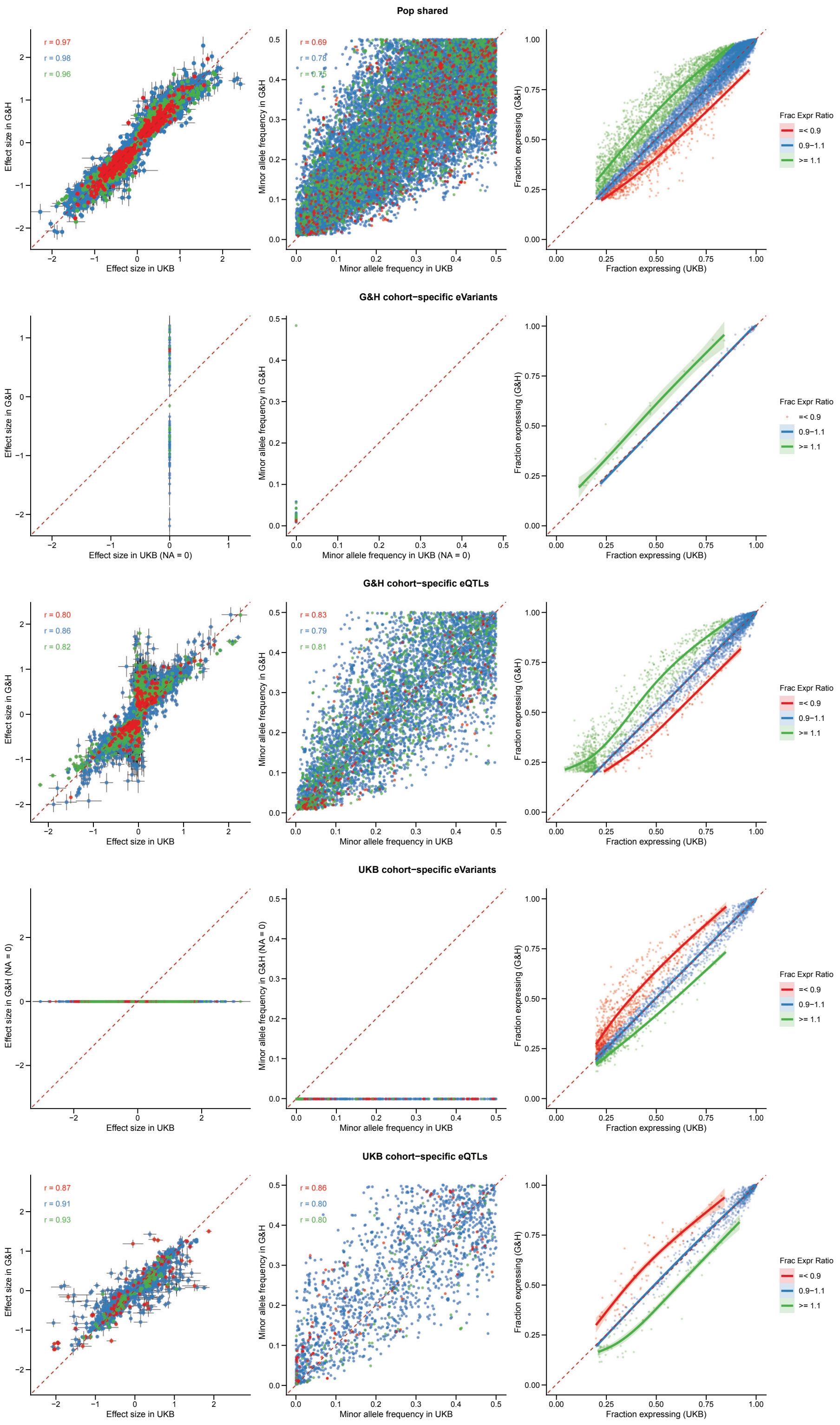


**Supplementary Fig. 16: Cross-population comparison of eQTL effect sizes, allele frequencies, and gene expression for all the subsets of variants highlighted in Figure 5.**

In all panels, the dashed red line indicates the identity line (y = x); Pearson correlation coefficients are shown in the top-left corner. Effect sizes were estimated using TensorQTL on pseudobulk expression within each cell type. The fraction of expression in each cohort was calculated as the number of donors with expression $> 0$ for a given cell type–gene pseudobulk pair, divided by cohort sample size. For G&H, this fraction was calculated across pooled Bangladeshi and Pakistani individuals (combining both expressing-individual counts and total sample sizes across the two groups). The fraction-of-expression ratio was then computed reciprocally between cohorts: G&H/UKB and UKB/G&H.

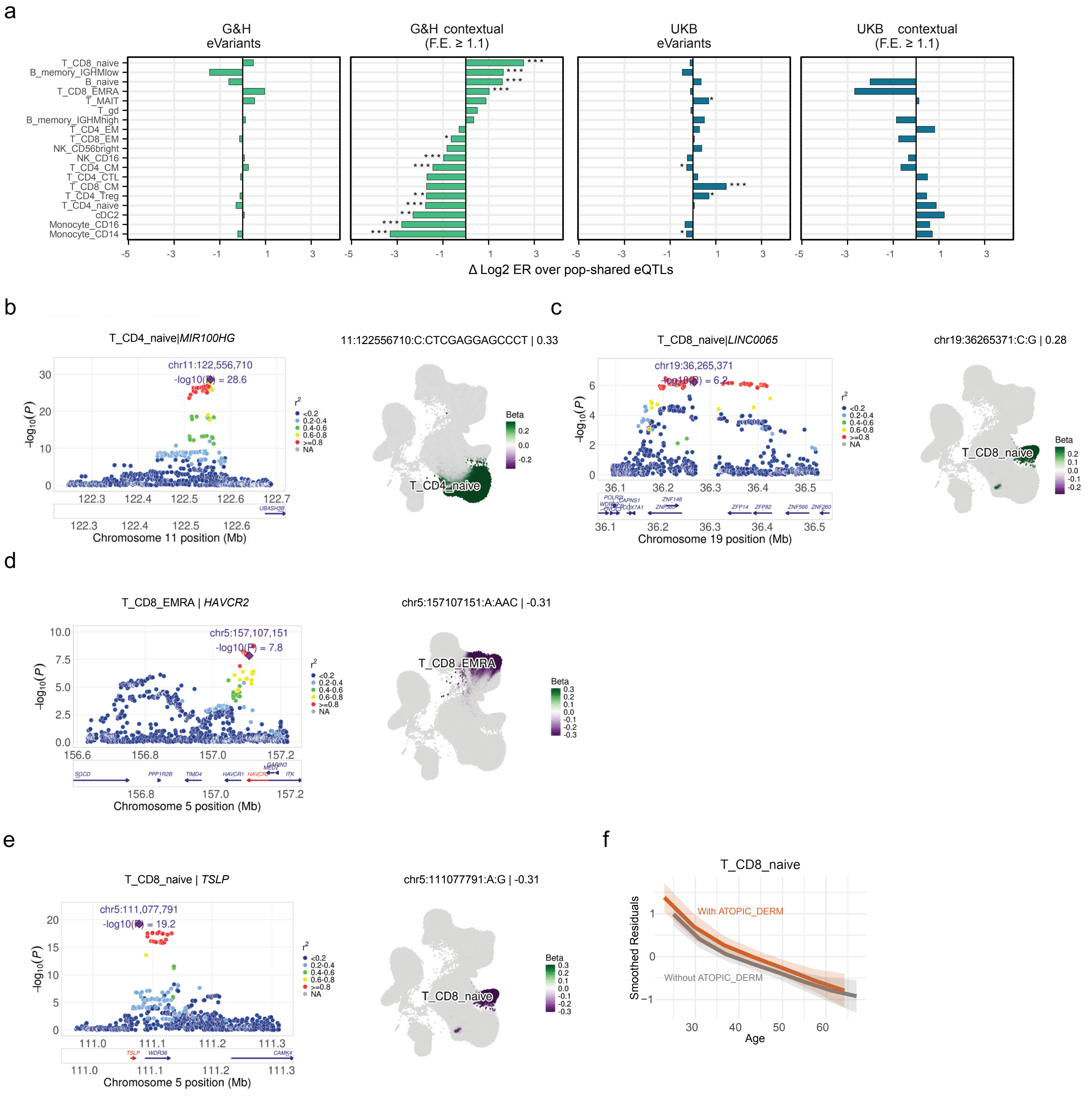


**Supplementary Fig. 17: Characteristics and examples of contextual eQTLs F.E. ≥ 1.1**

**a**, Difference in $\log_2$ enrichment between G&H eVariants, G&H contextual eQTLs F.E. ≥ 1.1, UKB eVariants, and UKB contextual eQTLs F.E. ≥ 1.1 relative to population-shared eQTLs. Significant enrichment and depletion patterns observed for G&H contextual eQTLs F.E. ≥ 1.1 were not recapitulated by G&H eVariants, which displayed distributions similar to those of population-shared eQTLs. Statistical significance was assessed using Fisher's exact test (* p value $<$ 0.05,** p value $<$ 0.01 and *** p value $<$ 0.001). **b**, LocusZoom plot (left) and UMAP projection (right) illustrating the eQTL 11:122556710:C:CTCGAGGAGCCCT (rs377086917) associated with *MIR100HG* expression in naive $CD4^+$ T cells. **c**, LocusZoom plot (left) and UMAP projection (right) illustrating the eQTL 19:36261765:C:T (rs73040148) associated with *LINC00665* expression in naive $CD8^+$ T cells. **d**, LocusZoom plot (left) and UMAP projection (right) illustrating the eQTL 5:157117907:C:T (rs10057431) associated with *HAVCR2* expression in EMRA $CD8^+$ T cells. **e**, LocusZoom plot of the eQTL 5:111077791:A:G (rs10073816) associated with *TSLP* expression in naive $CD8^+$ T cells. **f**, Association between naive $CD8^+$ T cell abundance and atopic dermatitis status in the G&H cohort.

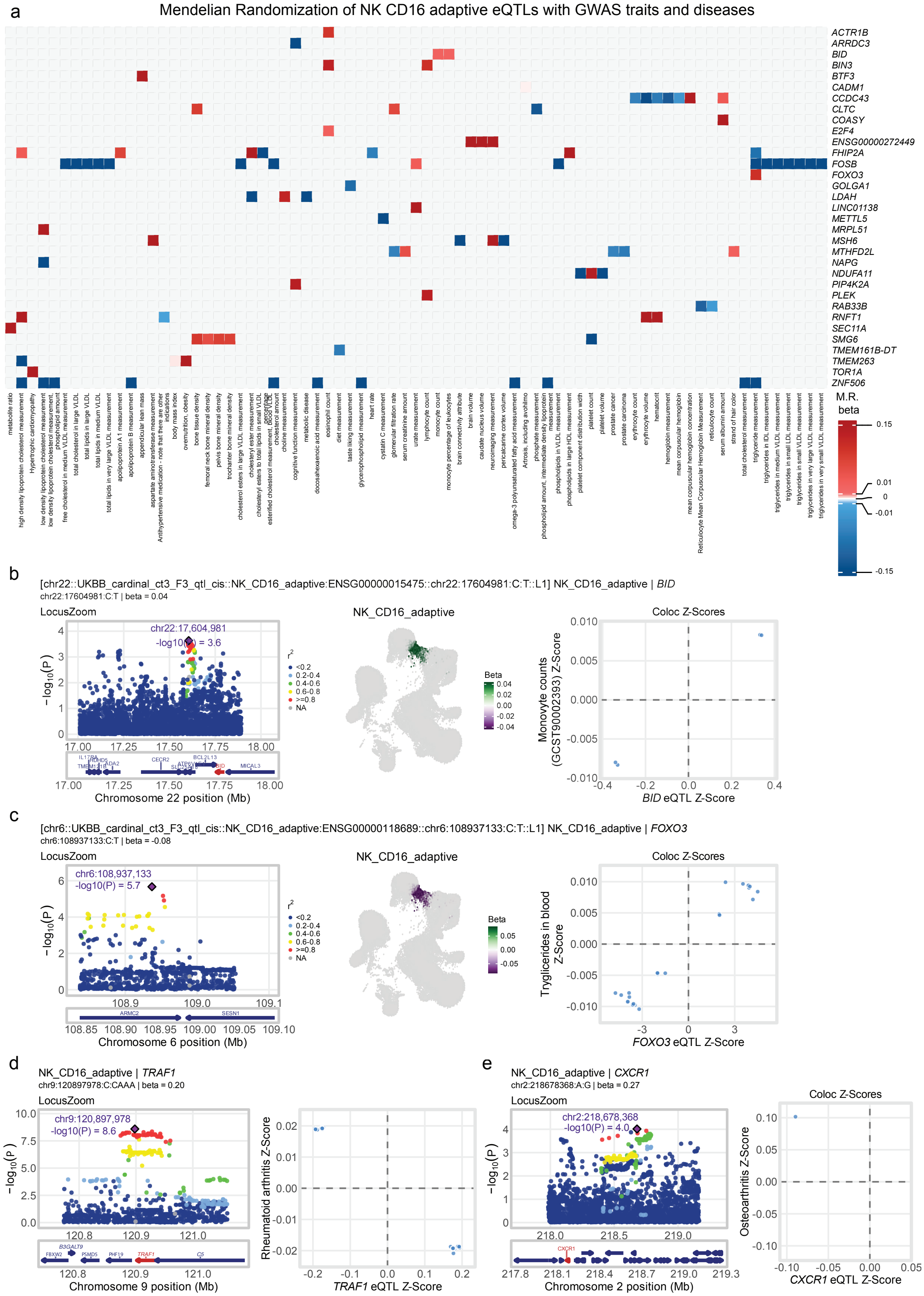


**Supplementary Fig. 18: Mendelian randomisation in NK_CD16_adaptive.**

**a**, Heatmap of Mendelian randomisation associations between NK_CD16_adaptive eQTLs and GWAS traits. Red denotes positive effects of gene expression on trait values, whereas blue denotes negative effects. **b**, NK cell–specific eQTLs with evidence of causal effects included *BID*, associated with increased monocyte counts (GCST90002393). **c**, NK cell–specific eQTLs with evidence of causal effects *FOXO3*, associated with elevated circulating triglyceride levels. **d**, eQTLs also detected in GTEx tissues included *TRAF1*, associated with rheumatoid arthritis (GCST002318). **e**, eQTLs also detected in GTEx tissues included *CXCR1* associated with knee osteoarthritis (GCST90566800).

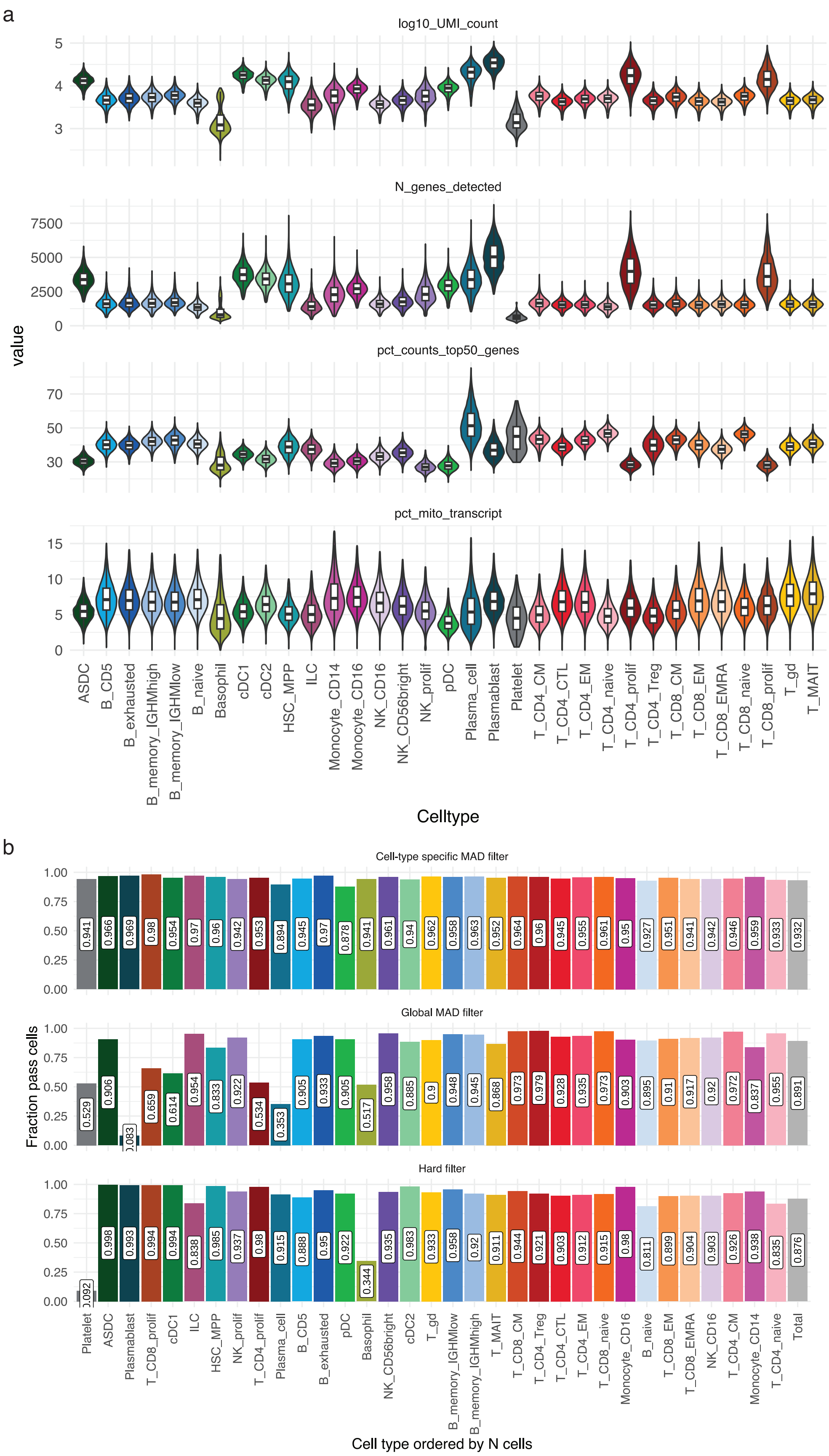


**Supplementary Fig. 19: Cell-level quality control**

**a**, Quality control metric distributions across cell types. **b**, Comparison of cell loss from adaptive versus hard QC filters across cell types. Using a cell-type specific MAD filter results in more uniform filtering, especially for rare cell types.

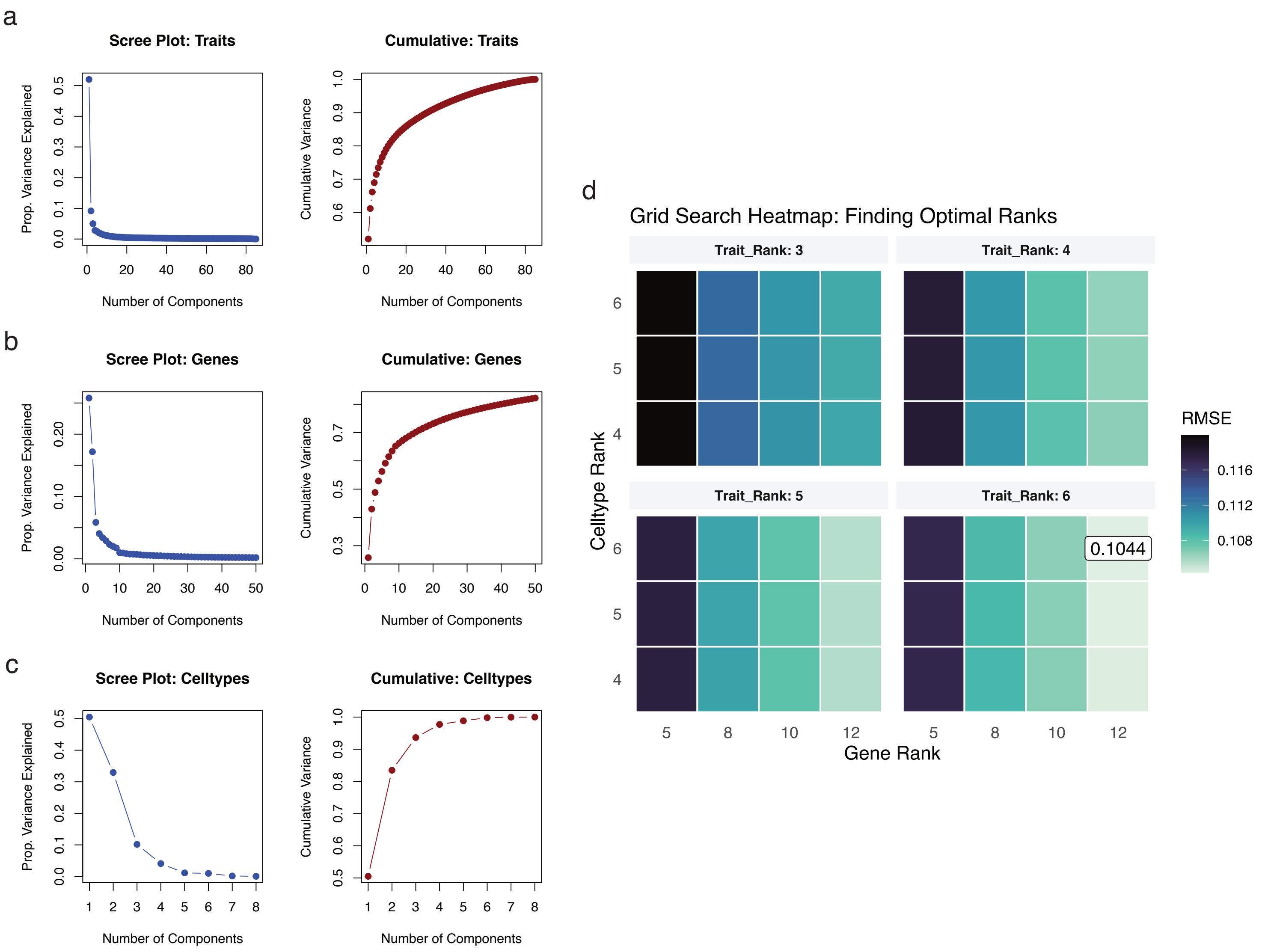


**Supplementary Fig. 20: Optimal ranks for Tucker decomposition**
Latent programs extraction. **a-c**, Scree plot and cumulative variance based on SVD of the trait (a), gene (b), and cell type (c) dimensions in the Tucker decomposition tensor. **d**, Rank boundaries were estimated using the elbow method and refined via parallelised grid search cross-validation. The optimal out-of-sample root mean square error (RMSE) is reported.

### Supplementary Table Legends

**Supplementary Table 1: Sample demographics summary on the QCed dataset**

Sample size, distribution of the main demographic variables (age, sex, ancestry) and disease prevalence for the main diseases in the UK Biobank and Genes & Health cohorts. We report values computed on the whole cohort and on the subset of individuals included in the CARDINAL dataset, after QC. Prevalence values reported here refer to the full QCed CARDINAL dataset, while numbers in Supplementary Table 5 refer to the restricted dataset. For numerical variables, the table reports the mean value (with min-max in brackets); for categorical variables, the table reports N samples in the group (with percentage over individuals with a valid measurement in brackets).

**Supplementary Table 2: Medical record prevalence on the restricted dataset**

Number of cases and prevalence for the 349 disease phenotypes considered in the study. Disease definitions (PHECODE, Phenotype label and Name pheno) were derived from the MVP–FinnGen–UK Biobank meta-analysis as described in methods. Counts below five are suppressed and shown as “¡5”.

**Supplementary Table 3: Pool and donor information for RNA sequencing samples**

Table describing donor information, laboratory metadata, pool level meta data, single cell and pool level quality control metrics. The table contains the following information: Pool ID, Identifier for a pool of donors; Pool_ID.Donor_Id, identifier for a pool of donors and Vireo donor ID; Experiment ID, Illumina NovaSeq run ID and run date; Sample ID, blood sample ID; Chromium channel ID, 10x RunID and 10x Chromium channel position; Viability, viability of PBMC cells prior to pooling; Time from venepuncture, time between venepuncture and PBMC extraction; lab_live_cell_count, concentration of PBMC prior to pooling; GEM Batch, 10x Chromium GEM batch identifier; Total Nr cells for donor, number of cells assigned to the sample by Vireo before QC; Genes detected with counts $> 0$, number of genes detected; Genes with UMI count $\geq 3$, number of genes with a minimal UMI count; Median UMIs per cell, Median UMI per cell; Collection Site, Donor collection site; Sample Draw Date, Date of Venepuncture; Donor with less than 100 cells in Total, Number of samples with less than 100 cells including any repeats; Donors in pool, Number of donors sequenced together in a 10x Chromium pool; Spikeins expected in the pool, Inlcusion of U937 and/or THP-1 cell-line spike-in samples in the pool; Number of Reads, Number of sequencing reads mapped for the 10x Chromium reaction; Fraction Reads in Cells, Fraction of Reads assigned to a valid cell barcode by cellranger; Mean Reads per Cell, Mean reads per cell; Droplets Containing a Single Cell, Number of droplets capturing a single cell for the 10x Chromium reaction; Sysmex Haematology data, Donor with Sysmex CBC Haematology data (24h); included_in_QCed_dataset: indicates if the sample was included in the dataset after QC.

**Supplementary Table 4: Cell annotation gene markers**

Manually curated cell type labels across 5 granularities (from broad to more granular): lineage_1, lineage_2, celltype_1 (CT1), celltype_2 (CT2), and celltype_3 (CT3). We also report the marker genes used to define the CT3 cell states (marker_genes) and the CT2 cell types (marker_genes_celltype_2). For the CT3 marker genes, we also compiled a minimal list distinguishing the CT3 states from each other (marker_genes_short), including a version excluding the lineage markers (marker_genes_short_-nolineage).

**Supplementary Table 5: Number of cells and number of samples per cell type**

For each cell type at CT1, CT2, and CT3 cell type annotation level, we report the number of cells, the number of carriers, and the mean number of cells per sample after QC in the UKB and G&H cohorts.

**Supplementary Table 6: Summary of donors and cells used in analyses**

Summary of the sample size (cells and individuals) for the datasets used in different analyses in the study.

**Supplementary Table 7: Compositional signatures for UK Biobank and Genes & Health**

For the main demographic variables, risk exposures and disease, the table reports the celltype signatures identified for CT2 and CT3 celltypes in the two cohorts, based on the MixMIL analysis described in methods. Numbers in bracket represent the estimated beta.

**Supplementary Table 8: MixMil signatures for UK Biobank and Genes & Health**

For the main demographic variables, risk exposures and disease, the table reports the celltype composition signatures identified for CT2 and CT3 celltypes in the two cohorts, based on the analysis of scCBC and the regression model described in methods. Numbers in brackets represent the attention enrichment averaged across donors for each cell type.

**Supplementary Table 9: Variance decomposition – genes with significant EHR component (UK Biobank)**

Detailed results of the gene expression variance decomposition analysis on UKB cohort, estimating for each gene celltype the proportion of gene expression variance explained by EHR component (var_ehr) or cis-genetics component (var_g). Only gene-celltype combinations where the EHR component has a significant contribution are reported. See methods.

**Supplementary Table 10: Variance decomposition – genes with significant EHR component (Genes & Health)**

Detailed results of the gene expression variance decomposition analysis on GH cohort, estimating for each gene celltype the proportion of gene expression variance explained by EHR component (var_ehr) or cis-genetics component (var_g). Only gene-celltype combinations where the EHR component has a significant contribution are reported. See methods.

**Supplementary Table 11: Variance decomposition – gene to trait significant correlations for EHR terms (UK Biobank)**

For gene-celltype combinations with a significant effect of the EHR component in the UKB cohort, the table reports the correlation estimates (pval and Pearson's r) between the EHR component BLUP and the single EHR traits (see methods). For each gene-celltype-trait combination, we also report whether the gene-celltype pair was tested in the eQTL analysis (tested_in_-eQTL) and whether it yielded a significant eQTL association in the relevant celltype (is_eQTL). Finally, we reported whether the same trait also resulted in a significant colocalisation between publicly available GWAS data and single-cell eQTL generated in this study (coloc_same_disease).

**Supplementary Table 12: Variance decomposition – gene to trait significant correlations for EHR terms (Genes & Health)**

For gene-celltype combinations with a significant effect of the EHR component in the GH cohort, the table reports the correlation estimates (pval and Pearson's r) between the EHR component BLUP and the single EHR traits (see methods). For each gene-celltype-trait combination, we also report if the gene-celltype pair was tested in the eQTL analysis (tested_in_eQTL) and if it yielded any significant eQTL association in the relevant celltype(is_eQTL). Finally, we reported whether the same trait also resulted in a significant colocalisation between publicly available GWAS data and single-cell eQTL generated in this study (coloc_same_disease).

**Supplementary Table 13: MR results for BMI**

Complete results for the Mendelian Randomisation analysis on BMI.

**Supplementary Table 14: Gene loadings from the VD latent factors**

Gene loadings for the 12 gene programs resulting from Tucker decomposition applied to variance decomposition results, estimating the correlation between EHR traits and gene expression in single-cell at CT1 cell type level (see Methods).

**Supplementary Table 15: Trait loadings from the VD latent factors**

Gene loadings for the 6 trait programs resulting from Tucker decomposition applied to variance decomposition results, estimating the correlation between EHR traits and gene expression in single-cell at CT1 cell type level (see Methods).

**Supplementary Table 16: Celltype loadings from the VD latent factors**

Gene loadings for the 6 cell type programs resulting from Tucker decomposition applied to variance decomposition results, estimating the correlation between EHR traits and gene expression in single-cell at CT1 cell type level (see Methods).

**Supplementary Table 17: GSEA results on gene loadings – GOBP**

Complete results of the gene set enrichment analysis performed using fgsea on the gene loadings for the 12 gene programs and the Gene Ontology Biological Processes gene sets (as obtained from MSigDB).

**Supplementary Table 18: GSEA results on gene loadings – REACTOME**

Complete results of the gene set enrichment analysis performed using fgsea on the gene loadings for the 12 gene programs and the REACTOME pathways gene sets (as obtained from MSigDB).

**Supplementary Table 19: GSEA results on gene loadings – Human Hallmark**

Complete results of the gene set enrichment analysis performed using fgsea on the gene loadings for the 12 gene programs and the Human Hallmark pathways gene sets (as obtained from MSigDB).

**Supplementary Table 20: GSEA results on gene loadings – ImmuneSigDB**

Complete results of the gene set enrichment analysis performed using fgsea on the gene loadings for the 12 gene programs and the ImmuneSigDB gene sets (as obtained from MSigDB).

**Supplementary Table 21: eGene discovery across tested cell types**

For each cell type tested in the UKB and G&H cohort, the table reports the number of sample, the mean number of cells / sample, the number of tested genes and the number of significant eGenes detected before and after fine-mapping.

**Supplementary Table 22: Pleiotropy classification of independent causal genetic variants (eSNPs) in UKB and G&H**

The pleiotropy category summarises the action of the eSNPs across cell types and genes into 4 categories, namely highly pleiotropic (multiple genes in multiple cell types), multi-context (single gene in multiple cell types), multi-gene (multiple genes in a single cell type), and specific (single gene in a single cell type). For each category we report the number of eSNPs and the percentage relative to the total number of eSNPs in the UKB and G&H cohort.

**Supplementary Table 23: Summary of population-shared, cohort-specific, and contextual eQTLs for the UK Biobank cohort**

**Supplementary Table 24: Summary of population-shared, cohort-specific, and contextual eQTLs for the G&H cohort**

**Supplementary Table 25: Credible sets for eQTL identified in CD16+ adaptive NK, cell-type specificity and colocalisation results**

For each credible sets identified for cis-eQTL signals in adaptive CD16+ NK cells (CT3 level), the table reports gene Ensembl ID, genomic region of association (GRCh38), credible-set identifier, most likely causal variant, its effect and other alleles, conditional estimated effect size, standard error, and association p-value. Further, the table indicates whether each signal is specific to adaptive CD16+ NK cells, based on the absence of colocalisation with signals in CD16+ NK cells at the CT3 level, other cell types at the CT2 level, and non-blood GTEx tissues. For specific adaptive CD16+ NK signals colocalising with disease or quantitative-trait associations that did not colocalise with eQTLs in CT2 cell types or GTEx non-blood tissues, the table reports the study, phenotype, credible set ID and EFO label as comma-separated lists in matching order.

**Supplementary Table 26: Colocalisation and Mendelian randomisation results for CD16+ adaptive NK cell-specific eQTL signals with trait and disease associations**

Each row represents a colocalisation between adaptive CD16+ NK-cell-specific eQTLand disease or quantitative-trait associations not colocalising with eQTLs in CT2 cell types or GTEx non-blood tissues. The table reports the posterior probability that the two associations share a causal variant (PP.H4), the adaptive CD16+ NK-cell eQTL credible-set identifier, the gene Ensembl ID and symbol; the colocalising association is described by its study and phenotype identifiers, credible-set identifier, EFO term and label, and harmonised trait category. The table also reports the genetic variant used as the Mendelian-randomisation instrument and the corresponding Wald-ratio estimate for the effect of genetically predicted gene expression on the disease or quantitative trait.

**Supplementary Table 27: Summary of differential gene expression analysis for Genes & Health at CT2**

For each cell type 2 - trait combination considered in the differential gene expression analysis, we report the number of tested genes (Tested genes), the number of significant DEGs (Significant genes (FDR $< 0.05$) ), the total sample size (Tested individuals) and, when relevant, the number of cases (Number of cases) and controls (Number of controls).

**Supplementary Table 28: Summary of differential gene expression analysis for UK Biobank at CT2**

For each cell type 2 - trait combination considered in the differential gene expression analysis, we report the number of tested genes (Tested genes) and the number of significant DEGs (Significant genes (FDR $< 0.05$) ).

**Supplementary Table 29: Colocalisation of co-regulated gene expression modules with traits and diseases in G&H cohort**

In the table, nsnps indicates the number of variants included in each colocalisation test; PP.Hx.abf reports the approximate Bayes factor posterior probability for the corresponding hypothesis Hx (0/1/2/3/4); trait_study_id identifies the source from which the G&H GWAS summary statistics were obtained (i.e. “GH_gwas_ICD10codes” and “GH_gwas_quant_traits”); trait_phenotype_id identifies the G&H trait/disease GWAS phenotype (quantitative trait or ICD10 code); trait_cs_name is the unique identifier of the fine-mapped G&H trait/disease GWAS credible set; GH_module is the module identifier for grouping related G&H gene eQTLs credible sets; cell_eGene reports the cell type(s) and eGene(s) (Ensembl ID) included in the G&H module, comma-separated.

**Supplementary Table 30: Data sources for GWAS summary-statistics for the 46 manually curated disease datasets**

Data sources for the 46 manually curated disease datasets used in this study. In the table, phenotype type indicates if the trait is binary/case-control (cc) or quantitative (quant); sdY reports the standard deviation of the phenotype, when available (required for quantitative phenotypes); s represents the proportion of cases in the total GWAS sample (required for binasry/case–control phenotypes); and N corresponds to the total GWAS sample size (including both cases and controls for case–control studies).
